# Genome-wide association meta-analysis identifies novel ancestry-specific primary open-angle glaucoma loci and shared biology with vascular mechanisms and cell proliferation

**DOI:** 10.1101/2021.12.16.21267891

**Authors:** Valeria Lo Faro, Arjun Bhattacharya, Wei Zhou, Dan Zhou, Ying Wang, Kristi Läll, Masahiro Kanai, Esteban Lopera-Maya, Peter Straub, Priyanka Pawar, Ran Tao, Xue Zhong, Shinichi Namba, the Global Biobank Meta-analysis Initiative, Serena Sanna, Ilja M. Nolte, Yukinori Okada, Nathan Ingold, Stuart MacGregor, Harold Snieder, Ida Surakka, Cristen Willer, Alicia R. Martin, Milam A. Brantley, Eric R. Gamazon, Nomdo M. Jansonius, Karen Joos, Nancy J. Cox, Jibril Hirbo

**Author notes:** Corresponding author: Jibril Hirbo.

## Abstract

Primary open-angle glaucoma (POAG) is a leading cause of irreversible blindness globally. There is disparity in POAG prevalence and manifestations across ancestries. We identify novel and unique genetics that underlie POAG risk in different ancestries by performing meta-analysis across 15 biobanks (of the Global Biobank Meta-analysis Initiative) with previously multi-ancestry studies. 18 novel significant loci, three of which were ancestry-specific, and five sex-specific were identified. We performed gene-enrichment and transcriptome-wide association studies (TWAS), implicating vascular and cancer genes. A fifth of these genes are primary ciliary genes. Extensive statistical analysis of genes in the *SIX6* and *CDKN2B-AS1* loci (implicated in POAG, cardiovascular diseases and cancers) found interaction between *SIX6* and causal variants in chr9p21.3, with expression effect on *CDKN2A/B*. We infer that some POAG risk variants may be ancestry-specific, sex-specific, or both. Our results further support the contribution of vascular, cancer, and primary cilia genes in POAG pathogenesis.

## Introduction

Glaucoma is a complex eye disease characterized by a progressive loss of optic nerve (ON), fibers, which manifests initially as visual field loss, and if untreated ultimately leads to irreversible blindness.^1^ Primary open-angle glaucoma (POAG) represents the most prevalent type of glaucoma. POAG affects the trabecular meshwork (TM), the inner retina with retinal ganglion cell axons forming the optic nerve, and, presumably due to transsynaptic degeneration, the visual pathways including the visual cortex.^2, 3^ High intraocular pressure (IOP) is a major risk factor identified in POAG patients.^4^ Other risk factors are advanced age and positive family history, and difference in prevalence has been shown for POAG across ethnicities and gender.^5–7^ Moreover, there is disparity in POAG clinical presentations and outcome across ancestries.^8,9^

Through genome-wide association studies (GWASs), significant progress has been made in understanding the genetic pathophysiology of glaucoma in humans. In addition, animal models have also provided a valuable resource for understanding the biological mechanisms.^10^ However, there is still a lack of understanding of the underlying pathologic mechanisms in POAG, limiting the development of specific interventions in patients.

Three hypotheses have been suggested to explain the mechanisms causing the optic nerve damage (OND) that leads to glaucoma: vascular, biomechanical, and biochemical. The vascular theory posits that OND is due to unstable ocular perfusion which leads to ischemia of the optic disk and retina. In the biomechanical theory, elevation of IOP, due to an increase in TM outflow resistance, impinges on the ON fibers and thus causes subsequent retinal ganglion cell death.^11–13^ The biochemical theory attributes OND to neurotoxic biological molecules, such as excitatory amino acids, caspases, protein kinases, oxygen free radicals, nitric oxide, and tumor necrosis factor-alpha.^12^ However, the etiology that underlies POAG may actually involve interactions among the three theories.

Several studies have found that POAG is associated with a variety of cardiovascular diseases and vascular risk factors.^11,14–23^ In a previous large multi-ancestry GWAS,^24^ results from gene-enrichment analysis have implicated perturbation of molecular mechanisms in the vascular system that contribute to blood vessel morphogenesis, vasculature development, and regulation of endothelial cell proliferation. Analysis in large electronic health records (EHR), coupled with a zebrafish-model system, showed association of reduced genetically predicted expression of a gene that encodes for glutamate receptor GRIK5, which potentially determines blood vessel numbers, integrity in the eye, and increased vascular permeability, with comorbid vascular and eye diseases.^25^ However, no study has previously performed detailed exploration of the genetics that underlies the potential vascular connection with POAG across ancestries. Large population-based and clinical based biobanks offer an opportunity to do large-scale *in silico* investigations to elucidate common molecular systemic pathways between POAG and vascular systems.

The Global Biobank Meta-analysis Initiative (GBMI) is a collaborative group that currently involves 19 biobanks from 12 countries spanning four different continents: North America (Canada, USA), East Asia (China, Japan), Europe (Iceland, Estonia, Finland, Netherlands, Norway, Scotland, UK) and Australia. Each GBMI-affiliated biobank has paired genetic and phenotypic data collated through different types of electronic health data, such as self-report data from questionnaires, billing codes, doctors’ narrative notes, and death registry for >2.1 million individuals representing diverse ancestries: African ancestry individuals (AFR), Admixed Americans (AMR), Central or South Asians (CSA), East Asians (EAS), Europeans (EUR) and Middle Eastern (MID). Detailed description of each biobank is found in Zhou et al., 2021.^26^ We conducted a large-scale meta-analysis of POAG GWASs from 15 GBMI biobanks from six ancestries (n=1,487,447). We leveraged the largest, most diverse data to date combined with sophisticated statistical methods to identify unique molecular actors across ancestries and to determine the biological connections of POAG with vascular dysregulation.

## Results

### Discovery of novel ancestry-specific POAG loci

We report here a multi-ancestry genome-wide association study of 26,848 primary open-angle glaucoma cases and 1,460,593 controls from 19 biobanks. By analyzing all the biobanks together, a total of 62 loci that reach the genome-wide significant threshold were identified (Figure 2A, Supplementary Table S3; Supplementary Figure 5). Six loci were not previously reported for POAG, and encompass the genes *F5*, *RPL37A*-*LINC01280*, *ZFP91-CNTF*-*GLYAT*, *KALRN*, *CCDC13* and *MIR2054-INTU* (Table 1). Of these six loci, the latter four were replicated in an independent glaucoma cohort (Figure 2B, correlation between the effect sizes r = 0.71).^24^ A locus defined here as *ANGPTL7-MTOR* encompasses previously reported rare variants and corresponds to rs143038218*_UBAID1* locus.^27,28^ A Bayesian cross-ancestry meta-analysis identified three additional loci, one of which is specific to African ancestry and appears to be a novel locus (rs77136907_*MYO1B;NABP1,* p=2.74e-08), using Meta-Regression of Multi-AncEstry Genetic Association (MR-MEGA)^29^ (Suppl. Table S7).^26^ Three previously reported loci and identified here might be due to other glaucoma subtypes (Supplementary information). Therefore, we cannot rule out potential effect of non-POAG glaucoma signals i.e especially due to primary angle closure in EAS populations where the glaucoma subtype is common,^6,30–32^ or from biobanks where phenotyping was based on self-reporting (Supplementary information).

**Figure 1:**
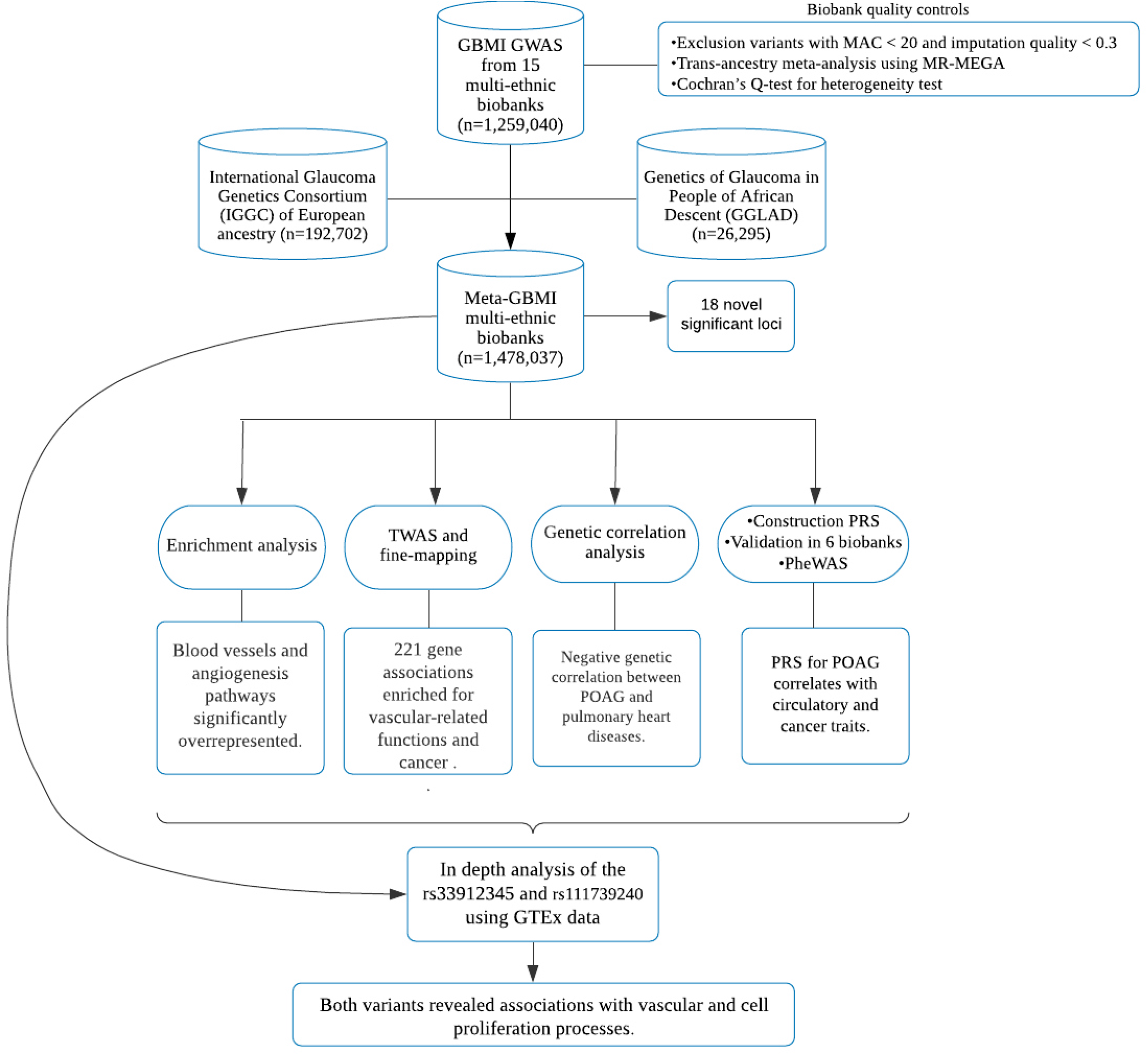
Workflow of this study. A total of 15 biobanks joined the GBMI multi-ancestry POAG meta-analysis, phenotyping was harmonized across the biobanks, biobank-specific quality control was performed and standardized genome-wide association study (GWAS) was conducted. To gain more discovery power to identify additional variants associated, a meta-analysis was performed among GBMI, IGGC of European ancestry and GGLAD. On the Meta-GBMI multi-ethnic biobanks, we performed functional impact, enrichment analysis, transcriptome-wide association study (TWAS), fine-mapping and genetic correlation analyses. Polygenic risk scores (PRSs) for POAG were constructed from the leave-biobank-out GBMI-IGGC-GGLAD meta-analysis summary statistics with PRS-CS and validated in six biobanks (BBJ, BioVU, Estonian, GLGS, Lifelines and UKBB). PRSs were then tested for association using phenome-wide association studies (PheWAS) across four biobanks. Then, to confirm and interpret our results, we examined the expression effects of missense variants in *SIX6-CDKN2B-AS1* and *TMEM167B* loci in GTEx data. MAC= minor allele frequency.

**Figure 2A:**
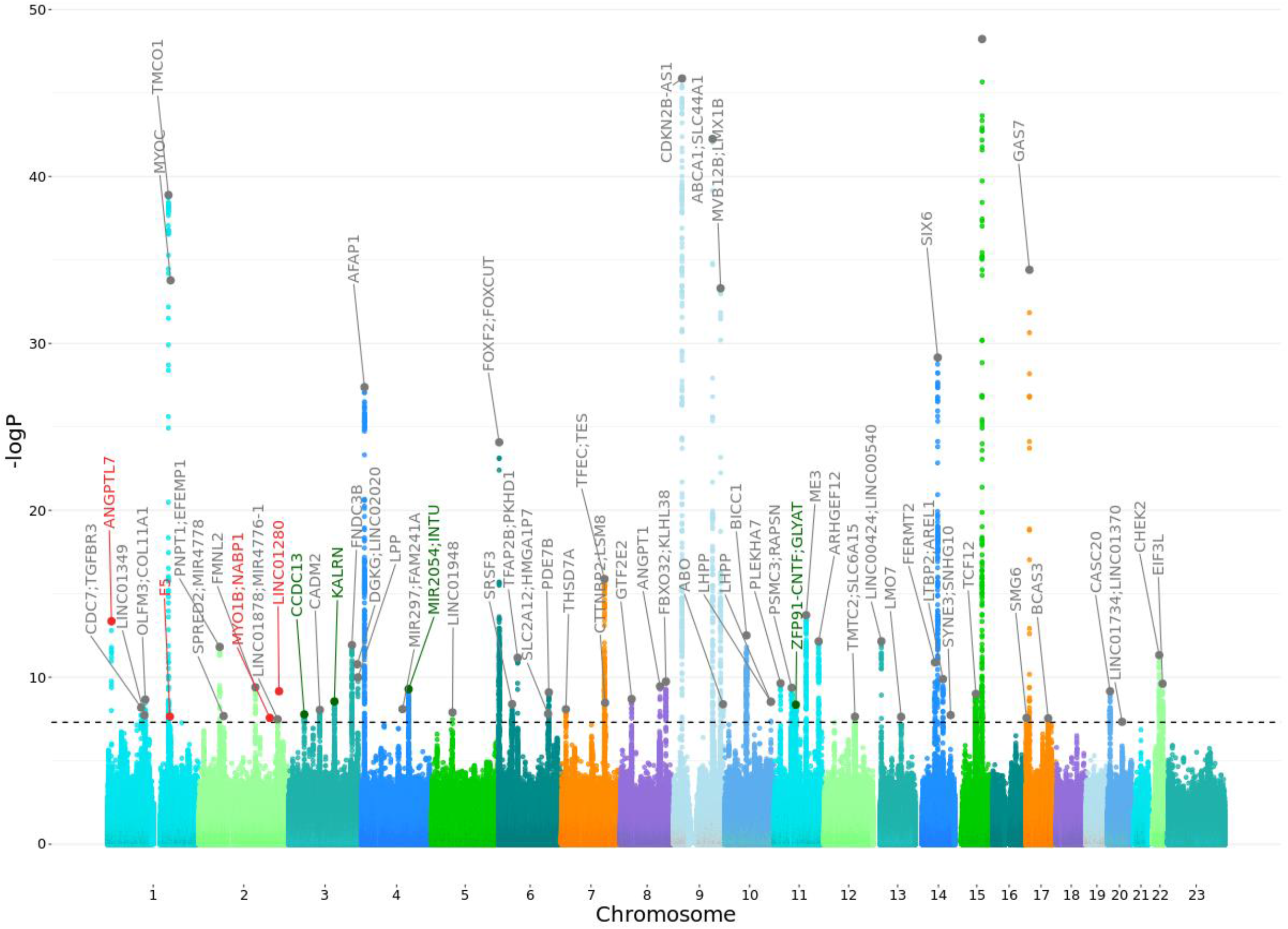
Manhattan plot of POAG in GBMI. The X-axis is the position on each chromosome and the Y-axis is the -log10 P-value from the GWAS for each SNP. The black line indicates the threshold for significance (P= 5e-8) after adjusting for multiple testing (Bonferroni adjustment). The four replicated novel regions and the nearest genes that reach the threshold for significance are indicated in green, in red are indicated the regions and the nearest genes that are potentially novel. In gray are reported the genes that are already known. Details of all genome-wide significant signals are in Suppl. Table S3.

**Figure 2B:**
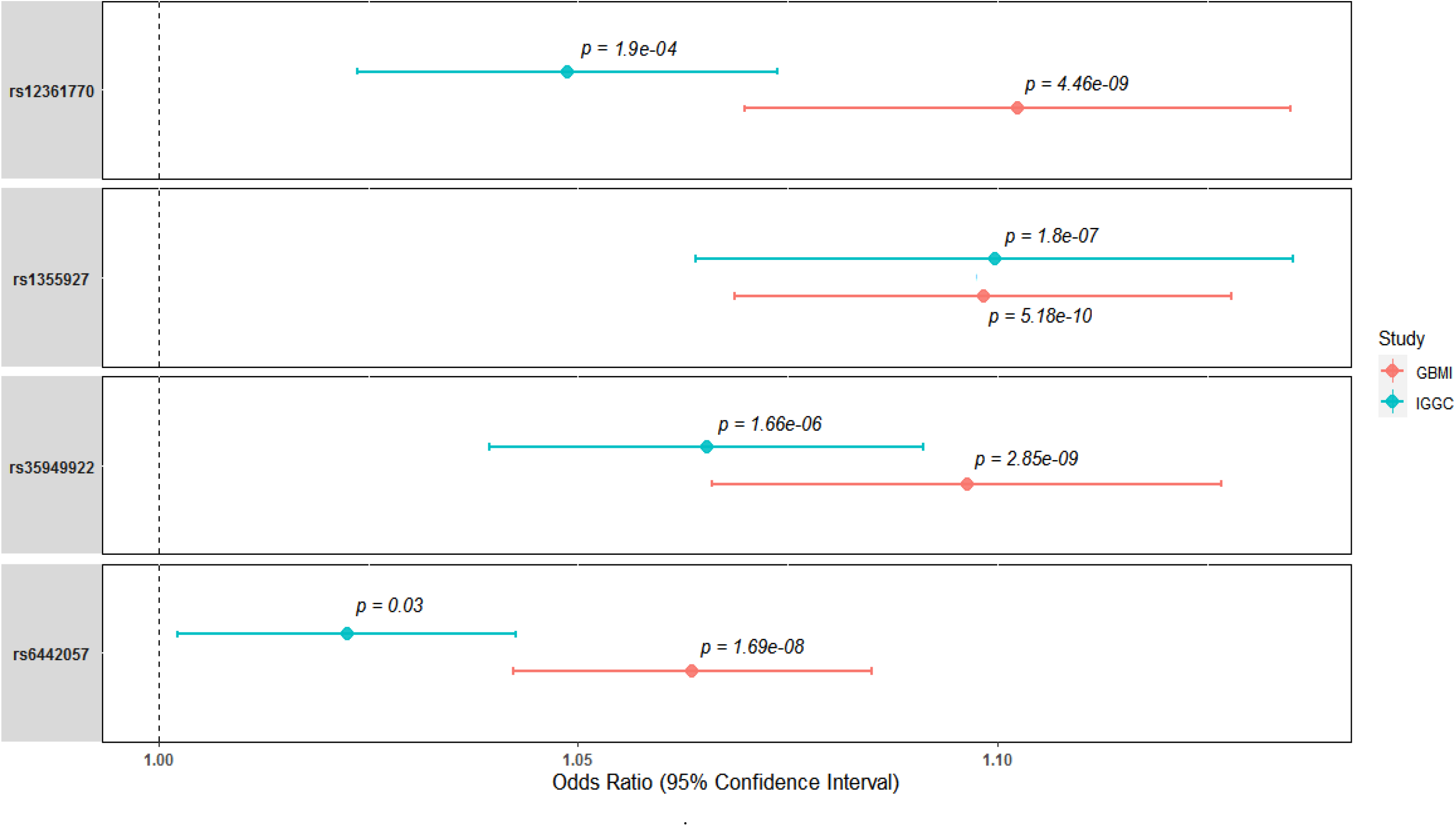
Effect size comparison of significant novel single nucleotide polymorphism in GBMI and IGGC. *p* indicates the p-value of each variant.

**Table 1:** Novel loci significantly associated with POAG (P < 5e-8) in the GBMI.

| Chr | Bp | Effect allele | Other allele | SNP | Gene | Function | N cases | N controls | MAF | Beta | Se | P | Hp | P in IGGC |
| --- | --- | --- | --- | --- | --- | --- | --- | --- | --- | --- | --- | --- | --- | --- |
| 1 | 169522686 | A | G | rs1469837390 | *F5* | intronic | 9505 | 327641 | 0.002 | 1.09 | 0.19 | 2.28E-08 | 0.01 | NA |
| 2 | 191664446 | C | G | rs77136907 | *MYO1B*;<br>*NABP1* | intergenic | 3134 | 641377 | 0.001 | 0.58 | 0.13 | 2.74E-08 | 0.00004 | NA |
| 2 | 216590253 | T | C | rs12476634 | *LINC01280*;<br>*RPL37A* | ncRNA,<br>exonic | 2538 | 484482 | 0.001 | 1.04 | 0.16 | 6.91E-10 | 0.03 | NA |
| 3 | 124587660 | T | C | rs35949922 | *KALRN* | intronic | 16658 | 1184946 | 0.18 | 0.09 | 0.01 | 2.85E-09 | 0.05 | 1.66E-06 |
| 3 | 42753558 | A | G | rs6442057 | *CCDC13* | intronic | 26848 | 1460599 | 0.26 | 0.06 | 0.01 | 1.69E-08 | 0.7 | 3.10E-02 |
| 4 | 127549411 | T | C | rs1355927 | *MIR2054*;<br>*INTU* | intergenic | 26848 | 1460599 | 0.07 | 0.09 | 0.01 | 5.18E-10 | 0.1 | 1.80E-07 |
| 11 | 58666759 | C | A | rs12361770 | *ZFP91-CNTF*;<br>*GLYAT* | intergenic | 17789 | 1213831 | 0.15 | 0.09 | 0.01 | 4.46E-09 | 0.4 | 0.000195 |

Abbreviations: Chr-Chromosome, BP-basepair position (hg38), SNP Single nucleotide polymorphism, -N number -MAF Minor allele frequency, -Se Standard error, -P P-value, -Hp Heterogeneity p-value, -NA Not present

To evaluate the cross-ancestry consistency in effect sizes, we estimated the correlation between betas of independent genome-wide significant SNPs between European, African and Asian populations in the GBMI dataset. The results showed the highest correlation between the European and Asian ancestry (Pearson correlation coefficient (r) = 0.87), while correlation between African with to other continental populations was much lower: European and African ancestry (r = 0.34), and African and Asian ancestry (r = 0.25) (Suppl. Figures S6-S8). However, three loci showed statistical heterogeneity in effect between ancestry and biobanks: novel rs12476634_*LINC01280* (in African and Hispanic Americans - African-Specific henceforth), well known rs74315329_*MYOC* (Europeans, European-specific henceforth), novel rs1469837390*_F5* (FinnGen & EstBB) and rs147660927*_ANGPTL7-MTOR*^27^(HUNT, FinnGen, EstBB, Lifelines, QSKIN) (Northern European-specific henceforth; Suppl. Table S3, S7).^26^ The well-known *MYOC* p.Gln368Ter variant, which hitherto had not been defined as ancestry specific, shows association in GBMI European-ancestry biobanks only.^33^ Additionally, >92% (289/314) carriers of this allele in the latest version of gnomAD database are of European ancestry, and the few that are reported in populations of other ancestries, have history of admixture with European populations. Furthermore, in 1000 genomes project this variant is present in Europeans and an individual of South Asian ancestry sampled in the UK.^34,35^ The three novel loci and newly defined locus that we classify as either African- or northern European-specific have similar patterns that are highly-specific to the two ancestries described here.^34,35^ The lead variants in northern-European-specific loci are only reported in Finnish and non-Finnish European individuals in gnomAD, while the lead variant in African-specific loci are observed in this study and the gnomAD/1000 genome databases in individuals of African and Hispanic Americans, who are generally African-admixed.^36–38^ In addition, 14 loci show significant effect size differences between ancestries based on Cochran heterogeneity test (Suppl. Table S7).

These ancestry specific loci and those that show heterogeneity between the ancestry might be driving most of the overall heterogeneity observed between the biobanks (Suppl. Figure S1).^26^ Phenotypic heterogeneity might also be a contributing factor with all self-reporting biobanks clustering together (Suppl. Figure S1) relative to those that used ICD-codes to identify cases. We did not find any EAS (BBJ and TWB) specific loci that demonstrated significant evidence for heterogeneity between the ancestries (Suppl. Table S7).

### Identification of novel sex-specific loci

We performed analyses separately by sex to identify any variants that demonstrated sex-specific association, or effect size heterogeneity by sex for the aforemention index variants. Sex-stratified GWAS identified one low-frequency African-specific association in females rs116625313_*PRKG2;RASGEF1B* (females p=2.85e-8, beta=1.52 vs males p=0.35, beta=-0.59) (Suppl. Table S4). In addition, four novel loci with POAG-association specific to males were identified (Suppl. Table S5) with low-frequency lead variants, three of which are African-specific (2-6% in population frequencies): rs111739240_*TMEM167B-C1orf194* on chromosome 1, rs17057880_*MIR3142HG-ATP10B* on chromosome 5 and rs114598725_*ARMC4* on chromosome 10, and one is European-specific, rs150385013_*LINC02024-LOC105374060* on chromosome 3 (>0.1% population frequencies, Suppl. Table S4-S5).

We next searched for coding variants at associated loci that explain the association signal and therefore implicate a highly-likely functional gene. The novel rs111739240_*TMEM167B-C1orf194* variant is in perfect LD (r^2^=1) in African ancestry populations with rs74113753 *C1orf194* missense variant that has also significant but slightly weaker association signal in our study and considered deleterious and possibly damaging by the SIFT and LoFtool, respectively.^39^ A proxy variant, rs17641032, that tags the missense variant (r^2^=0.7 in 1000genomes Africans) is at 7% and 3% frequencies in European and East-Asian population, respectively. However, the proxy variant is associated with POAG in African ancestry BioVU male subjects (AFR n=69 p=5.05e-4) but not in European ancestry subjects (EUR, n=213, p=0.892), confirming African-specific association of these locus. The proxy rs17641032 variant is a GTEx eQTL for *TMEM167B* and *AMIGO1* in eight tissues and one tissue, respectively.^40^ In GTEx tissue with the largest sample size, muscle skeletal (but not among the eight tissues reported in GTEx), the variant is associated with expression changes for three genes (*TMEM167B* p=0.00027, *CELSR2* p=0.0086 and *AMPD2* p=0.041). However, only *CELSR2* has male-specific expression changes at this variant (males p=0.0063, females p=0.38) in GTEx muscle skeletal tissue (Suppl. Figure S10). While the other two genes have weakened signals in both genders as expected due to reduced sample sizes in sex-stratified analysis in muscle skeletal and five more tissues out of seven, which had signals with nominal significant association out of all the 49 GTEx tissues (Suppl. Table S6). In addition, TWAS-PheWAS analysis revealed associations of the *CELSR2* with traits/phenotypes within the endocrine/metabolic (lipid traits) and circulatory groups with hyperlipidemia and angina pectoris, respectively, as the top phenotype associations (Suppl. Table S22).

In addition, 14 of the loci that have association signals in combined both-sex meta-analysis have significant albeit attenuated signals, only in either male (3 loci) or females (11 loci) in sex stratified analysis (Suppl. Table S4, S5). Four of these loci (near *CADM2, DGKG, KALRN* and *ARHGEF12*) show significant effect size differences between males and females based on Cochran heterogeneity test (Suppl. Table S4, S5, Suppl. Figure S9). All four genes prioritized using TWAS that are near lead variants for the four loci that show significant heterogeneity between males and females (*CADM2, DGKG, KALRN* and *ARGHEF2*), have also been shown to have sex-biased expression patterns.^41,42^

We examined differences in POAG prevalence between genders in BioVU subjects who are ≥ 40yrs and self-identify as European Americans (EA) or African (AA) ancestry. In 12,755 POAG cases in overall total of 1,372,397 BioVU subjects, there was higher odds of POAG in AA males relative to females (OR=1.15, CI 1.06-1.24, p=3.85e-4) while the risk was marginally lower in males than females in EA (OR=0.96, CI 0.92-1 (p=0.048).

We further performed meta-analysis of GBMI and public datasets from IGGC and GGLAD, generating the largest and most diverse GWAS to date, to identify additional loci that are associated with POAG. Ten additional novel loci that encompass the following prioritized genes *LOC654841*, *KBTBD8, ADGRL3, DDIT4L, HMGXB3, KCNK5, MAD1L1, APPL2, CATSPERB,* and *FENDRR* attained genomewide significance, all of which were associated with POAG at nominal significance in the GBMI and IGGC data (Suppl. Table S7, Suppl. Figure S11, S12). Five of these ten genes in the novel loci are involved in cardiovascular conditions and six in cancer processes.^43–55^ Three loci that were novel in GBMI, *ANGPTL7*, *CCDC13*, and *LHPP,* were attenuated to sub-genome wide significance level in GBMI-IGGC-GGLAD meta-analysis (p-values of 0.0043, 6.202e-08 and 2.506e-07, respectively, Suppl. Table S3, S7, S12)

### Enrichment analyses prioritize vascular and cell proliferation mechanisms

To identify the functional roles of POAG-associated variants and which tissues are mediating the genetic effects, we performed enrichment analyses with DEPICT using meta-analysis of GBMI-IGGC-GGLAD summary statistics (see methods section). DEPICT prioritized 101 co-regulated genes, of which 43 (43%) have been previously reported for POAG and 58 (57%) that were novel (Supplementary Table S8). Nearly 50% (49 genes) of the genes identified here were reported to be associated with vascular traits including cardiac disease, and arterial stiffness measurement while nearly a third (34 genes) were associated with cancer, including cutaneous melanoma, keratinocyte carcinoma, breast carcinoma, prostate carcinoma, and lung carcinoma (Supplementary Table S8). The gene-enrichment analysis was performed using the identified 110 biological pathways, reaching the Bonferroni corrected level of significance (p< 3.45e-6) (Supplementary Table S9). Among the top and common enrichment signals were biological pathways involved in vascular related blood vessel development/morphogenesis, angiogenesis and epithelial cell proliferation (Supplementary Table S8). In addition, several biological pathways crucial in cell development and proliferation like the IGF1 subnetwork, insulin-like growth factor binding and Wnt signaling were significantly enriched (Supplementary Table S9). Tissue/cell type enrichment analysis prioritized 10 significant tissues and cell types, with musculoskeletal and cardiovascular systems being the most represented (Supplementary Table S10).

### TWAS Analyses identify novel associations

TWAS of the GBMI-IGGC-GGLAD meta-analysis summary statistics identified 19 gene-trait associations (p<2.5e-6, Bonferroni correction for mean 20,000 genes tested) in GTEx tissue biologically most relevant to ocular traits, brain cortex tissue (Suppl. Table S11). Most of these genes (16/19) showed associations in at least one other tissue among the additional 22 other GTEx tissues potentially relevant to ocular diseases (Suppl. Table S11-S13). The JTI model generated most genes and gene-trait associations among the three cis prediction models (Suppl. Table S12, S13, Suppl. Figure S13). However, the majority of the gene-trait associations in JTI overlap with what was obtained using PrediXcan and UTMOST models (Suppl. Figure S13). Using the three cis models made it possible to have a comprehensive TWAS prioritization of gene-trait associations, for example one additional gene-trait association that transect novel GWAS loci identified in this study was prioritized using PrediXcan and UTMOST models but not the most robust JTI (Suppl. Table S13).

Overall, 221 out of 271 gene-trait associations across the 23 tissues were estimated to define the 90% credible set at the locus via probabilistic fine-mapping. 116 of the gene-trait associations in the 90% credible set intersect with 57 out of the 103 GWAS loci identified. A total of 14 of these gene-trait associations were novel, nine of which were unique to GBMI (Suppl. Table S12, S13). 156 gene-trait associations intersect 64 previously known loci, while 86 did not intersect with any of the genome-wide significant risk loci and correspond to 67 loci with subgenome-wide GWAS signals (TWAS ‘loci’) that will potentially attain significance in a more powered GWAS (Suppl. Figure S13, Suppl. Table S12, S13). For example, *SEC31B* (SEC31 Homolog B, COPII Coat Complex Component), a gene on chromosome 10, which is a novel significant TWAS association signal in three GTEx brain tissues using the JTI model (Suppl. Table S12). The SNP located near this gene, rs11190559, was the strongest SNP (p=2.4e-5) with 47 other variants in the locus with association signal below 1e-4 (Suppl. Table S12,S13). The *SEC31B* gene has been reported in the context of two anomalous pigmentary syndromes with ocular manifestations; retinitis pigmentosa 37 and Hermansky-Pudlak syndrome 1.^56^ A GWAS variant in the vicinity of the gene is associated with heart rate.^57^ Moreover, loss-of-function mutations in *SEC31B* promote colorectal cancer metastasis and a rare form of anemia.^58,59^

Seven of the novel TWAS ‘loci’ were unique to PrediXcan and/or UTMOST models (Suppl. Figure S13, Suppl. Table S13). Overall, nearly two thirds of the genes cumulatively prioritized using the three cis TWAS models (181/271) have vascular-related and/or cell senescence/proliferation functional roles, or have been implicated in vascular or neoplastic diseases. A fifth of these vascular and cancer related genes are cilia-related genes (Suppl. Table S14).

We confirmed that all but four TWAS-prioritized genes (14/18) that transect novel loci associated with POAG are robustly expressed in all eye tissues based on publicly available Ocular Tissue Database (OTD).^60,61^ However, the four genes missing in OTD: *ABHD18*, *ACKR2*, *LAMTOR3* and *ALDH1L2* have been shown to be expressed in mouse and pig retina.^62,63^

### PRS prediction performance and effect of POAG liability across EHR

Prediction performance of the leave-biobank-out GBMI-IGGC-GGLAD POAG meta-analysis as discovery GWAS in four biobanks with European ancestry subjects as estimated by *R*^2^ on the liability scale ranged from 0.0166 (95% CI 0.01, 0.025) for Lifelines to 0.0484 (95% CI 0.042, 0.056) for UKBB (Figure 3A). However, consistent with previous findings, performance was lower for the two non-European ancestry populations (Figure 3A).^64^ Results for the more balanced case-control ratio of ∼1:4 in BioVU and Lifelines showed an improvement in the *R*^2^ (Supplementary Figure S14).^65–68^ Similarly, European ancestry subjects with polygenic risk scores (PRSs) in the highest risk decile of the PRS distribution had 2.1 to 4-fold higher odds of POAG compared with those in the mid decile (95% CI=1.8,2.24 and 3.34,4.79). In African ancestry individuals the odds ratio between the highest and mid decile polygenic risk was slightly lower but significant [2.3-fold (95% CI=1.21,4.97)] and in East-Asian samples 1.72-fold (95% CI=1.59,1.85) (Figure 3B). Further, the PRSs were robustly associated with POAG phecode across all biobanks (Suppl. Table S15-21).

**Figure 3A:**
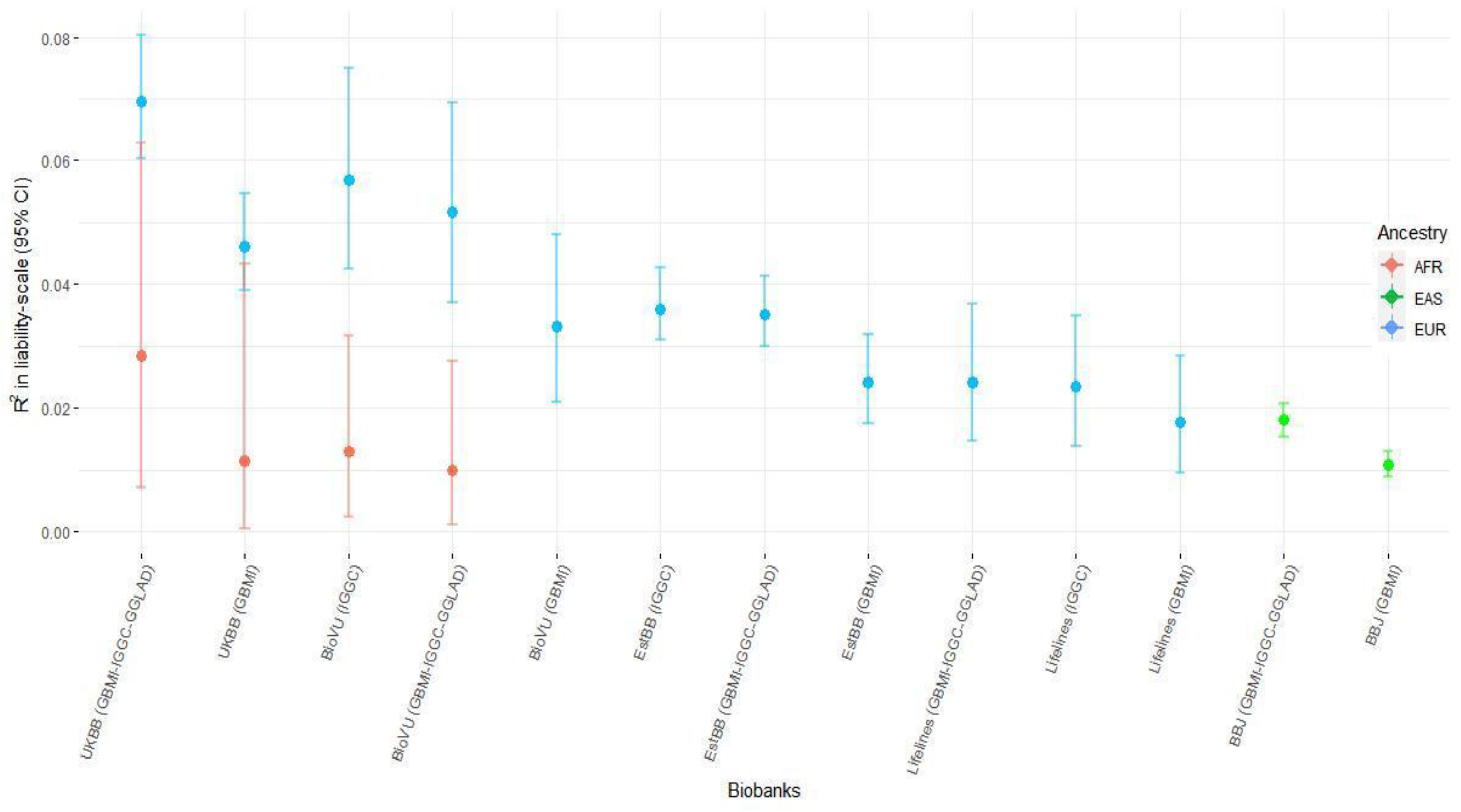
Prediction performance of GBMI-IGGC-GGLAD and IGGC POAG meta-analysis across 5 biobanks. The proportion of variance in POAG explained by PRS (Nagelkerke’s R² in liability scale) is reported in the y-axis. In parentheses for each biobank is the source dataset used to estimate the marginal effect size of single nucleotide polymorphisms. The polygenic risk scores of POAG were generated using PRScs auto. Performance of UKBB and BBJ using IGGC was not checked because data from the two biobanks are part of IGGC meta-analysis. The proportion variance of African individuals in BioVu using GBMI was not significant. AFR= African ancestry, EAS= East Asian ancestry, EUR= European ancestry.

**Figure 3B:**
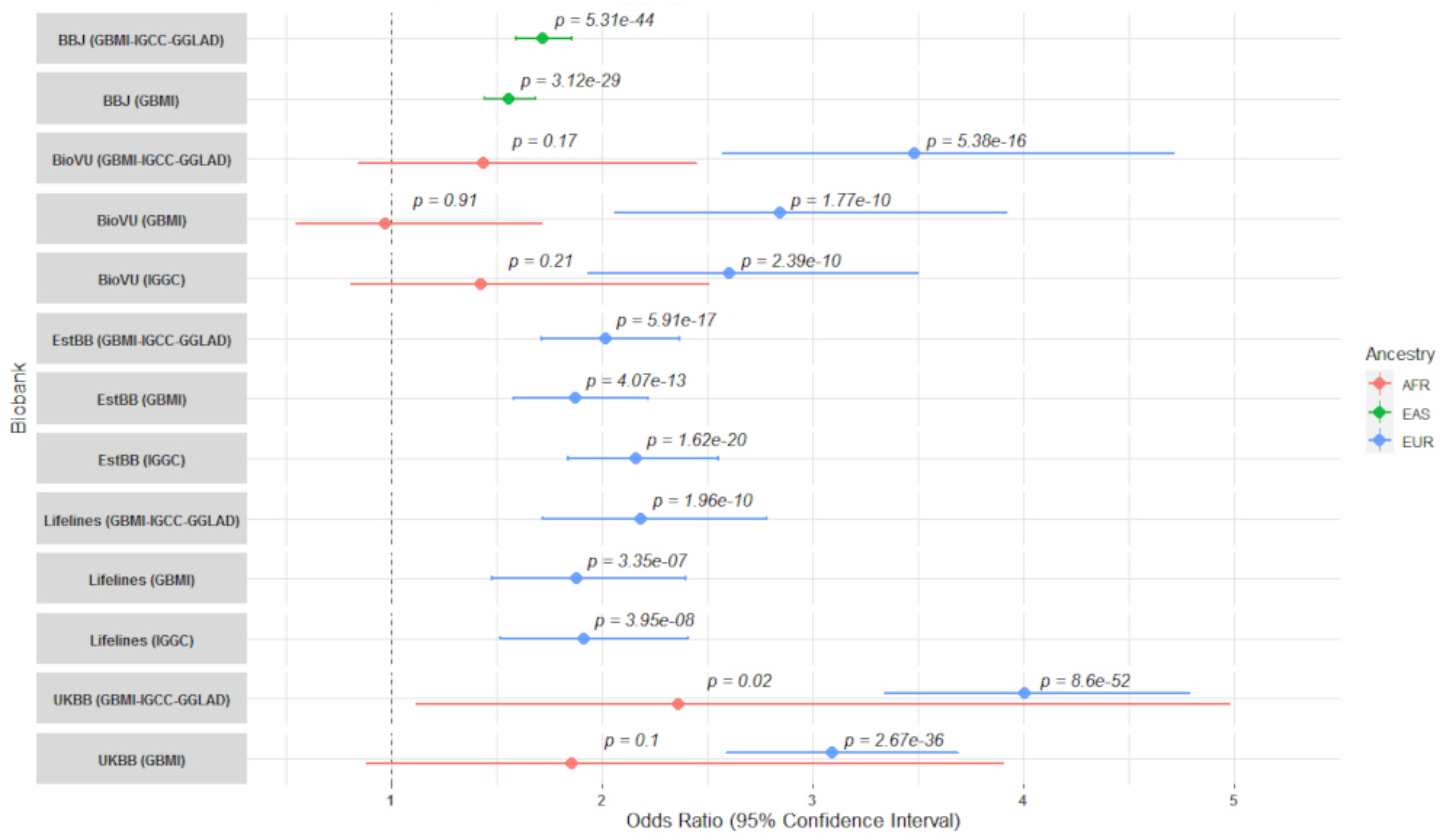
The odds ratio (OR) between top decile and mid-decile PRS POAG across five biobanks. The dashed line indicates OR=1. The averaged OR was calculated using the inverse-variance weighted method. PRS was stratified into deciles with the mid-decile (40-60%) used as the referenced group.In parentheses for each biobank is reported the discovery dataset used to estimate the marginal effect size of single nucleotide polymorphisms. AFR= African ancestry, EAS= East Asian ancestry, EUR= European ancestry.

**Figure 3C:**
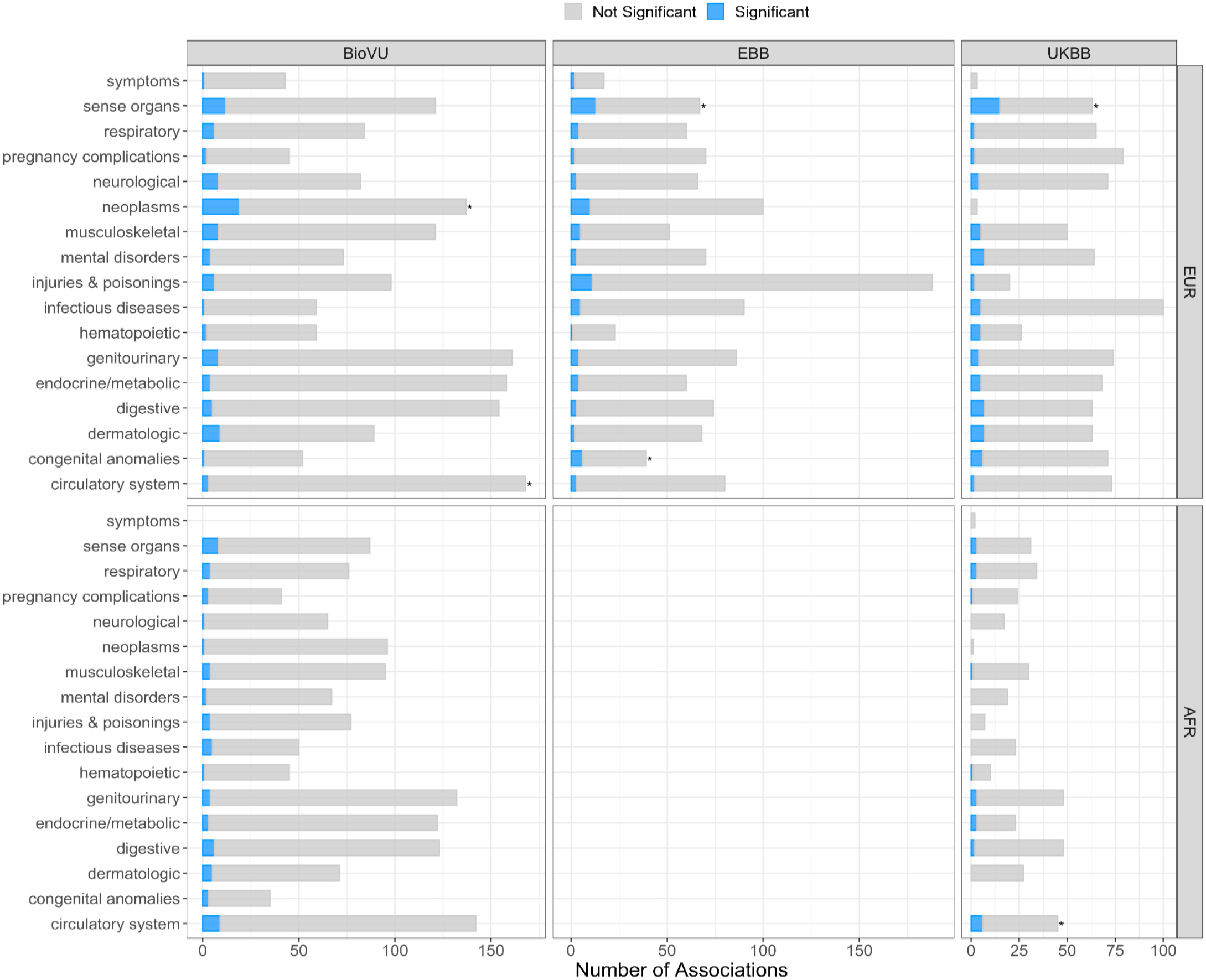
Proportion of phecode groups associated with polygenic risk generated from leave-biobank-out POAG GBMI-IGGC-GGLAD meta-analysis across three biobanks. Asterisks indicate groups that are significantly enriched.

In general, summary statistics from IGGC yielded better predictions and odds ratios across the five biobanks and the GLGS cohort, relative to GBMI data (Figure 3A, 3B, Suppl. Figure S14).^64^ Moreover, there was no significant improvement in predictions when using GBMI-IGGC-GGLAD meta-analysis summary data as a source relative to IGGC (Figure 3A). This could be due to phenotype heterogeneity and potentially unbalanced case-control ratio in GBMI relative to IGGC. These were exemplified by elevated prediction observed for the more accurately phenotyped GLGS cohort with balanced case-control ratio compared to equally balanced target data in BioVU and Lifelines (Supplementary Information, Suppl. Figure S14).

As expected, the genetic risk for POAG was associated with ocular traits, but also with other phenotypes, like circulatory, neoplasm and musculoskeletal traits, across the five biobanks (Figure 3C, Suppl. Table S15-S21). Additional association signals with circulatory traits and neoplasms were observed in African (eg. UKBB, Fisher’s test p=0.0376) and European ancestry (eg. BioVU p=0.0002) cohorts, respectively (Figure 3C, Suppl. Table S15, S18). We further explored POAG comorbidity pattern across circulatory phecodes (n=171, ≥ 100 cases) in a total of 1,968,903 BioVU subjects, African ancestry (n=273,379) and European ancestry (n=1,695,524), after excluding all individuals with other eye phecodes (phecode 360-379) and correcting for age and sex. African ancestry individuals who have POAG have significantly higher odds of comorbidity with a circulatory phecode relative to European ancestry individuals (OR=2.63, CI=1.66-4.16, p=3.4e^-5^).

### Genetic correlation analysis

We observed negative genetic correlation between POAG and all but one of the 11 vascular traits tested, atherosclerosis (Suppl. Table S23). However, only pulmonary heart diseases had statistically significant genetic correlation (Suppl. Table S23). The two neoplastic traits that were tested, prostate and breast carcinomas were positively correlated but not statistically significant.

### Interaction between the *SIX6* and *CDKN2B-AS1* loci

We confirmed that the haplotype containing the rs33912345 risk allele has reduced *SALRNA1* and *SIX6* expression relative to the haplotype containing the reference allele (Figure 4A-B), potentially indicating that the missense rs33912345 variant contributes to etiology of POAG by downregulating the expression of the genes in the *SIX6* locus.

**Figure 4A:**
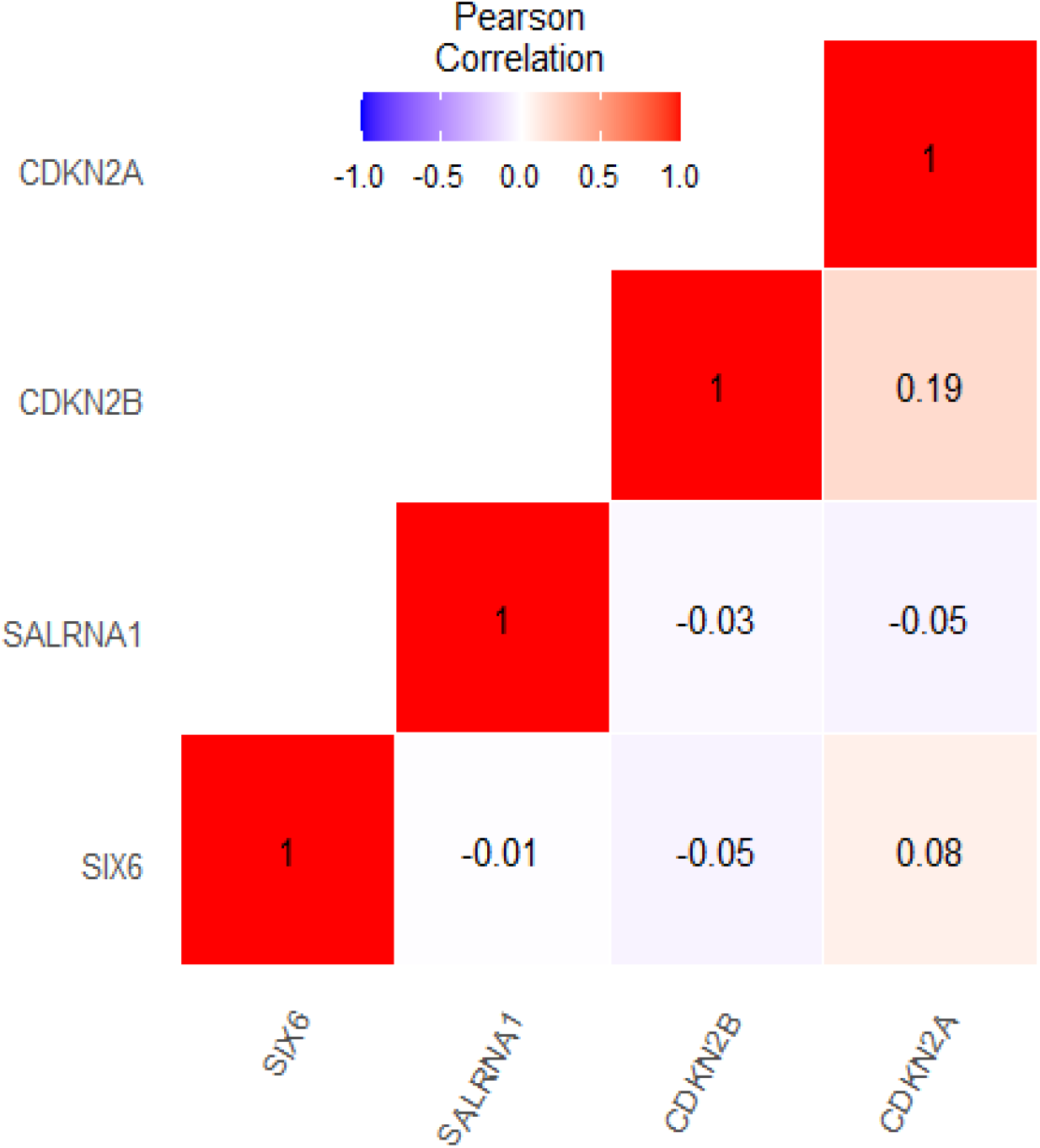
Correlations in GTEx muscle skeletal tissue measured gene expressions between genes in the SIX6 and CDKN2B-AS1 loci.

**Figure 4B:**
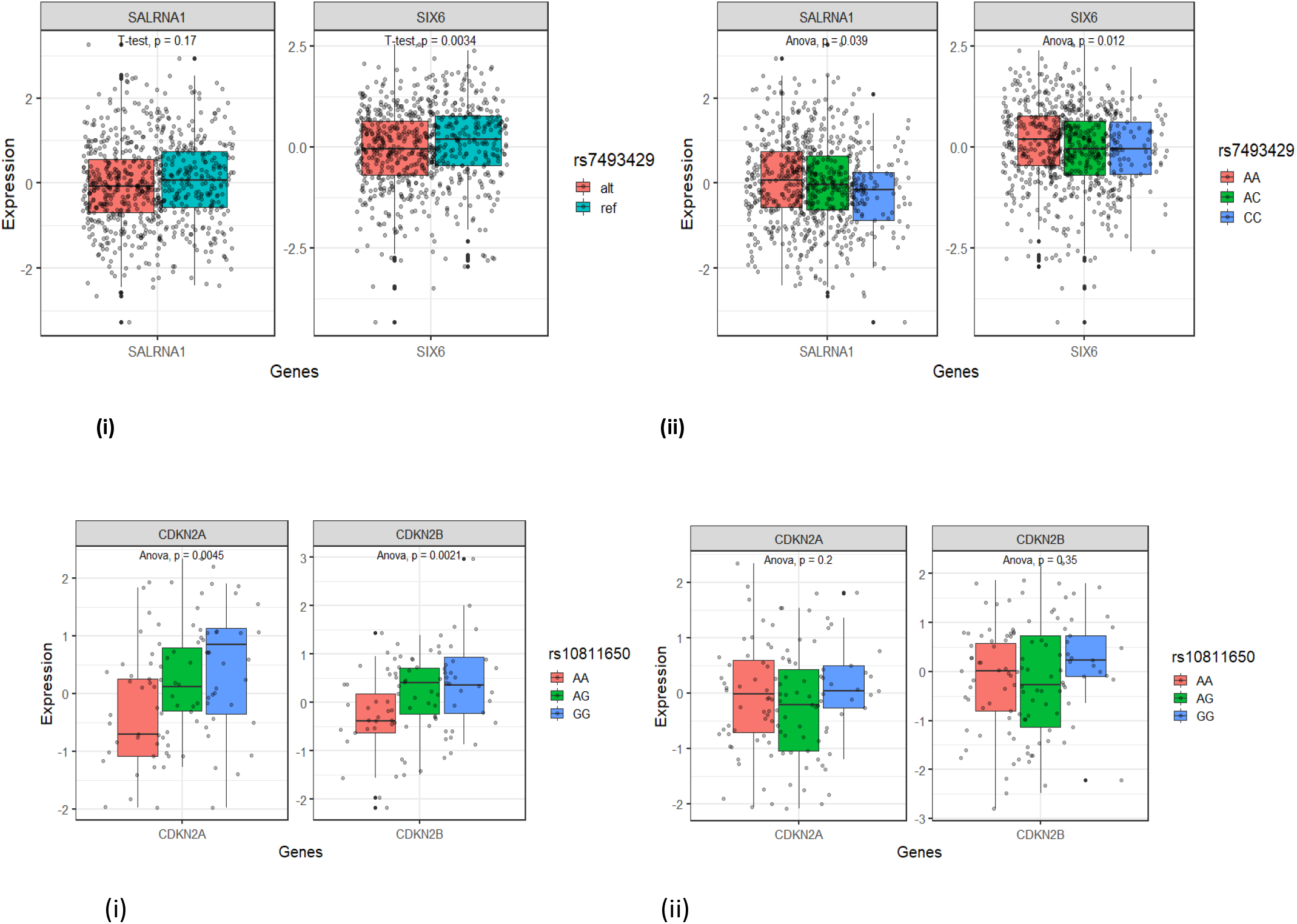
Effect of rs33912345 variants in GTEx measured gene expressions in Muscle skeletal tissue (n-716). Difference in GTEx measure gene expressions *SIX6* loci genes – *SIX6* and *SALRNA1* between i) individuals who carry the rs33912345C causal allele (represented here by proxy variant rs7493429, r2=0.74) versus those with the wild type allele ii) different genotype causal/wild type combination. p-values (p) are based on T-test (i) and ANOVA tests (ii).

**Figure 4C:**
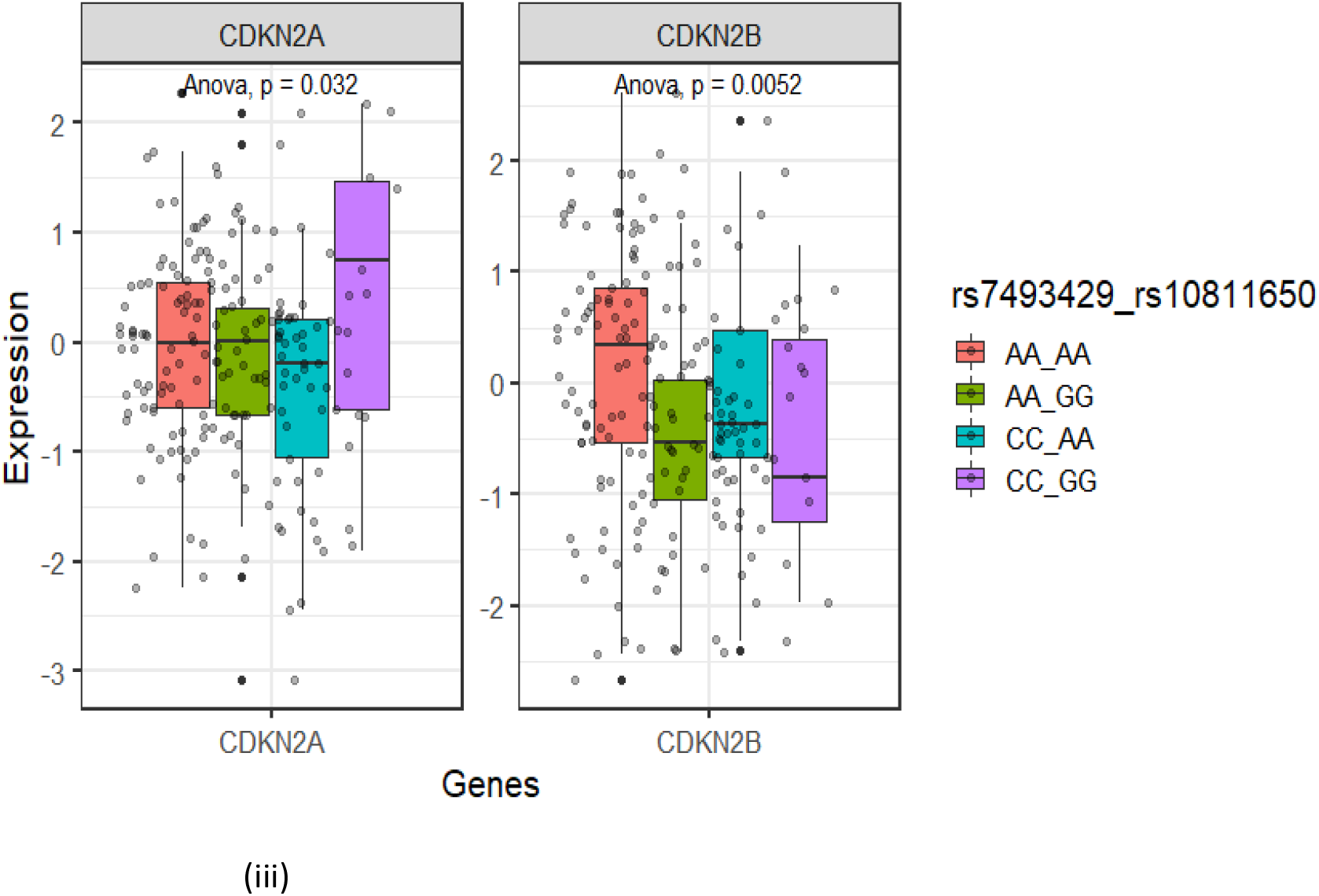
Effects of genetic interaction between rs33912345_*SIX6* proxy variant and rs10811650_*CDKN2B-AS1* on *CDKN2B-AS1* locus gene expressions. (**i**) Expression pattern in individuals who carry wild type rs33912345_*SIX6* proxy variant in combination with rs10811650_*CDKN2B-AS1* genotypes in brain cortex, (**ii)** expression pattern in individuals who carry rs33912345_*SIX6* proxy variant causal allele in combination with rs10811650_*CDKN2B-AS1* genotypes in brain cortex (**iii**) expression pattern in individuals who carry homozygous rs33912345_*SIX6* proxy variant in combination with rs10811650_*CDKN2B-AS1* genotypes in muscle skeletal. p-values (p) are based on ANOVA tests.

**Figure 4D:**
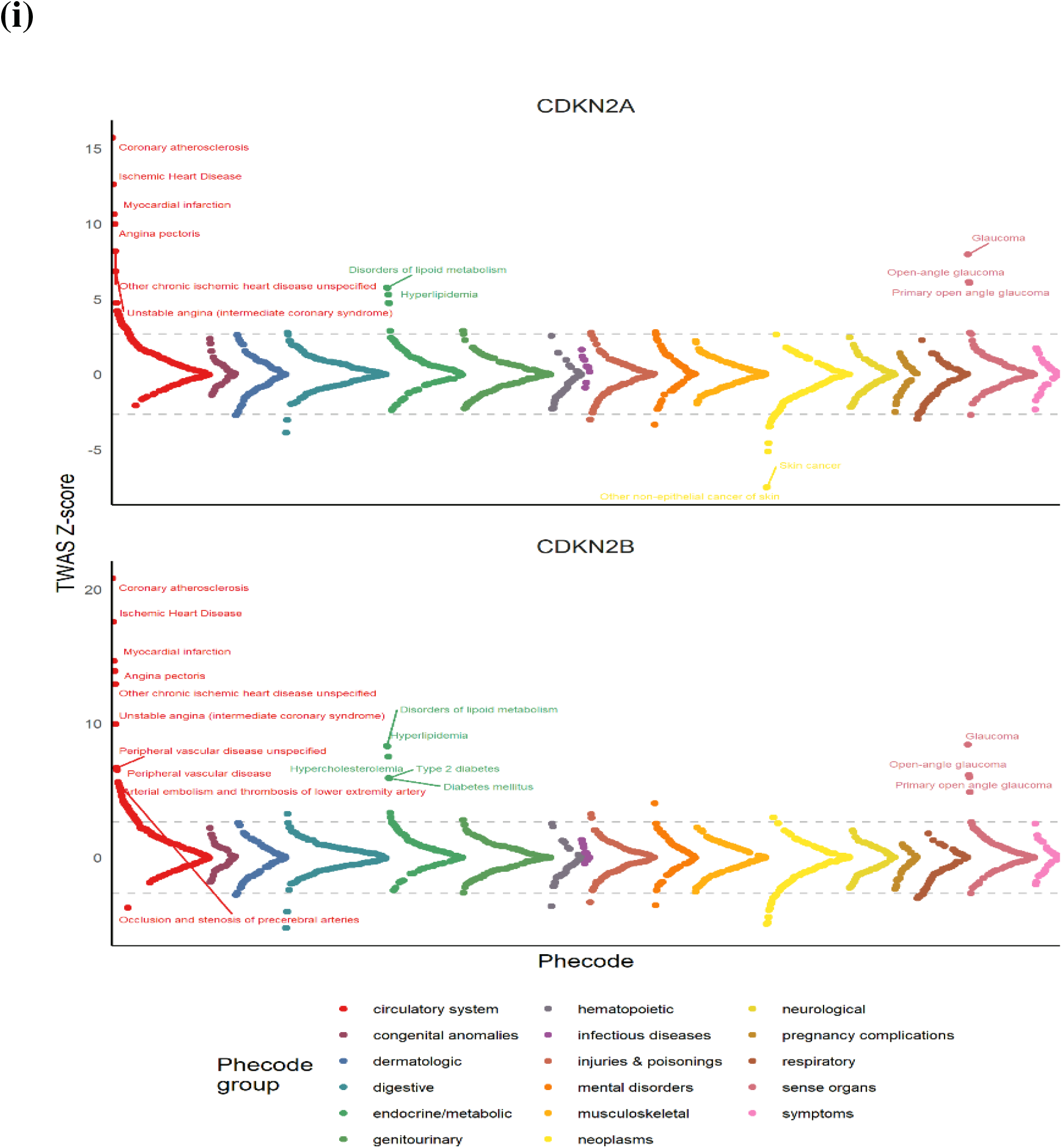
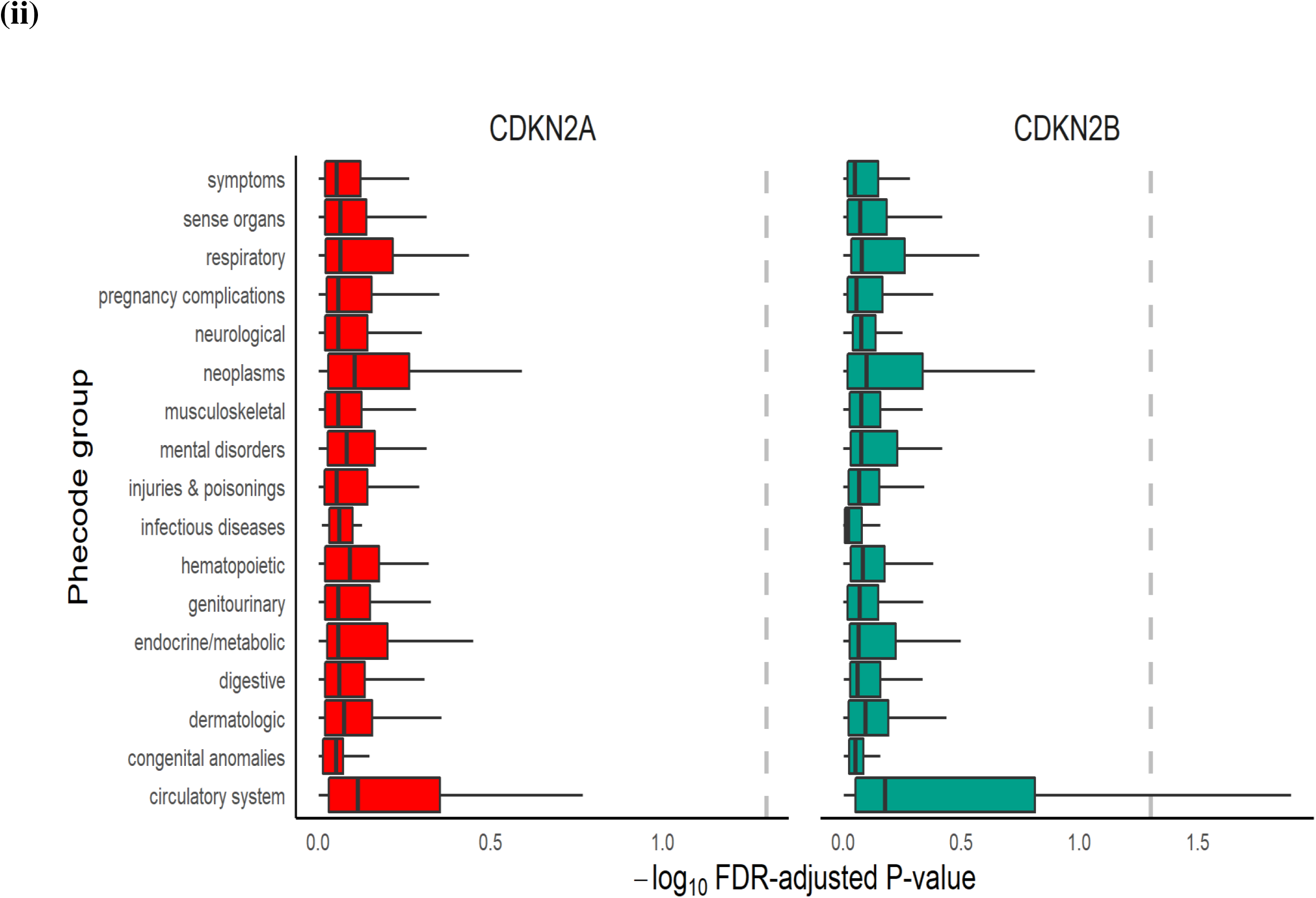
GReX-PheWAS for categorizing phenome-wide associations for CDKN2A and CDKN2B genetically-regulated expressions in BioVU and UKBB. (**i**) Miami plot of TWAS Z-scores (Y-axis) across phenotypes, colored by phecode group. The dotted gray line shows the significance threshold for Benjamini-Hochberg FDR correction and phenotypes are labeled if the association passes Bonferroni correction. (**ii**) Boxplots of -log_10_ Benjamini-Hochberg FDR-adjusted p-values of genetical GTAs across 9 phenotype groups. The dotted gray line shows FDR-adjusted p-value = 0.05.

To further study if there is an association signal in *SIX6* locus independent of rs33912345 missense variant, we rebuilt the prediction model in skeletal muscle tissue by excluding all variants in LD with the missense variant (r^2^<0.1). We found weak or no association signals with either *SIX6* (p=0.041 vs p=5.93e-15 for original model) or *SALRNA1* (p=0.777 vs p=1.96e-32 for the original model) with LD-constrained rebuilt PrediXcan and UTMOST models, respectively, indicating that all the association signals detected in the locus in the original gene models are attributable to the missense variant. Similarly, all the chr9p21.3 association signals observed with POAG are mainly attributable to exonic CDKN2B-AS1 rs1008878 variant (Suppl. Figure S15).

We next explored interaction between rs33912345 proxy variant and *CDKN2* genes, and potential consequences on the expression pattern of these chr9p21.3 genes. We first confirmed that there was correlation between expression levels of *SIX6* and chr9p21.3 loci TWAS prioritized genes in GTEx data in muscle skeletal tissue (Figure 4A, Suppl. Figure S16-18). We also observed interaction between the two loci for both POAG and vascular related traits (Phecodes 411, 411.1 - 411.9) in BioVU (Suppl. Table S24). We further found that the proxy variant has significant effect on expression of *CDKN2A* and *CDKN2B* via interaction with chr9p21.3 top GWAS variants that are associated with vascular and neoplastic traits in GWAS catalog (rs10811650 and rs2891168). The eQTL effect of the chr9p21.3 variants disappear in the presence of rs33912345 proxy causal allele (Figure 4B-C). However, in the presence of homozygous rs33912345 proxy, the eQTL effect is observed for those with homozygous causal alleles for the local variants (Figure 4B-C, Suppl. Figure S16-18).

We explored potential biological consequences of the interaction between the two loci by performing TWAS-PheWAS for the two CDKN2B-AS1 loci genes in the UKBB (n=396,618) and BioVU (n=59,805) cohorts. We applied the best performing model with the highest prediction ability estimated from the cross-validation for each gene among PrediXcan, UTMOST, and JTI.^69–71^ For the two chr9p21.3 genes the best performing models in brain cortex tissue were JTI (*CDKN2A*, r^2^=0.0257) and UTMOST (*CDKN2B*, r^2^=0.0413). We performed GReX-PheWAS for the two chr9p21.3 genes in the same two cohorts, followed by meta-analysis of the two PheWAS (n=456,423) across 731 traits and diseases, grouped into 17 categories. The GReX-PheWAS analysis revealed associations for *CDKN2A/B* in the *CDKN2B-AS1* locus with glaucoma and enrichment for phenotypes of the circulatory and neoplasm groups with coronary atherosclerosis and skin cancers as the top phenotype associations (Figure 4D). These phenotypes include traits that are a result of senescence (upregulation) and cell proliferations (downregulation). We also detected associations with lipid disorders, which have been implicated in both these two groups, consistent with the defined functions of the two genes, suggesting that an etiology that connects these traits is a balance between cell proliferation and senescence.^72,73^

## Discussion

In this study, we aimed to determine the genetic mechanisms underlying POAG. We leveraged a suite of statistical tools to analyze a combination of massive GBMI genome-wide discovery resources, previously published GWAS and publicly available GTEx data. In a large-scale GBMI GWAS meta-analysis for POAG across 15 biobanks (n=1,487,447) we identified a total of 62 risk loci, of which six were novel and four of these were replicated in IGGC.^24^ The replicated associations encompass loci near genes *INTU-MIR2054, KALRN,* and *ZFP91-GLYAT-CNTF, CCDC13,* with only the latter not confirmed by TWAS.^74–77^ Meta-analysis of GBMI, IGGC and GGLAD cohorts revealed additional novel associations in the loci that encompass *KBTBD8, LOC654841, ADGRL3, DDIT4L, HMGXB3, KCNK5, MAD1L1, APPL2-KCCAT198, CATSPERB* and *FENDRR* genes. TWAS prioritized six novel POAG susceptibility genes that correspond to five of the ten additional novel loci, and all have functional roles or have previously been implicated in vascular and/or neoplastic related pathologies: *COL4A4* ^78^*, MFF*^79^ *(LOC654841), LAMTOR3*^80,81^ *(DDIT4L), HMGXB3*^82,83^ *(HMGXB3), ALDH1L2*^84^ *(APPL2-, KCCAT198)* and *CCDC88C*^85–87^ *(CATSPERB).* By virtue of being gene-based and a more well powered analysis than GWAS, TWAS identified potentially 67 additional novel ‘loci’ that are at subgenome-wide GWAS association. ^58,59^ Overall, we leveraged multiple GWAS and TWAS methods in a massive multi-ancestry study, cumulatively identifying 176 POAG risk loci. Review of literature on functional roles of the genes that fall within these loci coupled with enrichment, genetic correlation and PheWAS analyses all point POAG risk to potential link to vascular and proliferation mechanisms.

Two thirds of all the TWAS-prioritized POAG associated genes identified in this study have previously been associated with vascular related traits and/or implicated in carcinoma and act as tumor suppressors. In addition, all the genes that are near or regulated by the sex associated novel loci are also implicated with risk of vascular related diseases, serum cholesterol metabolism and neoplasms.^42,88–94^ We performed enrichment analyses in which cell proliferation and angiogenesis pathways were significantly represented. In summary, enrichment analysis of the largest GWAS for POAG reported reinforced the role of vascular processes in glaucoma. The results also highlight pathways related to cell proliferation mechanisms that have not been reported in previous GWAS studies. Therefore, these results add more insights into understanding glaucoma pathogenesis. We further performed genetic correlation with vascular related GWAS traits and explored pleiotropy of genetic risk for POAG across the phenome in five biobanks, and confirmed association of POAG genetic risk burden with vascular and neoplastic related traits. Interestingly, consistent with our findings, an accompanying manuscript in this issue - on genetic drug discovery/repurposing for the GBMI POAG GWAS - identified four molecules approved for treatment of vascular-related conditions and two additional molecules approved for neoplasm related conditions.^95^ In addition, an anti-hyperlipidemic drug initially developed for the treatment of coronary artery disease, probucol, was prioritized.^95^

Previous studies showed disparity in POAG prevalence across ancestries and sexes. While non-genetic factors are likely to drive many of the observed health disparities characterized to date, differences in some underlying genetics including differences in frequency among populations, unique unobserved rare variants and/or difference in effect sizes at common SNPs play a significant role in differential disease etiology.^96^ In the meta-analysis on both sexes we observed three novel loci and two known POAG locus that showed ancestry-specific effects. Sex-stratified association analyses identified additional three African-specific novel loci that were associated with POAG in males only: *ATPB10*, *TMEM167B-C1orf194* and *ARMC4*, and one African-specific novel locus *PRKG2;RASGEF1B* that is associated with POAG in females only. *PRKG2,* the closest gene to the lead female associated variant, is induced by estrogen and progesterone.^97^ Interestingly, estrogen has been reported as having protective effects in glaucoma.^98^ 14 additional loci show significant effect size differences between ancestries, three of which also have significant difference in effect between males and females. Genes prioritized using TWAS that are near the lead variants for the loci that show effect difference between the sexes have sex-biased gene expression patterns.^41,42^ We further performed in-silico validation of the novel male associated African-specific *TMEM167B-C1orf194* locus and showed that the variant is a male-specific regulator of *CELSR2. CELSR2* has previously been reported to be differentially expressed between males and females and associated with cardiovascular traits.^42^ *ARMC4* has also been shown to be differentially expressed between males and females.^42,89,90^

Results from polygenic risk score PheWAS across the BioVU, UKBB and EstBB also suggest that genetic risk burdens of POAG are associated with vascular related traits, especially in African ancestry, and neoplasms in European ancestry populations. Moreover, we observed higher incidence of POAG comorbidity with vascular related traits in African ancestry relative to individuals of European ancestry. These potentially suggest unique shared biology between POAG and circulatory traits in African ancestry populations. It is not clear if our findings are linked to other reports that show higher genetic burden of systemic vascular complications in individuals of African ancestry compared to individuals of European ancestry.^99–101^ Consistent with previous findings,^102^ our study also shows gender related disparity in POAG in African ancestry. A larger epidemiological and genetic study in individuals of different ancestries will clarify the role of GWAS signals identified here or other additional signals in differential risk burden between ancestries and genders, distinct from what has been reported globally.^103^

We further performed extensive in-silico validation of the *SIX6* and *CDKN2B-AS1* loci, using GTEx and EHR data across BioVU and UKBB. In our study we found evidence of significant interaction between *SIX6* rs33912345 and causal variants in chr9p21.3, with concomitant effect on expression of a primary cilia gene *CDKN2A,* and *CDKN2B* in the *CDKN2B-AS1* locus. These effects are particularly enhanced between homozygous rs33912345 and homozygous causal variants in chr9p21.3. Our results highlight a regulatory mechanism for rs33912345, which aligns to previously reported regulatory roles for this variant. For example, a Brazilian population study reported additive association effect of an independent chromosome 16 homozygous rs1362756 in *SALL1* gene in the presence of rs33912345 CC genotype, but not with heterozygous or wildtype genotypes.^104^ Furthermore, Carnes and co-authors reported that patients who have rs33912345 CC genotype have a significantly thinner retinal nerve fiber layer, (a risk factor for glaucoma) than patients homozygous with non-risk allele.^105^ This suggest that trans-effect of the rs33912345 variant in POAG risk is complex and potentially involve multiple loci and genes. In mouse model with elevated IOP, the *SIX6* missense variant was shown to increase the risk of glaucoma-associated vision loss by disrupting the development of the neural retina by upregulating the *CDKN2A* expression, leading to a reduced number of retinal ganglion cells (RGCs)^105–108^ In addition, TWAS-PheWAS analysis for the two chr9p21.3 locus genes revealed associations with circulatory and neoplasm traits. Thus, the *SIX6/CDKN2A/B* association pattern observed here potentially reflects the balance between tumor suppression, senescence (apoptosis, autophagic) which favors angiogenesis, and cell proliferation or tumorigenesis. Future analyses will clarify on the trans effect of the rs33912345 with other POAG loci in the genome in the context of glaucoma etiology.

By integrating evidence from vascular and cell proliferation mechanisms that emerged from our results, we suggest that primary cilia may potentially be a common link between vascular diseases and glaucoma pathophysiology. The novel associated genes identified in the GBMI GWAS, *CCDC13, CNTF* and *INTU,* the gene with male-specific association in Africans, *CELSR2,* and more than 40 other genes prioritized using TWAS, have been found to be linked to primary cilia, a microtubule-based cellular structure located on the surfaces of vertebrate cells.^109–111^ In addition, the other male specific associations were in *ATP10B* and *ARMC4* genes, which are cilia-related genes implicated in ocular, vascular, and neoplastic traits.^89,112–117^ Primary cilia cells are crucial in normal function of the smooth muscles and play important roles in cell proliferation mechanisms.^118^ In fact, primary cilia, through their dysfunction, contribute to cancer via interference in cancer signaling pathways, such as the Wnt signaling pathway.^118,119^ Primary cilia on vascular endothelium have been proposed to play a critical role in the regulation of vascular barrier.^118^ The vascular barrier controls the exchange of molecules and ions between blood and tissues and prevents dangerous substances from entering the tissue and causing damage.^119,120^ Primary cilia are present in the eye, i.e. retinal pigment epithelium, and they carry out sensory function through various signaling pathways.^120,121^ Loss of primary cilia is associated with several pathologies that have anomalies in these mechanisms, such as retinopathy and Leber congenital amaurosis.^122,123^

In the eye, the TM also contains primary cilia and these structures change with IOP. When the IOP is elevated, primary cilia shorten, promoting the expression of tumor necrosis factor α and transforming growth factor β (TGF-β), perhaps initiating mechanisms leading to glaucoma.^124^ Some investigations have suggested a direct link between Wnt signaling and glaucoma, since Wnt signaling is involved in the regeneration of the optic nerve after injury and also in IOP regulation.^85,125,126^ Recent studies have shown that one of the first genes linked to glaucoma, *MYOC* producing myocilin, is present in the base of primary cilia and its expression affects pathways related to ciliary signaling, such as TGF-β.^127,128^ This suggests that ciliary pathways are associated with the secretion of myocilin, of which accumulation in the TM in mutant *MYOC* causing several open-angle glaucoma subtypes.^129^ Investigating the mechanisms that influence specific primary cilia functionality will contribute to understanding the role of this structure in the pathogenesis of glaucoma, and eventually to develop novel drugs and therapies.

### Strengths and limitations of the study

In this study, there are several strong methodological points as well as limitations. The study represents the largest multi-ancestry genetic study conducted in POAG. The large sample size provided great statistical power to detect novel associations that were also replicated, with the same direction of effect, in the independent IGGC data. Furthermore, the meta-analysis of two datasets and vast resources from different biobanks afforded an opportunity to explore genetic interactions between likely causal variants of glaucoma, and to conduct in silico investigation of the potential shared biology between POAG, cancer, vascular traits and POAG.

Whereas our sample size is larger compared with other POAG studies, several issues that are inherent in a biobank collaborative context like GBMI might lead to a conservative signal detection. POAG phenotyping methods in GBMI were different across the biobanks. In addition, most of the phenotyping was for glaucoma and not specifically POAG, potentially introducing biological heterogeneity. Even though this is not a major issue in biobanks with a majority of European and African ancestry subjects where POAG is the predominant glaucoma subtype, the signals detected from Asians will also probably be from primary angle-closure glaucoma, where this subtype predominates.^6,30–32^ In addition, signals from other minor glaucoma subtypes (such as exfoliation glaucoma) might also introduce noise. Moreover, despite using a powerful approach to mitigate the effect of unbalanced case-control sampling inherent in a biobank context, the attenuating effect of inappropriate controls will persist, making our signal detection conservative.

Furthermore, despite the fact that this analysis used multi-ancestry meta-analysis, the subjects are still predominantly of European ancestry. We believe that recruitment of cohorts that include ethnic minorities will improve knowledge transferability and health equity.

Finally, even though we did extensive in-silico validation using available data and gleaned broad biological pathways that might be implicated in POAG, we did not perform functional validation of the novel genes identified in this study. Therefore, further in vitro studies using relevant human tissues and in vivo using model organisms are still necessary to define the biological role of these genes in POAG and potential link to vascular and neoplastic traits.

### Conclusion and future directions

Our study, the largest and most diverse GWAS in POAG up to date, identified ten ancestry-specific loci, five of which were associated with POAG in either male or female only. A larger genetic study in individuals of diverse ancestries will clarify the role of GWAS signals identified here or other additional signals in differential risk burden between different continental populations, and males and females for this trait. Moreover, there is a need for further studies to ascertain the magnitude of inter-loci interactions and the roles of the *SIX6* missense variant in the overall interaction landscape. Further, the role of these interactions in difference in burden for POAG and vascular/cancer related traits between the ancestries need to be elucidated. In summary, by integrating evidence from vascular and cell proliferation mechanisms that emerged from our results, we suggest that primary cilia may potentially be involved in glaucoma pathophysiology. Further investigations are needed to elucidate their role in glaucoma.

## Data Availability

All data produced in the present study are available upon reasonable request to the authors

## Acknowledgements

We would like to acknowledge Melinda Aldrich for critical review on an earlier version of the manuscript. We would like to acknowledge the organizing committee of the International Common Disease Alliance for intellectual contributions on the set up of the GBMI as a nascent activity to the larger effort. We also thank Daniel King from the Hail team and Sam Bryant from the Stanley Center Data Management team for helping with the Google bucket set up and data sharing, and Sinead Chapman for helping in paper submission.

Eric R. Gamazon is supported by the National Institutes of Health (NIH) Awards R35HG010718, R01HG011138, R01GM140287, and NIH/NIA AG068026. Valeria Lo Faro was supported by the European Union’s Horizon 2020 research and innovation programme under the Marie Skłodowska-Curie grant agreement No.675033 (EGRET plus). Additional funding was provided by the Rotterdamse Stichting Blindenbelangen (grant ID B20150036).

Karen Joos was supported by the Joseph Ellis Family and William Black Research Funds, and an Unrestricted Departmental Grant to the Vanderbilt Eye Institute from Research to Prevent Blindness, Inc., NY.

We acknowledge all the participants in the study and biobanks that were involved in generating POAG summary data: BioBank Japan (Yukinori Okada, Koichi Matsua, and Masahiro Kanai), BioMe (Ruth Loos, Judy Cho, Eimear Kenny, Michael Preuss, and Simon Lee), BioVU (Nancy Cox and Jibril Hirbo), Colorado Center for Personalized Medicine (Kathleen Barnes, Michelle Daya, and Chris Gignoux), deCODE Genetics (Kári Stefánsson and Unnur Þorsteinsdóttir), Estonian Biobank (Andres Metspalu, Reedik Mägi, Tõnu Esko, and Priit Palta), FinnGen (Aarno Palotie, Mark Daly, Samuli Ripatti, Mitja Kurki, and Juha Karjalainen), Generation Scotland (Caroline Hayward and Riccardo Marioni), HUNT (Kristian Hveem, Cristen Willer, and Sarah Graham, Ben Brumpton, and Brooke Wolford), Lifelines (Esteban Lopera, Serena Sanna, Harold Snieder), Michigan Genomics Initiative (Sebastian Zoellner, Michael Boehnke, Lars Fritsche, and Anita Pandit), Taiwan Biobank (Yen-Feng Lin, Yen-Chen Feng, and Hailiang Huang), and UK Biobank (Konrad Karczewski and Alicia Martin).

## Author contributions

Study design: V.L.F, W.Z, N.J.C, J.H.

Data collection/contribution: V.L.F, S.S., N.J.C., J.H., Y.W., E.L., K.L., M.K., Y.O., P.S., P.P., A.R.M., N.M.J., GBMI

Data analysis: V.L.F, A.B., W.Z, D.Z., P.S, J.H.

Writing: V.L.F, J.H

Revision: V.L.F., A.B., W.Z., D.Z., Y.W., K.L., M.K., E.L., P.S., P.P., R.T., X.Z., S.N., S.S., I.M.N., H.S., I.S., A.R.M., M.A.B., C.W., S.M., N.I, E.R.G., N.M.J., K.J., N.J.C., J.H.

## Declaration of interest

Eric R. Gamazon receives an honorarium from the journal Circulation Research of the American Heart Association as a member of the Editorial Board.

SM is a co-founder and holds stock in Seonix Pty Ltd.

## STAR Methods

### Association testing

The overall design of this study is reported in Figure 1. As part of the GBMI, a large-scale multi-ancestry meta-analysis of genome-wide association studies was conducted including a total of 26,848 cases and 1,460,599 controls from 15 global biobanks across six ancestries (Suppl. Table S1, Suppl. Figure S1-2). Phenotype definition, biobank specific quality control and standardized GWAS was performed by each contributing biobank. POAG phenotyping was done using any of the three approaches: phecode mapping (BioVU, UKBB, HUNT, MGI and CCPM), ICD9/ICD10 codes (BioMe, FinnGen, EstBB, MGB, deCODE) and either one or a combination of physicians’ diagnosis, glaucoma medications or self-reporting (BBJ, TWB, Lifelines, GS and QSkin) (Suppl. Table S1).^130^ Potential effect of heterogeneity of phenotyping was checked by comparing a subset of manually reviewed and phecode defined patients in BioVU with an independent traditionally phenotyped cohort (Supplementary information, Suppl. Table S2).

Meta-analysis was performed followed by the variant-level quality control for each biobank by flagging markers with different allele frequencies compared to gnomAD and excluding markers with imputation quality score < 0.3 (Suppl. Table S3).^26,35^ Gender-specific association analysis was also conducted across 9 biobanks in 7916 cases and 269,105 controls (males) and across 10 biobanks in 9538 cases and 342,870 controls (females) (Suppl. Table S4-5, Suppl. Figure S3-4). The number of independent loci and number of top hits for each biobank contributing to GBMI are reported in Suppl. Figure S5.

To increase the power to discover additional variants associated with POAG, a meta-analysis was performed combining GBMI (n=1,259,040), International Glaucoma Genetics Consortium (IGGC) of European ancestry (n=192,702) and Genetics of Glaucoma in People of African Descent (GGLAD) (n=26,295) summary statistics, excluding any biobank cohorts from GBMI that overlap with the two other datasets in our analysis (FinGenn, BioME and UKBB Africans). This represents the largest and most diverse GWAS in POAG up to date (number of cases=46,325, number of controls=1,431,712) (Suppl. Table S7, Suppl. Figure S9-10).^24,131^ Fixed-effect meta-analyses based on inverse-variance weighting were performed. We further performed ancestry specific meta-analysis of the data. We defined genome-wide significant loci by iteratively spanning the ± 500 kb region around the most significant variant and merging overlapping regions until no genome-wide significant variants were detected within ± 500 kb. The most significant variant in each locus was selected as the index variant. To identify independent variants, clumping was performed in PLINK ( http://pngu.mgh.harvard.edu/purcell/plink/) using r^2^ < 0.05 as linkage disequilibrium (LD) metric threshold, physical distance of 500 kb, 1000genomes Europeans as LD reference panel and the significance threshold for index SNPs was set to P<1×10^-5^.^132^ Quantile-quantile (Q-Q) and Manhattan plots were generated to visualize the results.

POAG prevalence in European and African American were estimated in 3,414,079 BioVU patients that are ≥40 years older and self-identify as European Americans (EA, n=1,218,124, 555,132 males 662,825 females) or African Americans (AA, n=154,273, 66,691 males 87,541 females). Comorbidity with circulatory related phecodes (phecodes=394-459) in a total of 1,968,903 individuals with ≥2 instances of mention of the relevant phecode excluding individuals with any mention of eye phecodes (phecodes=360-379) (AA, n= 273379, 1549 cases 271830 controls, and EA, n= 1695524, 5446 cases 1,690,078 controls) was inferred by performing logistic regression analysis conditioned on gender and age as covariates. Odd ratio comparison was done using epitools in R.

### SNPs and gene annotations

Polymorphisms associated at a genome-wide significant level (p<5e-8) LD clumped as above (http://pngu.mgh.harvard.edu/purcell/plink/).^132^ Significant polymorphisms were annotated with the gene inside whose transcript-coding region they are located, or alternatively, the nearest gene. In addition, the polymorphic sites were functionally annotated using ANNOVAR.^133^ Exonic SNPs (single nucleotide polymorphism) were investigated further using SNPnexus to uncover non-synonymous variants.^134^ The possible damaging effects of non-synonymous SNPs on protein structure and function were predicted using the Sorting Intolerant From Tolerant (SIFT) and Polymorphism Phenotyping (PolyPhen) scoring tools.^135,136^

### Enrichment analysis

To identify the functional roles and tissue specificity of the associated variants, we performed gene prioritization and tissue- and gene-set enrichment analyses using DEPICT (Suppl. Table S8-10) in which we prioritized POAG-associated genes using a co-regulation-based method across multiple different tissues.^137^ The tool assesses the potential role of genes independent of the presence of an eQTL, making it possible to test more genes.

For the gene-set enrichment analysis, gene expression data from 77,840 samples was used to predict gene function for all genes in the genome based on similarities in gene expression. In DEPICT, the probability of a gene being a member of a gene set was estimated based on their co-functionality to prioritize the most likely causal genes. A total of 14,461 reconstituted gene sets was generated which represent a set of biological annotations: Gene Ontology (GO) gene sets, REACTOME gene sets, Kyoto Encyclopedia of Genes and Genomes (KEGG) gene sets, InWeb protein-protein interactions, and Mouse Genetics Initiative gene sets (MP). Bonferroni correction was applied for multiple comparisons of 14,461 independent tests (p <0.05/14,461).

For tissue enrichment, microarray data from 37,427 human tissues of 209 Medical Subject Heading (MeSH) annotations from Affymetrix HGU133a2.0 platform microarrays was used to identify genes with high expression in different cells and tissues.

### TWAS and fine-mapping analyses

We used three gene-based methods, PrediXcan, joint tissue imputation (JTI) and unified test for molecular signatures (UTMOST) for correlating the genetic component of gene expression with phenotype.^69–71^ PrediXcan estimates gene expression weights by training a linear prediction model in a reference sample with both gene expression and SNP genotype data.^138^ UTMOST and JTI methods borrow information across transcriptomes of different tissues, leveraging shared genetic regulation, to improve prediction performance in a tissue-dependent manner.^71^

There is no ocular tissue in GTEx data, so we used proxy tissues that have biological functions that we inferred to be crucial in visual perception. These include: vascular tissues (artery and heart tissues) that are crucial in production and drainage of aqueous humor (defects in this system can cause glaucoma), tissues of the nervous systems, which the eye is considered its extension, and the liver tissue because it is a pivotal metabolic center.^139–141^ We consider the brain cortex as the most relevant GTEx tissue since the visual cortex is located there.

For the three models, gene expression prediction models were trained for 23 different human nervous, vascular and liver tissues (Suppl. Table S10-11) using GTEx v8 data.^142^ The corresponding genotype data were imputed using the University of Michigan Imputation Server, with 1000Genomes (phase 3 ver. 5) as a reference panel.^143,144^ Models with non-zero weights that met set significance criteria (FDR<0.05 from the cross-validation in each tissue) were retained in the database. For each model, there was also a corresponding file with covariance data for the SNPs in each model. The three models were applied to the GBMI-IGGC-GGLAD GWAS meta-analysis summary results (n=1,478,037: 46,325 cases, 1,431,712 controls) (Suppl. Table S10-11).

Fine-mapping of TWAS association signals was done using FOCUS, a probabilistic gene-level fine-mapping method, to define credible sets of genes that explain the expression-trait signal at any given locus.^145^ The default non-informative priors implemented in FOCUS were used to estimate the posterior inclusion probability and a 90% credible set of genes at a given locus.

### Polygenic risk scores (PRS) and pleiotropy

PRS for POAG were constructed from the leave-biobank-out GBMI-IGGC-GGLAD meta-analysis summary statistics in six different biobanks, BioVU (n=85,615), UK Biobank (n=370,088), Biobank Japan, BBJ (n=178,726), Lifelines (n=14,930), Estonian Biobank (n=53,821), and glaucoma cohort GLGS^146^ (n=3,739), with PRS-CS-auto option, and the best performing European reference panel (based on R2) of either 1KG Phase 3 or UK Biobank was used to estimate LD.^34^ In the target samples, genotypes were filtered for SNPs using the following criteria: minor allele frequency< 0.01, missing genotype rate<0.05, filters out variants which have Hardy-Weinberg equilibrium exact test p-value < 1e-6, and exclude individuals with missing data < 0.1. Only unrelated individuals with genetic relatedness less than 0.05 were retained. We used EHR/or self-reported health conditions (depending on the data availability in each biobank) in a total of 617,565 individuals across five biobanks (excluding GLGS) to explore pleiotropy of genetic risk for POAG. We then evaluated the predictive performance of the PRSs generated using leave-biobank-out meta-analysis results in the five biobanks. PRSs were then tested for association with 17 disease categories using a phenome-wide association study (PheWAS).

### Genetic correlation analysis

To evaluate genetic correlations between POAG and vascular traits, cross-trait LD score regression analysis was performed using LDSC software ^147^ with GBMI-IGGC-GGLAD meta-analysis summary statistics and, 13 well powered published summary statistics for vascular and neoplastic traits, respectively, obtained from GWAS catalog.^148^ For this we used default LD scores estimated for the European ancestry populations.^147^

### Expression effects of missense variants (male-specific rs74113753 & trans-ancestry rs33912345)

Variants in *SIX6* and chr9p21.3 *CDKN2B-AS1* loci have been associated with POAG ^149–156^ and related risk factors and endophenotypes, such as peripapillary retinal nerve fiber layer,^157^ optic nerve degeneration,^153,158^ and vertical cup-disc ratio (VCDR)^159–165^ across all ancestries. The rs33912345 missense variant (p.His141Asn) is the lead variant in *SIX6* locus, and is outside the DNA binding site and is speculated to affect the ability of the *SIX6* gene to interact with other transcription factors and cofactors.^108^ rs33912345t is also a GTEx eQTL for several genes within the *SIX6* locus.^142^ The *SIX6* locus and the *CDKN2B-AS1* locus on chr9p21.3 are known to be associated with POAG^149–156^ and cardiovascular disease^166^. Studies in both glaucoma models and cell lines indicated that *SIX6* missense variants interact with genes in the chr9p21.3 locus.^107,108^ However, no comprehensive analysis of this interaction has been done in human genetic and transcriptomic data.

The effect of the missense rs33912345 variant on expression patterns of genes *SIX6* and *SALRNA1* in the *SIX6* locus and on genes *CDKN2A* and *CDKN2B* in the *CDKN2B-AS1* locus were determined by performing regression analysis of residuals of the GTEx normalized gene expression levels. For each tissue, the possible confounders (gender, platform, the first five principal components, and Probabilistic Estimation of Expression Residuals (PEER) factors)^71^ have been accounted for in the analysis. Since the missense variant did not pass the QC filter, we used a proxy SNP, rs7493429 which was in high LD with the missense variant (r^2^=0.724, D’=0.99) determined using R package LDlinkR (https://github.com/CBIIT/LDlinkR).^167^ We also evaluated the effect of African specific missense variant rs74113753 using proxy rs17641032 variant (-862bp, r^2^=0.699, D’=1, 1000 genomes)^167^ on measured gene expression residuals for a total of 24 genes in a 500 kb window either side of the variant (*HENMT1-AMPD2*) that passed quality filters in GTEx muscle skeletal tissues (n=706) in all the samples and sex stratified set (males=469, females=237).

We further evaluated the effect of the variant rs33912345 in the *SIX6* locus on trait association to variants in *CDKN2B-AS1* locus in four steps. We first checked the differences in the gene expressions of *SIX6* and *SALRNA1* measured in skeletal muscle tissues (one of the only tissues with GTEx expression data remaining after correcting for confounders) between the haplotype background that has the reference and derived allele. Secondly, we rebuilt the muscle skeletal genes expression model while excluding any variants that were in LD with the rs33912345 missense variant (r^2^>0.1). The PrediXcan and UTMOST muscle tissue models were the best performing gene models for *SIX6* (r^2^=0.131, compared to JTI r^2^=0.120) and *SALRNA1* (r^2^=0.024 compared to PrediXcan r^2^=0.016), respectively. Thirdly, we evaluated the Pearson correlations in gene expressions between genes in the *SIX6* and *CDKN2B-AS1* loci. We then checked for statistical interaction between the *SIX6* sentinel SNP and two genome-wide significant variants from the *CDKN2B-AS1* locus with traits that represent groups in LD with glaucoma association in the loci. The chr9p21.3 POAG associated GWAS variants are in LD in European ancestry individuals (r^2^>0.1) with more than 140 GWAS catalog variants in the ∼260 kb cluster associated with vascular^166^, carcinoma, and neurological traits.^148^ We used two variants in the locus that are associated with a whole range of the cardiovascular and cancer related traits; rs2891168 (coronary artery disease, myocardial infarction, and beta blocking agent use measurement),^50,168–172^ and rs10811650 (breast cancer, melanoma, and hair color).^117,173–175^ We finally determined if these interactions had effects on expression patterns of *CDKN2A & CDKN2B* in GTEx data brain cortex tissue. Mean expression difference between allele combinations was assessed using Student’s t-test and ANOVA (Suppl. Table S20).

To determine phenotypic consequences of the *SIX-CDKN2B-AS1* loci interactions, and African male-specific missense variant, we further performed TWAS-PheWAS for the two chr9p21.3 genes and *CELSR2* (that have male-specific expression association with the missense variant) gene in summary statistics from the UKBB (n=396,618) and BioVU (n=59,805), followed by meta-analysis of the two PheWAS (n=456,423) across 731 traits and diseases grouped into 17 categories (Suppl. Table S21).^176,177^

## Important Links

https://github.com/gamazonlab/MR-JTI

https://ftp.1000genomes.ebi.ac.uk/vol1/ftp/phase3/

https://ldlink.nci.nih.gov/?tab=home

https://gnomad.broadinstitute.org/

https://www.ebi.ac.uk/gwas/

https://gtexportal.org/home/

## Supplemental Information titles and legends

**Supplementary Figure S1:**
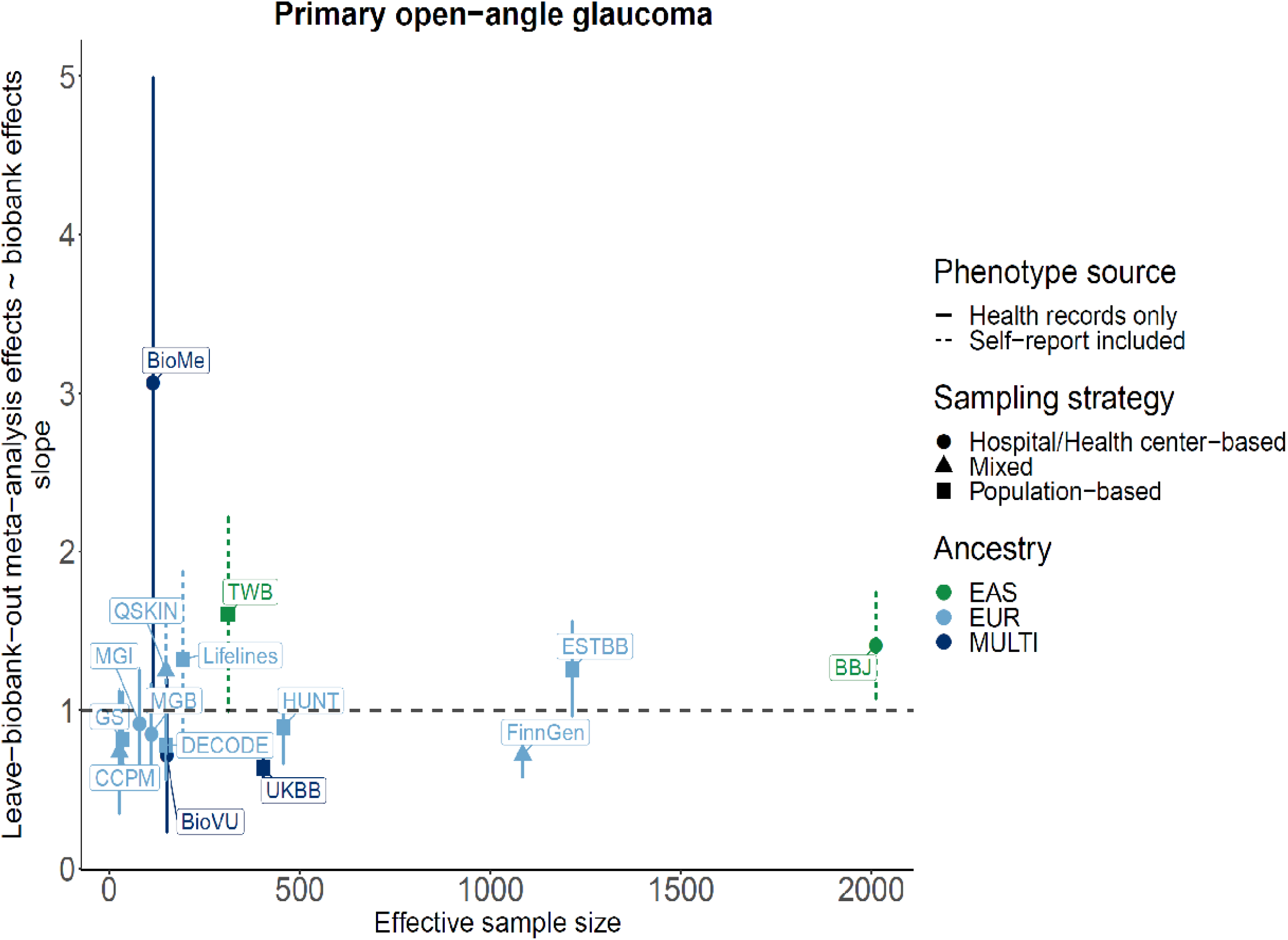
Leave-biobank-out meta-analysis vs biobank effects of the index variants for loci identified in this study. In each case index variants with strong signal way above the genome-wide threshold all-biobank meta-analysis (p-value < 10^-10) for the regression.

**Supplementary Figure S2:**
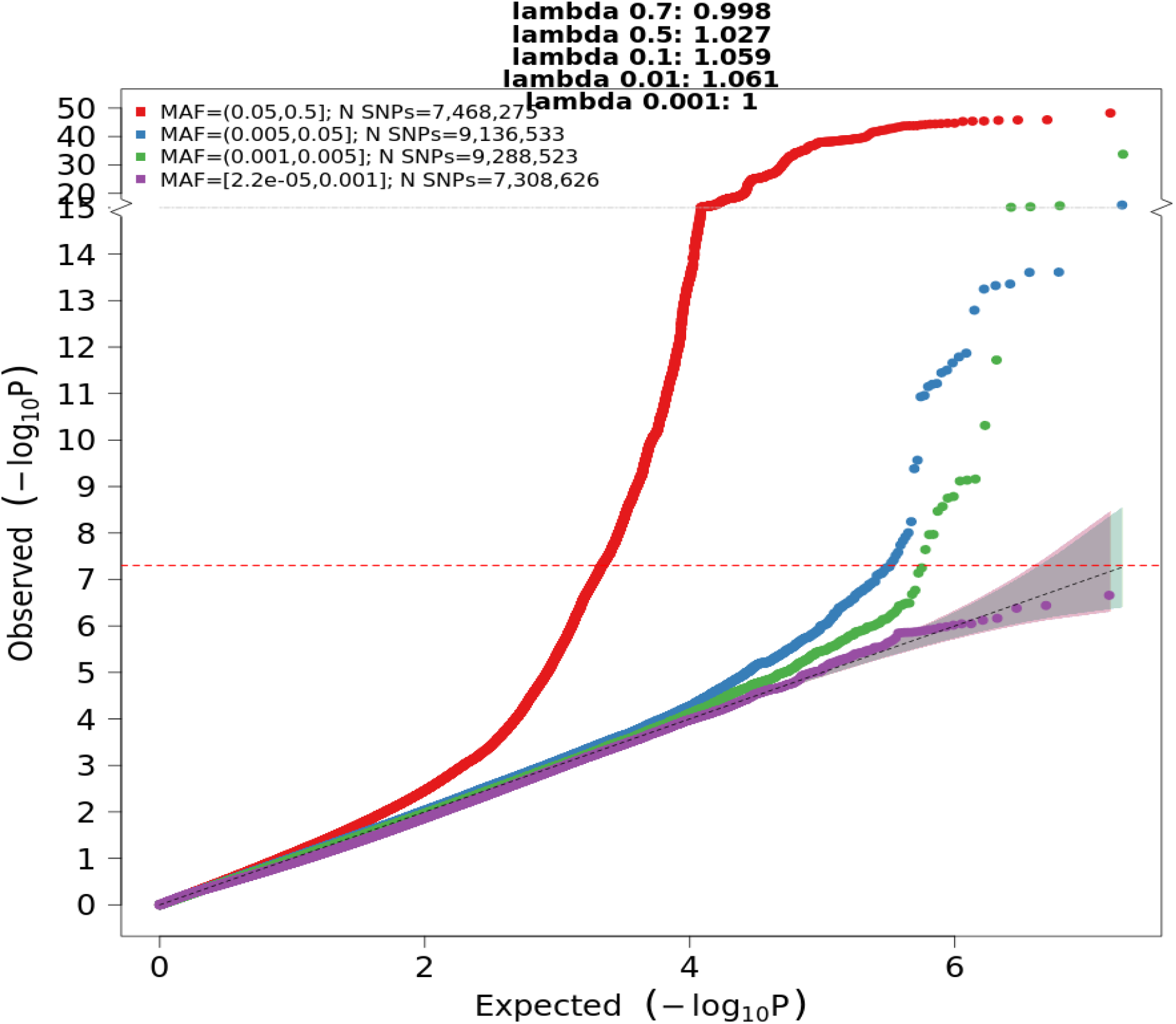
QQ plot for the GBMI POAG meta-analysis of all individuals. The meta-analysis included a total of 26,848 cases and 1,460,599 controls from 15 global biobanks across six ancestries.

**Supplementary Figure S3:**
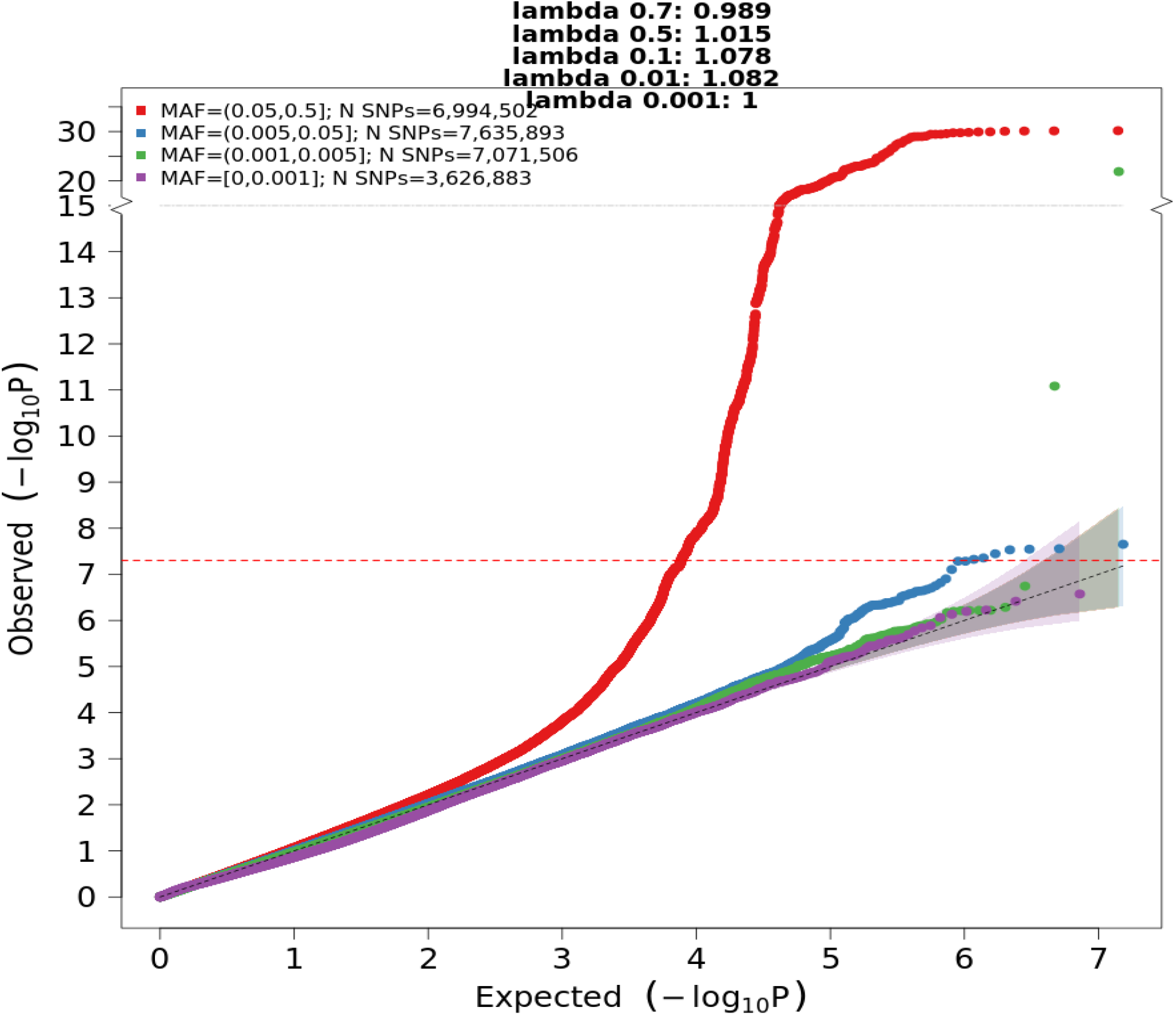
QQ plot for the GBMI POAG meta-analysis of female individuals. The meta-analysis was across 9 biobanks in 7,916 cases and 269,105 controls.

**Supplementary Figure S4:**
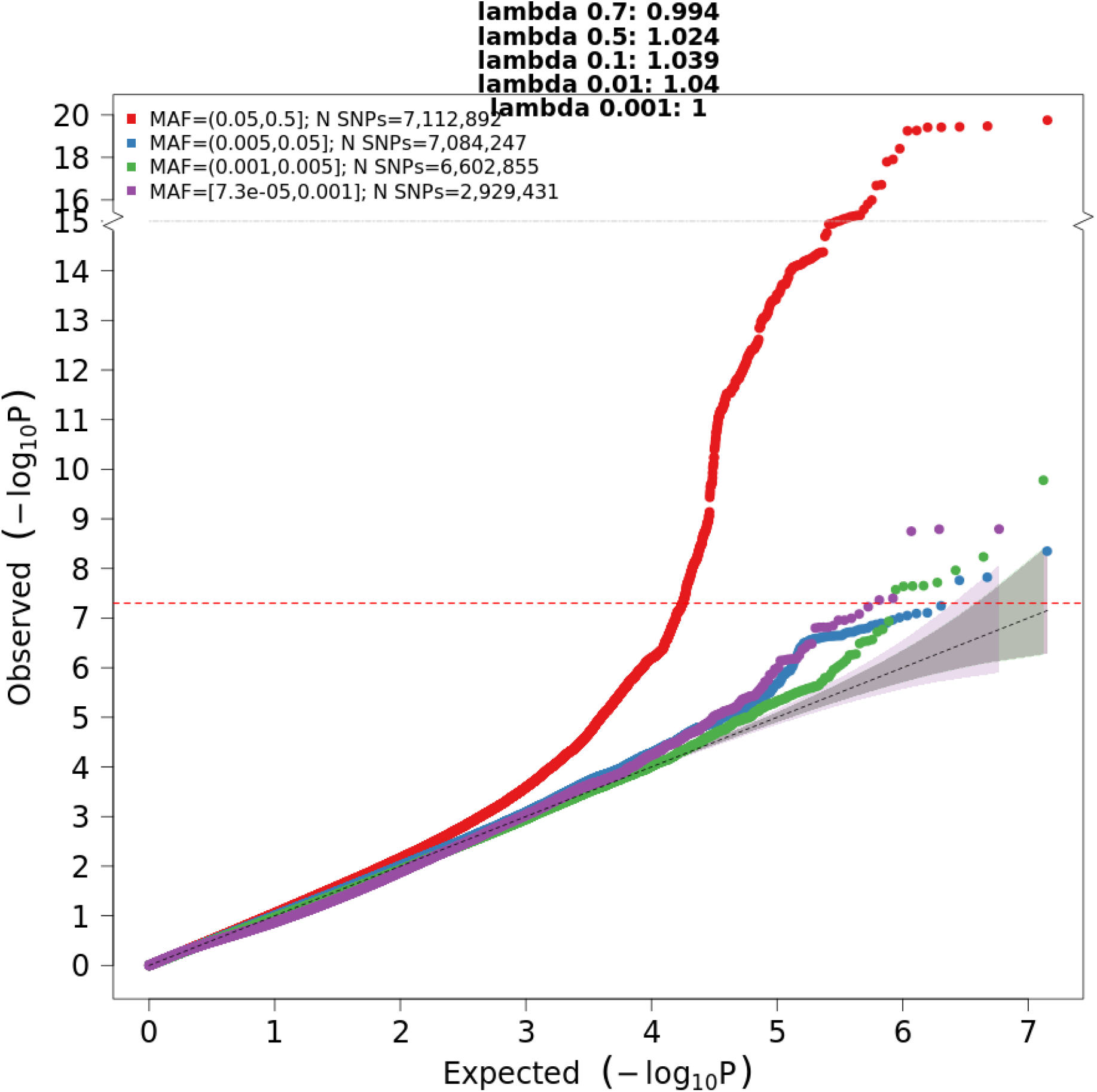
QQ plot for the GBMI POAG meta-analysis of male individuals. The meta-analysis was across 10 biobanks in 9,538 cases and 342,870 controls.

**Supplementary Figure S5:**
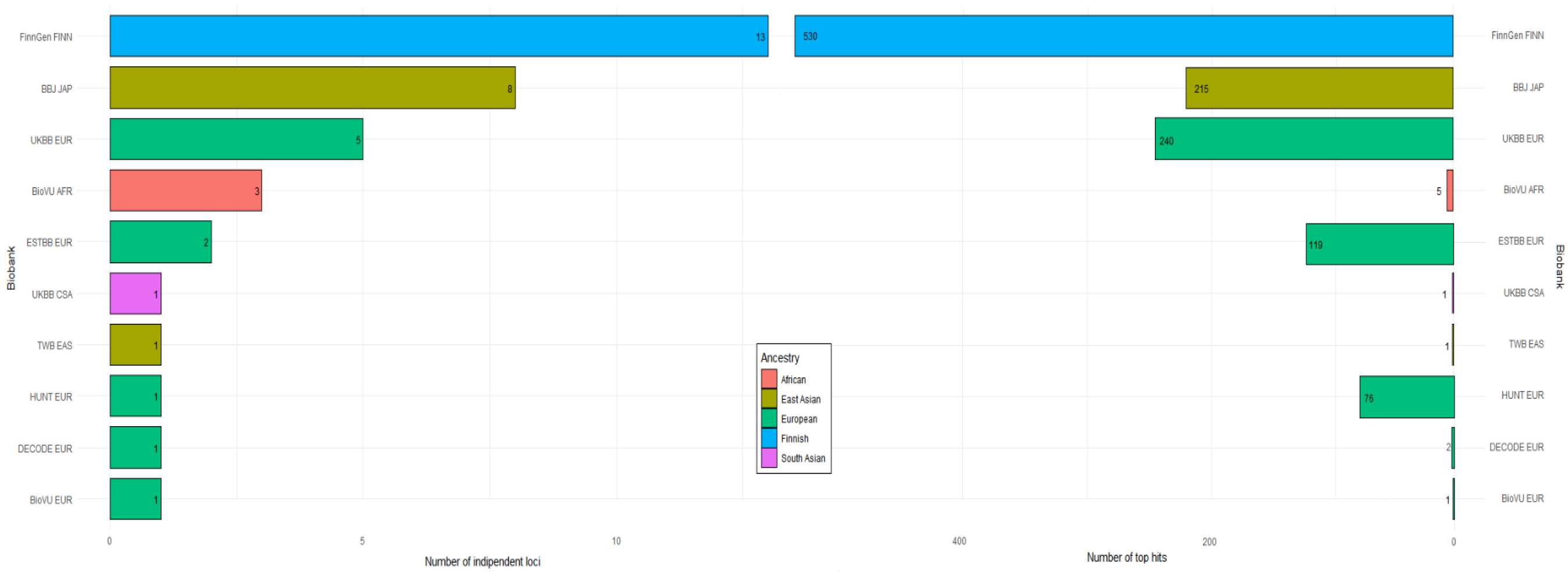
The number of independent POAG GWAS hits (p<5e-8) and loci for each biobank contributing to GBMI.

**Supplementary Figure S6:**
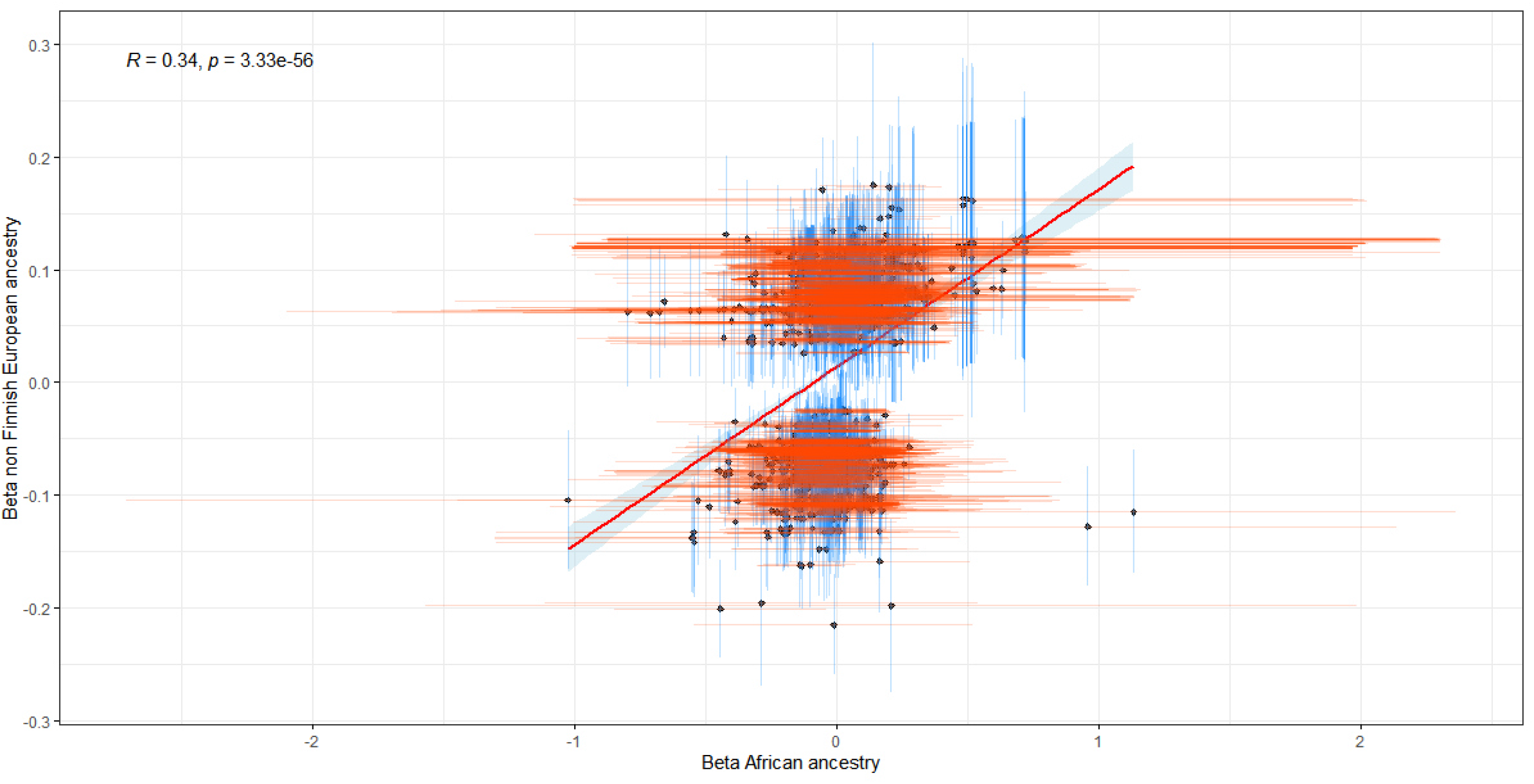
Correlation of SNP effect size estimates between non-Finnish European and African ancestry in GBMI dataset. The x-axis shows effect estimates in beta scale for the independent genome-wide significant loci obtained from the European GBMI for POAG. The y-axis shows the effect estimates in beta scale for the same SNPs obtained from the African GBMI dataset.

**Supplementary Figure S7:**
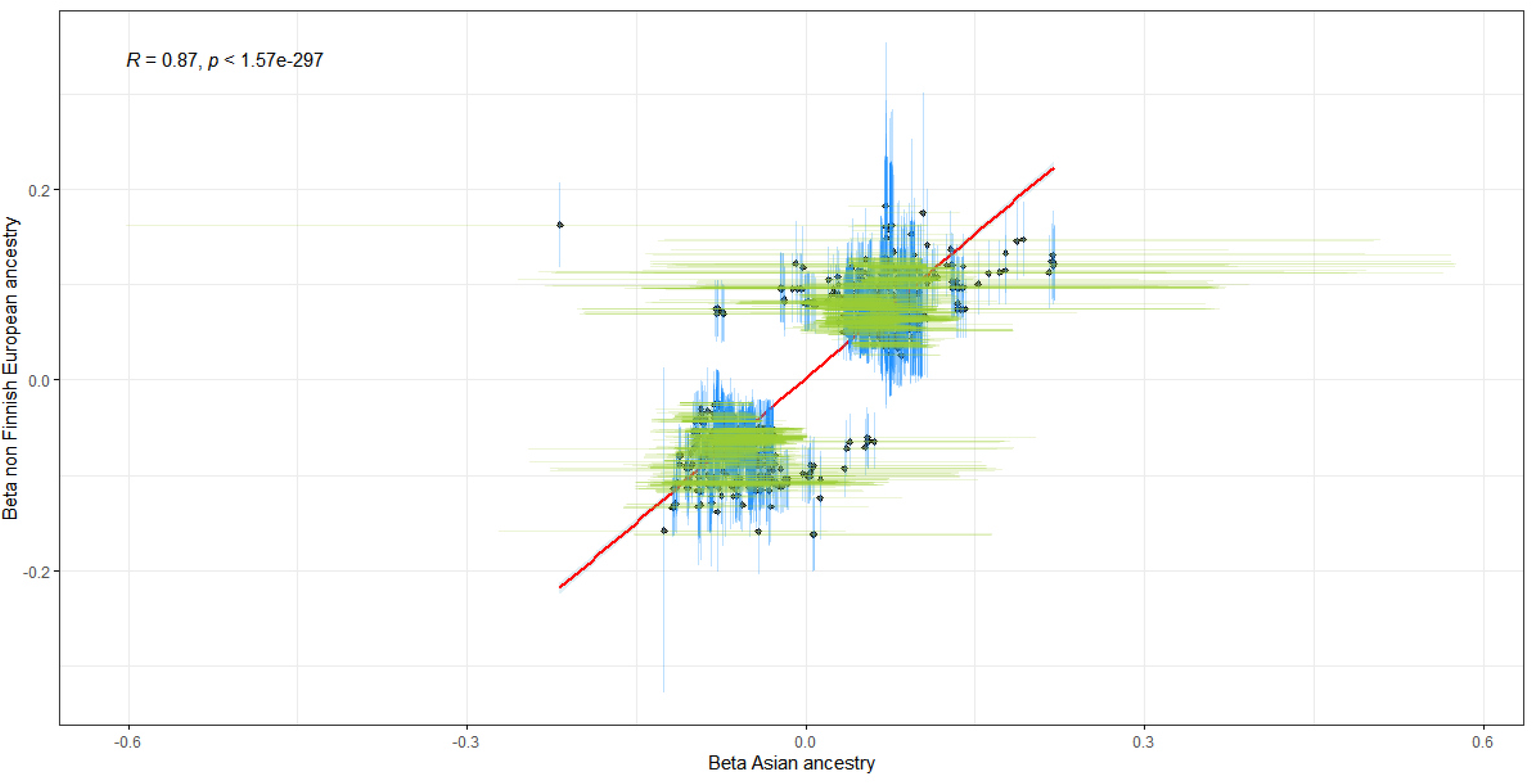
Correlation of SNP effect size estimates between non-Finnish European and Asian ancestry in GBMI dataset. The x-axis shows effect estimates in beta scale for the independent genome-wide significant loci obtained from the European GBMI for POAG. The y-axis shows the effect estimates in beta scale for the same SNPs obtained from Asian GBMI dataset.

**Supplementary Figure S8:**
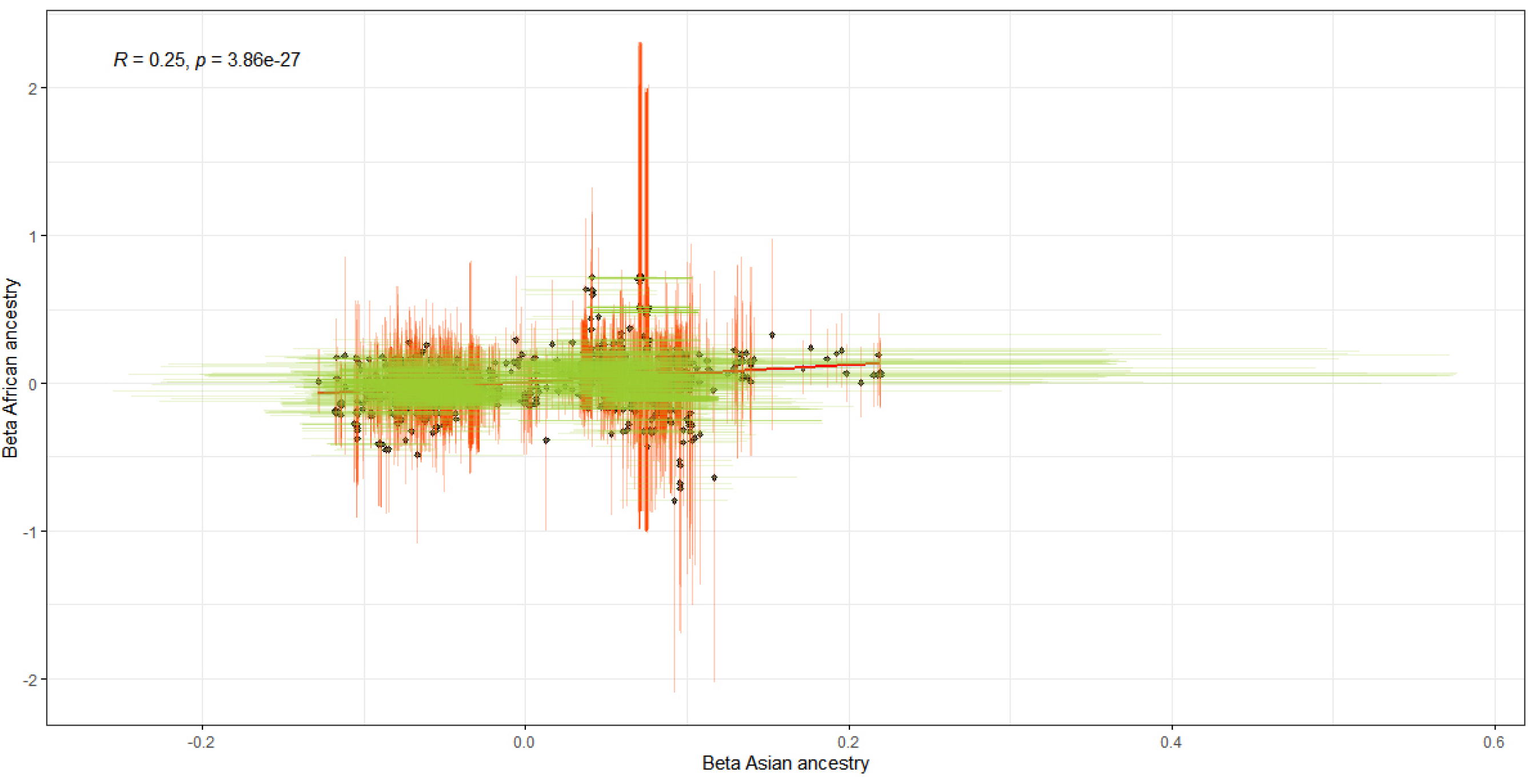
Correlation of SNP effect size estimates between African and Asian ancestry in GBMI dataset. The x-axis shows effect estimates in beta scale for the independent genome-wide significant loci obtained from the African GBMI for POAG. The y-axis shows the effect estimates in beta scale for the same SNPs obtained from Asian GBMI dataset.

**Supplementary Figure S9:**
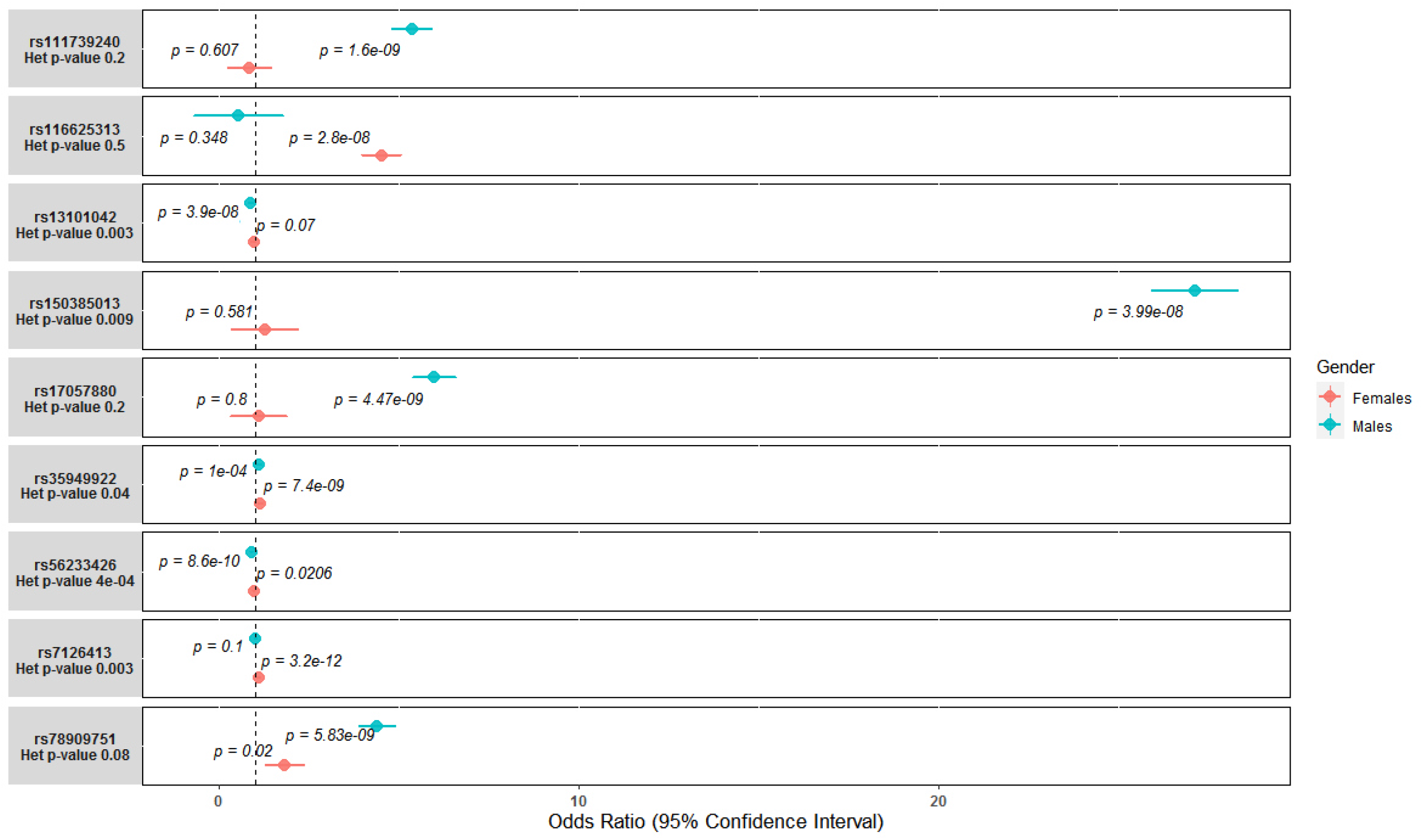
Sex-specific loci and significant loci in the combined both sex meta-analysis that show differential signal between the two sexes. -*p*= p-values; -Het p-value= Heterogeneity test p-value.

**Supplementary Figure S10:**
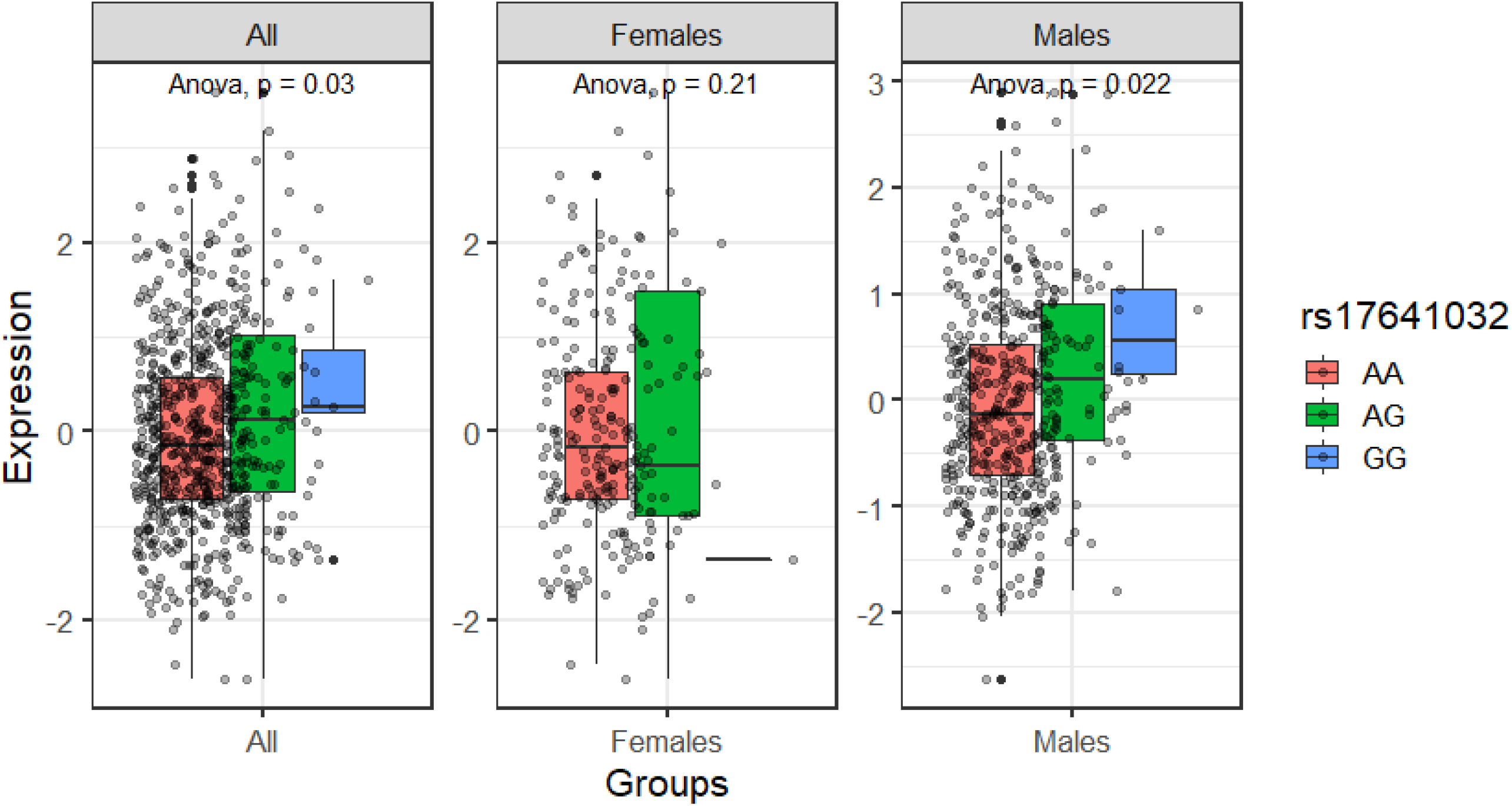
Sex difference in effect of proxy rs1741032 variant in expression of CELSR2 gene in GTEx muscle skeletal. p-values (p) are based on ANOVA tests.

**Supplementary Figure S11:**
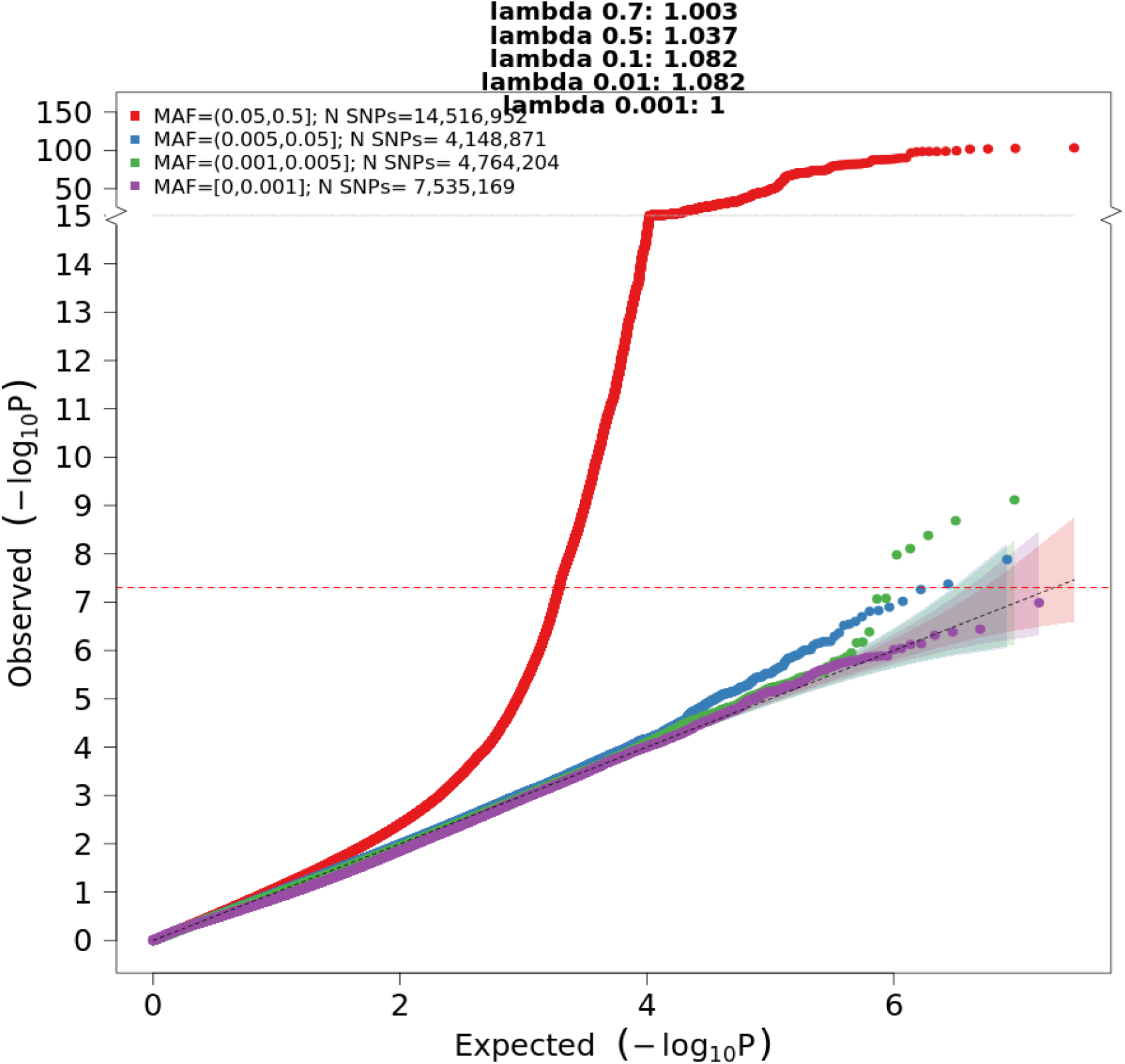
QQ plot for the GBMI-IGGC-GGLAD POAG meta-analysis. This was meta-analysis that include GBMI (n=1,259,040), International Glaucoma Genetics Consortium (IGGC) of European ancestry (n=192,702) and Genetics of Glaucoma in People of African Descent (GGLAD) (n=26,295) datasets.

**Supplementary Figure S12:**
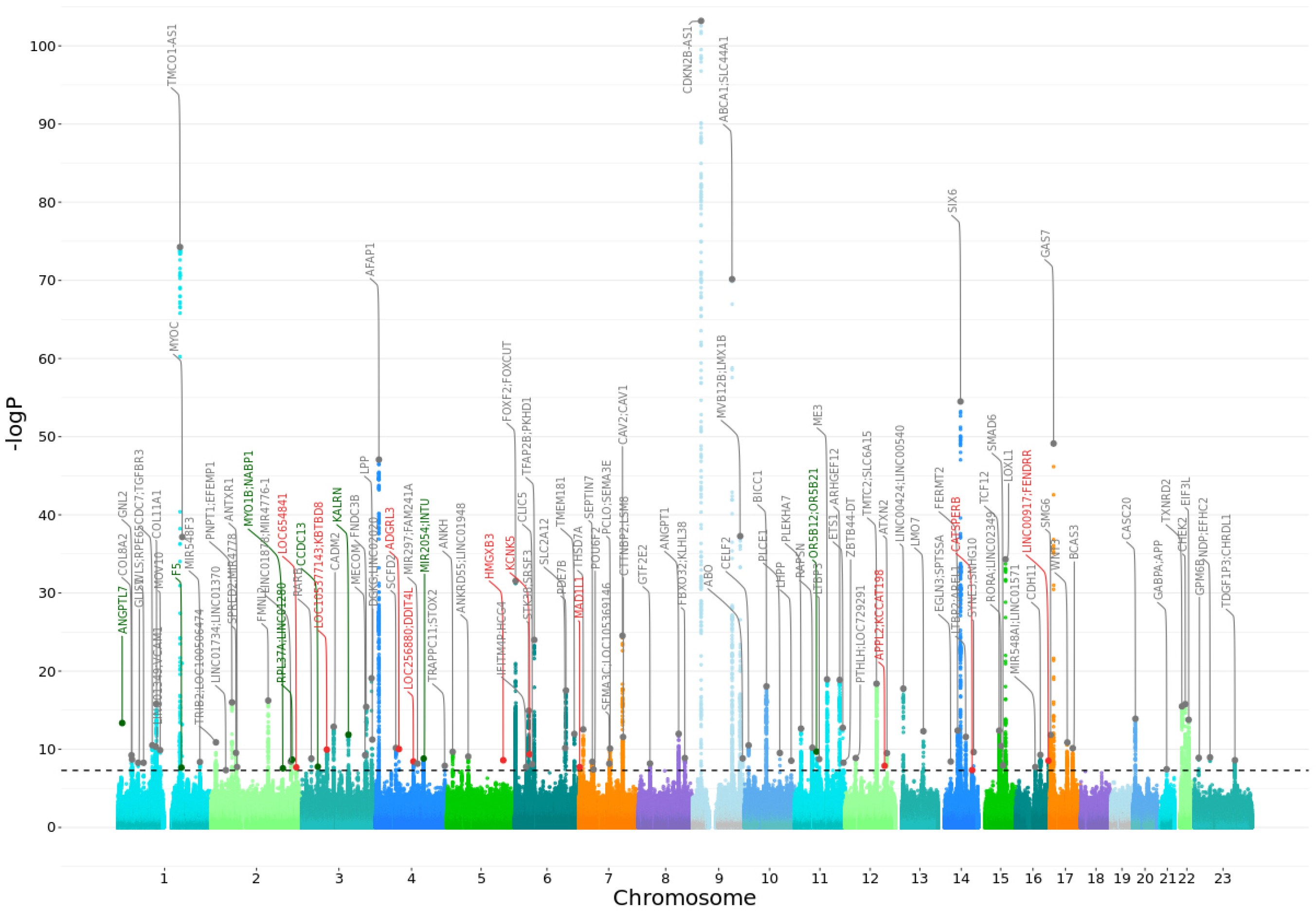
Manhattan plots for the cross-ancestry GBMI-IGGC-GGLAD POAG meta-analysis. Each dot represents a SNP, the x-axis shows the chromosomes where each SNP is located, and the y-axis shows −log10 P-value of the association of each SNP with POAG across-ancestry meta-GBMI. The nearest gene to the most significant SNP in each locus is reported. Novel loci are labeled in red and those identified in GBMI but fall below genome-wide threshold in GBMI-IGGC-GGLAD meta-analysis are labeled in green. The red horizontal line shows the genome-wide significant threshold (P-value = 5e-8; −log10 P-value = 7.30). Details of each genome-wide significant hit are in Suppl. Table S7.

**Supplementary Figure S13:**
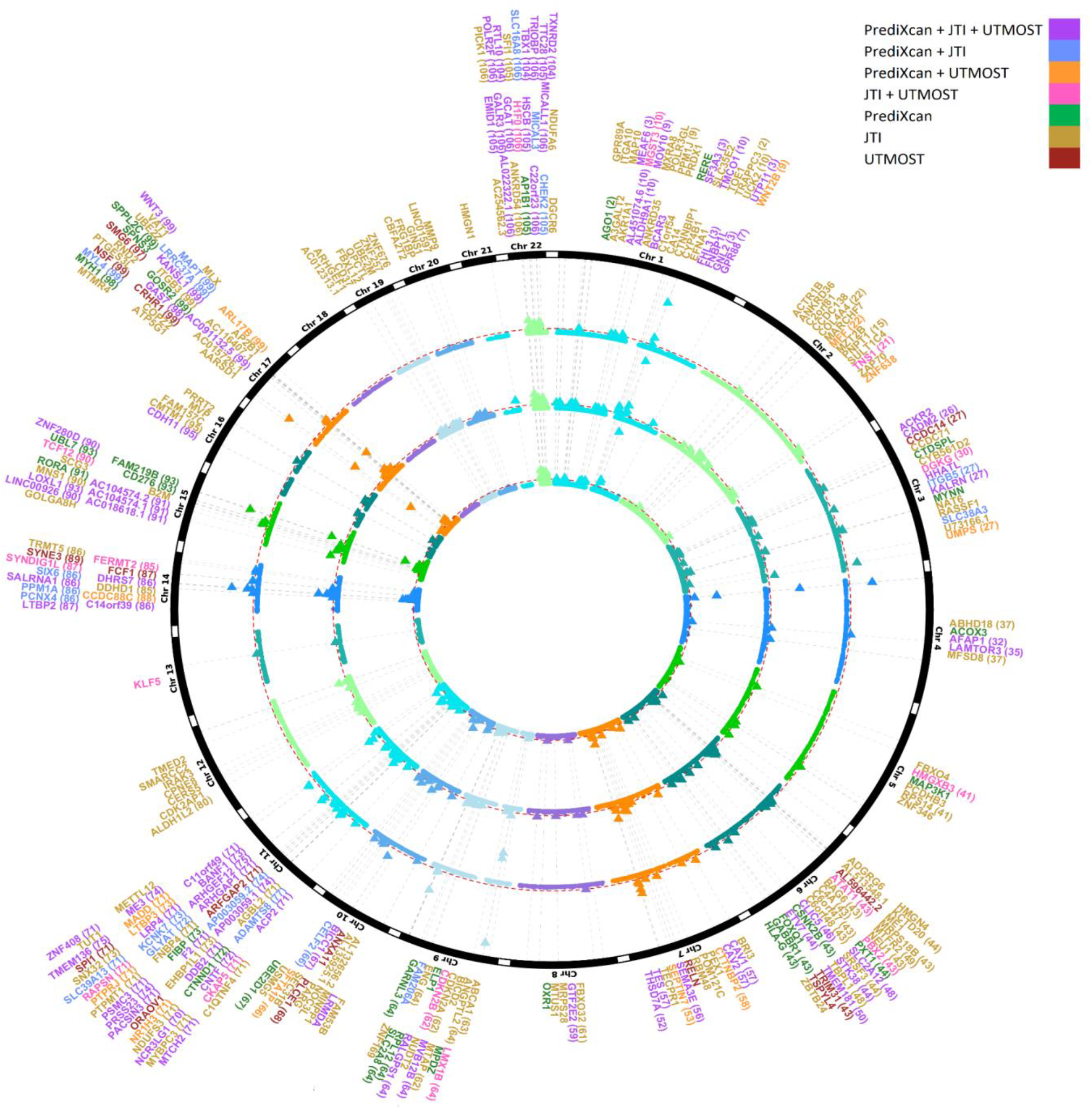
A circular-Manhattan plot showing the POAG TWAS significant gene signals (threshold p value < 2.5e-6, marked with a dashed red line in each ring) identified by the PrediXcan, JTI, and UTMOST models in 23 GTX tissues. The circles from the innermost to outermost represent: the Manhattan plot of PrediXcan, JTI, UTMOST, chromosome numbers (each one with different color) and genes identified by the models. Numbers in parentheses correspond with locus#index in Suppl. Table S7. Each genome-wide significant gene signal is represented as a colored triangle. More details are reported in Supplementary Table S12 and S13.

**Supplementary Figure S14:**
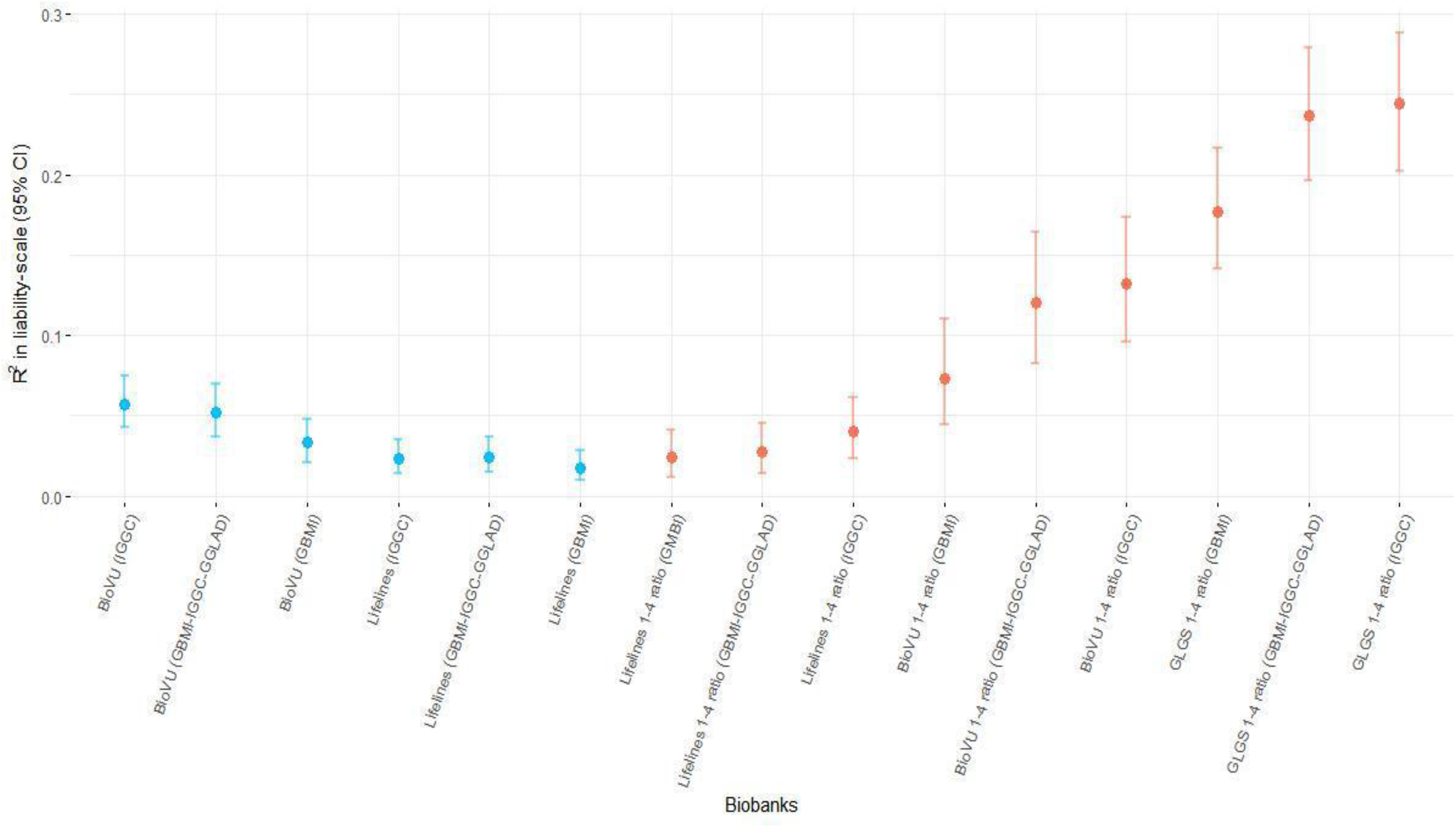
Prediction performance of the leave-biobank-out GBMI, GBMI-IGGC-GGLAD, and IGGC POAG meta-analysis as source GWAS. The prediction performance was estimated by R2 on the liability scale in BioVU and Lifelines biobanks with all samples and case control ratio of ∼1:4. GLGS is a traditional case control cohort with best phenotyping and represents a homogenous POAG case set.

**Supplementary Figure S15:**
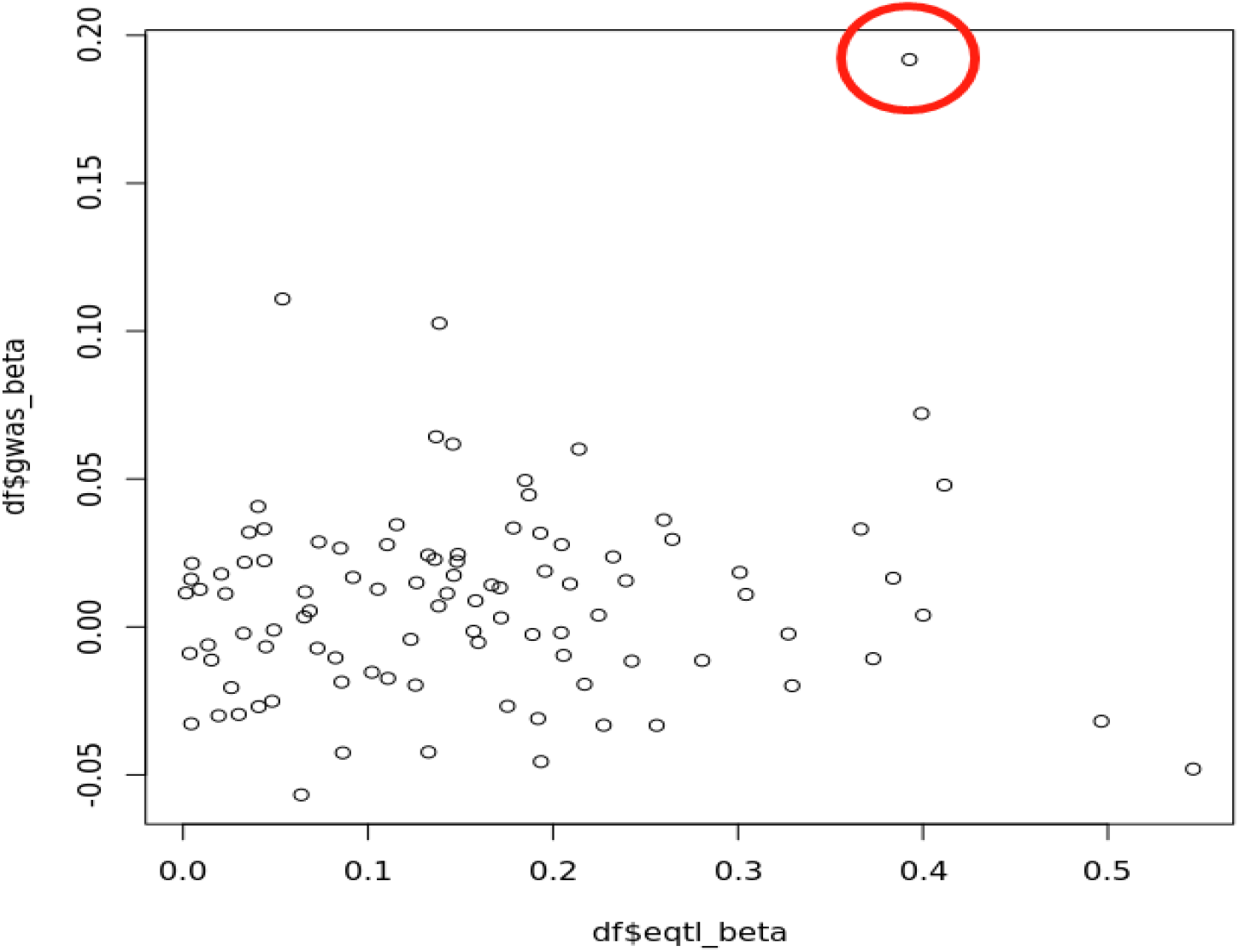
Brain Cortex eQTL_beta v.s. GWAS_beta scatter plot for the CDKN2A gene. TWAS result was dominated by the exonic CDKN2B-AS1 rs1008878 variant (the red circle). On excluding the top two signals in the locus, there was no significant TWAS signal.

**Supplementary Figure S16:**
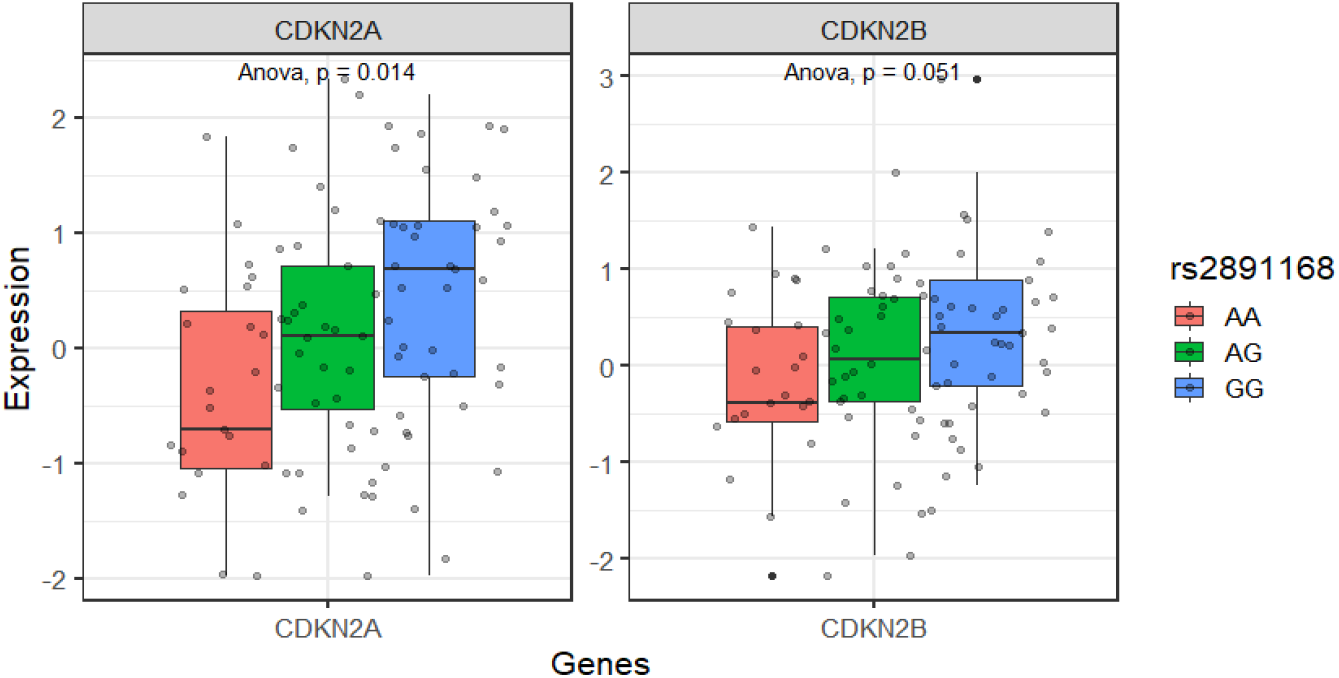
Expression pattern in individuals who carry wild type rs33912345_*SIX6* proxy variant in combination with rs2891168_CDKN2B-AS1 genotypes in brain cortex. p-values (p) are based on ANOVA tests.

**Supplementary Figure S17:**
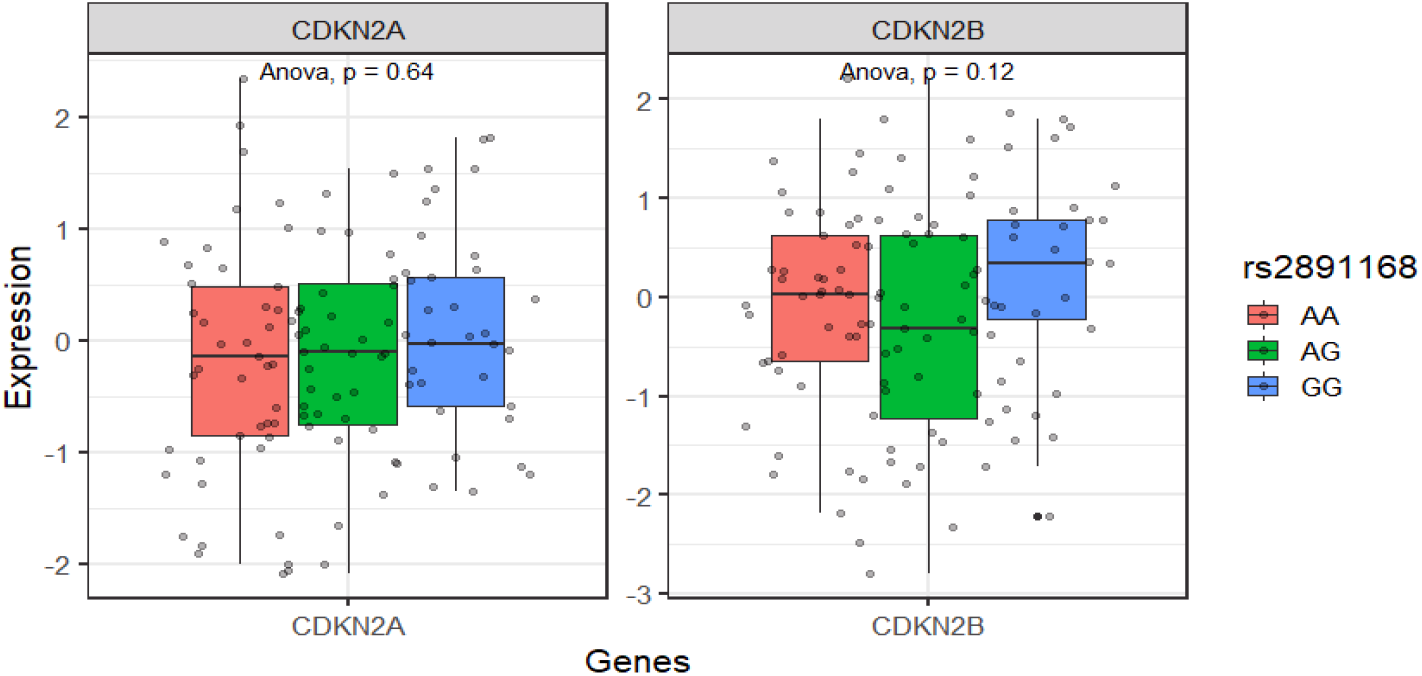
Expression pattern in individuals who carry rs33912345_*SIX6* proxy variant causal allele in combination with rs2891168_CDKN2B-AS1 genotypes in brain cortex. p-values (p) are based on ANOVA tests.

**Supplementary Figure S18:**
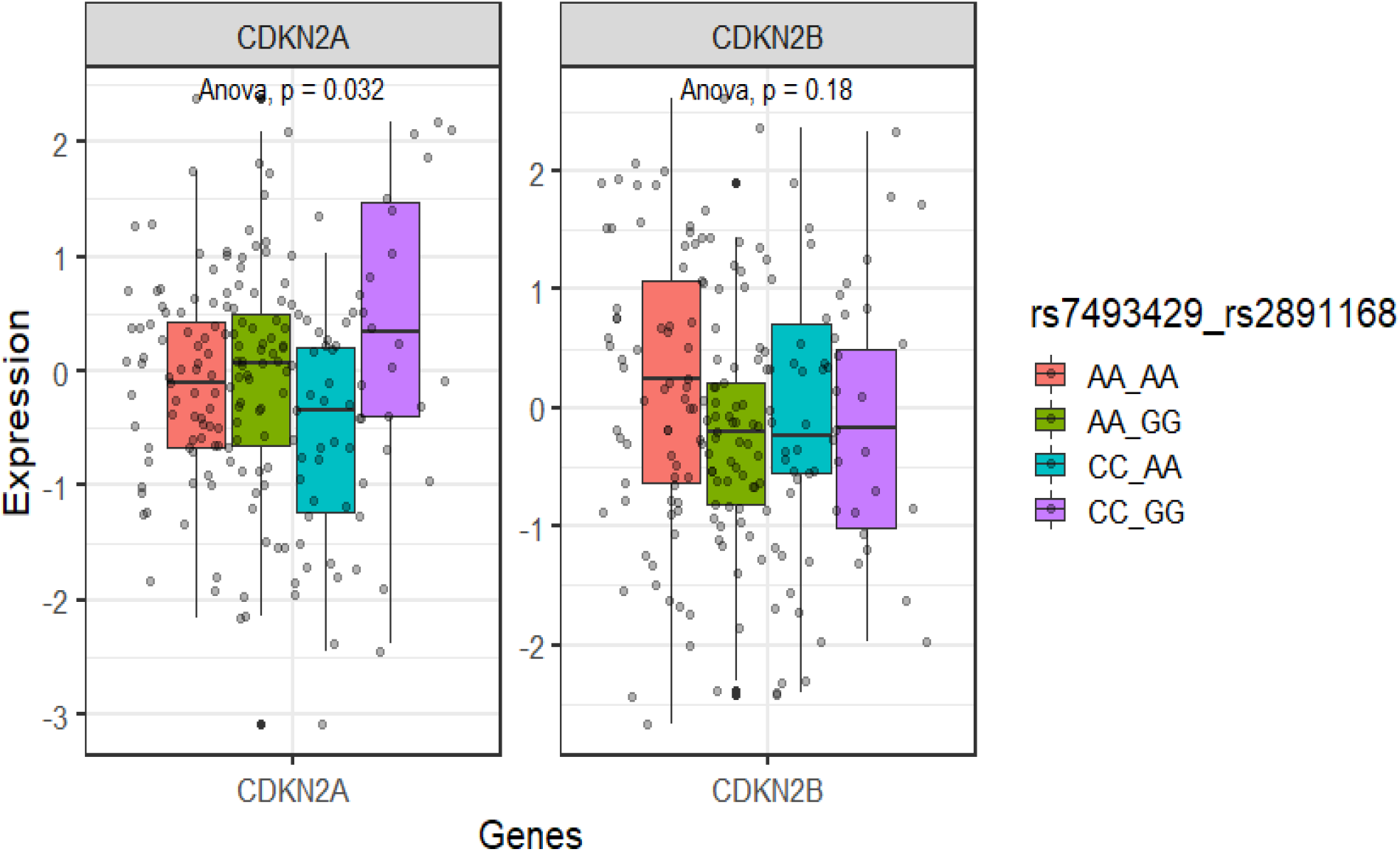
Expression pattern in individuals who carry homozygous rs33912345_*SIX6* proxy variant in combination with rs2891168_CDKN2B-AS1 genotypes in muscle skeletal. The eQTL effect is observed when the causal alleles are in both homozygous states. p-values (p) are based on ANOVA tests.

**Supplementary Table S1: Overview of sample size, gender, ancestry, and phenotyping of POAG in the biobanks participating in GBMI.** The last two columns report how POAG was defined in each cohort and potential inclusion of non-POAG glaucoma subtypes, respectively. OAG - other non-POAG Open angle glaucoma that potentially included using the phenotyping strategy, ICD10 H40.13 - pigmentary, H40.14 - capsular glaucoma with pseudoexfoliation, H40.15 - residual stage of open-angle glaucoma. Other glaucoma subtypes range from primary angle closure to glaucoma due to causes like trauma, inflammations and drugs

**Supplementary Table S2: BioVU as case example for possible inclusion of non-POAG as cases in phenotyping across the GBMI biobanks.**

**Supplementary Table S3: Summary statistics for the most significant genome-wide associated SNPs from the GWAS GBMI, conducted in 26,848 glaucoma cases and 1,460,599 controls.** The table reports results also for each GBMI biobank. In each case chromosome locations, effect sizes, p value, direction of effects for the lead variant is shown. The effect direction is for BBJ, BioMe_AMR, BioVU_AFR, BioVU_EUR, CCPM, DECODE, ESTBB, GS, HUNT, Lifelines, MGB, MGI, QSKIN, TWB, UKBB_SAS, UKBB_EUR, IGGC_EUR.

**Supplementary Table S4: Summary statistics for the most significant genome-wide associated SNPs from the GWAS GBMI female-stratified across 10 biobanks in 9,538 cases and 342,870 controls.** Loci index corresponds to those in Suppl. Table S3. For comparison, effect sizes and other parameters in male stratified analysis are shown.

**Supplementary Table S5: Summary statistics for the most significant genome-wide associated SNPs from the GWAS GBMI male-stratified genome-wide significant SNPs across 9 biobanks in 7,916 cases and 269,105 controls**. Loci index corresponds to those in Suppl. Table S3. For comparison, effect sizes and other parameters in male stratified analysis are shown.

**Supplementary Table S6: Male specific association of proxy rs17641032 with expression of *CELSR2* gene in GTEx data.** Significant results are reported in bold.

**Supplementary Table S7: All the loci identified in the GWAS meta-analysis performed in this study.** The first column lists the method used either inverse variance or MR-MEGA. Loci colored in red and blue are those identified in GBMI meta-analysis only using either inverse-variance or MR-MEGA, respectively, while loci in black are loci from the GWAS meta-analysis of GBMI, IGGC and GGLAD, using inverse-variance method. In each case chromosome locations, effect sizes, p value, direction of effects for the lead variant is shown. Loci that show heterogeneity in effect size biobank and ancestry specific effect sizes are shown in subsequent columns. The effect direction is for BBJ, BioMe_AMR, BioVU_AFR, BioVU_EUR, CCPM, DECODE, ESTBB, GS, HUNT, Lifelines, MGB, MGI, QSKIN, TWB, UKBB_SAS, UKBB_EUR, IGGC_EUR, GGLAD_AFR.

**Supplementary Table S8: Gene prioritization results in the meta-analysis of GBMI, IGGC and GGLAD.** The table provides the UCSC gene name, the p-values calculated by DEPICT, and lists the traits associated in GWAS catalog for each gene.

**Supplementary Table S9: Enrichment results for reconstituted gene sets representing canonical pathways from the GO, ENSG, MP, REACTOME and KEGG databases in the meta-analysis of GBMI, IGGC and GGLAD**. The first column lists the identifiers of the original gene set, the second and third columns list the gene description and the DEPICT gene set enrichment P-values.

**Supplementary Table S10: Tissue enrichment analyses in the meta-analysis of GBMI, IGGC and GGLAD.** The column lists the Medical Subject Headings (MeSH) terms and the DEPICT tissue enrichment p-values.

**Supplementary Table S11: TWAS results for GBMI-IGGC-GGLAD meta-analysis in GTEx v8 Brain cortex using JTI model.** For each gene number of other relevant GTEx tissues in which we observed significant association, the best association signal and tissue are listed.

**Supplementary Table S12: TWAS results of the GBMI-IGGC-GGLAD meta-analysis summary statistics in GTEx v8 data using JTI.** The sample size corresponds to 1,478,037 individuals, of which 46,325 cases and 1,431,712 controls. In addition, if gene association signals within the GWAS locus are identified using PrediXcan and UTMOST is indicated.

**Supplementary Table S13: Additional TWAS associations of the GBMI-IGGC-GGLAD meta-analysis summary statistics in GTEx v8 data using PrediXcan and UTMOST models.** The sample size corresponds to 1,478,037 individuals, of which 46,325 cases and 1,431,712 controls.

**Supplementary Table S14: TWAS gene signals involved in functions or implicated with vascular and neoplastic related traits.** For each gene we catalog vascular-related and/or cell senescence/proliferation functional roles or diseases, and if it has been reported as cilia-related gene. The references cited are reported in Supplementary Information. NA= not annotated according to literature review for the specific function.

**Supplementary Table S15: PheWAS results in BioVu of European ancestry.** PheWAS results (p-values, odd ratios, betas, standard errors, number of cases and controls) are shown for phenotype classes.

**Supplementary Table S16: PheWAS results in BioVu of African ancestry.** PheWAS results (p-values, odd ratios, betas, standard errors, number of cases and controls) are shown for phenotype classes.

**Supplementary Table S17: PheWAS results in UKB of European ancestry.** PheWAS results (p-values, odd ratios, betas, standard errors, number of cases and controls) are shown for phenotype classes.

**Supplementary Table S18: PheWAS results in UKB of African ancestry.** PheWAS results (p-values, odd ratios, betas, standard errors, number of cases and controls) are shown for phenotype classes.

**Supplementary Table S19: PheWAS results in UKB of Asian ancestry.** PheWAS results (p-values, odd ratios, betas, standard errors, number of cases and controls) are shown for phenotype classes.

**Supplementary Table S20: PheWAS results in EBB.** PheWAS results (p-values, odd ratios, betas, standard errors, number of cases and controls) are shown for phenotype classes.

**Supplementary Table S21: PheWAS results in BBJ.** PheWAS results (p-values, odd ratios, betas, standard errors, number of cases and controls) are shown for phenotype classes.

**Supplementary Table S22: GReX-PheWAS for CELSR2 gene across 731 traits and diseases grouped into 17 categories.** The table provides results of UKBB (n=396,618), BioVU (n=59,805), and meta-analysis of the two PheWAS (n=456,423).

**Supplementary Table S23: Genetic correlation between POAG and vascular and neoplastic traits.**

**Supplementary Table S24: Main and interaction effect of SIX6 and CDKN2A/CDKN2B variants in POAG and vascular traits, and the top chr9p21.3 GWAS variants that are associated with vascular and neoplastic traits in GWAS catalog (rs10811650 and rs2891168), analyzed in the BioVU cohort.** -POAG Primary open-angle glaucoma -CAD Coronary artery disease -HD Heart disease

## Supplementary Information

### Checking for Potential for phenotype heterogeneity in BioVU Samples

POAG was ascertained in the BioVU dataset using phecodes in VUMC electronic health records. All subjects (n=144,017) with an ophthalmology examination code (CPT = 92002, 92004, 92012, or 92014) were selected using at least two instances of POAG phecode that covers ICD-9 and/or ICD-10 codes 365.11, H40.11XX, 365.12, H40.12XX (n=17,824) while controls exclude those with codes for glaucoma or optic nerve disease (365.XX, 377.XX, H40.XXXX, H47.XXXX (n=53,919). The groups were then filtered to include only subjects with genotyping data from Illumina MEGA-Array.

The de-identified electronic health records of the glaucoma subjects were then reviewed by their BioVU number to confirm the diagnoses. The patients’ names, addresses, other forms of identification, and the treating physicians’ names were removed. Records which were available included the problem list stating POAG or NTG, medication lists, clinical notes including detailed ophthalmic examinations, ophthalmic operative notes, correspondence letters and discharge summaries. Additional high-level confirmation included an anterior segment examination without secondary-glaucoma evidence, and open angles by gonioscopy.

Only those subjects (n=1040) who had POAG reported in any of the above categories by the treating physicians were included as cases in the manual review. Any subject, who was coded for POAG without supporting confirmation or who had contradictory information in the medical record, was excluded to minimize inclusion errors. Supporting information including: reported peripheral vision loss, intraocular pressure, glaucomatous optic nerve appearance, glaucoma surgeries and use of glaucoma medications was reviewed and noted. A total of 716 individuals who had MEGA array genotyped have their EHR manually curated. A total of 220 out of the 716 individuals conflicted with manually curated records of which 172 are those with only a single ICD9+ICD10 mention of POAG. A total of 48 phecode assignations conflicted with manually curated records: 5 cases were missed by phecode phenotyping, 4 were pigmentary glaucoma, 7 POAG suspect, 9 pseudoexfoliation syndromes (XFS) and 23 primary angle closure glaucoma (PACG) (Suppl. Table S2). Using any mention of ICD9 or ICD10 POAG had a far much higher number of conflicts (114) with manually curated information. Thus conservatively, we estimate that ∼1.2% (∼320) and ∼3.2% (∼560) might potentially be XFS and PACG cases, respectively, among the total GBMI 26,848 POAG cases.

Therefore, as further quality check step, we compared our GBMI-IGGC-GGLAD meta-analysis signals with well powered GWASs for XFS and PACG, and determined that three of the loci might be attributed to these two glaucoma subtypes: 1) The rs3825942_*LOXL1* loci that is main XFS signal and previously reported in GWAS of glaucoma or recent POAG study that include Biobank level.^1,2^ 2) The rs11024102_*PLEKHA7* has been associated only with PACG and glaucoma in East Asian ancestry individuals, while the lead variant in rs58812088_ *FNDC3B* in our study is just ∼12kb away from and in high LD (r2=0.7) with PACG lead variant in rs16856870_*FNDC3B* locus.^3,4^ However, three loci: rs993471_*COL11A1*, rs2276035_*ARHGEF12,* rs12150284_*GAS7* have previously been shown to be POAG-PACG shared loci.^4,5^

### Groningen Longitudinal Glaucoma Study (GLGS)

The GLGS study consists of cases from the Groningen Longitudinal Glaucoma Study (GLGS). The GLGS consisted of Dutch individuals with a diagnosis of POAG from the hospital-based Groningen Longitudinal Glaucoma Study (GLGS). The original GLGS cohort has been described in detail by Heeg et al. (2005).^6^ After the inclusion of the initial cohort in 2000-2001, the GLGS continued as a dynamic population, that is, new participants were added during follow-up. We included glaucoma patients who visited the outpatient department of the University Medical Center Groningen in 2015 and who gave written informed consent for a blood sample being taken for genetic analyses. In the GLGS, glaucoma patients had to show glaucomatous visual field loss in at least one eye. For glaucomatous visual field loss, two consecutive tests had to be abnormal in at least one eye, after an initial test that was discarded to reduce the influence of learning. Defects had to be compatible with glaucoma and without any other explanation. Those with pseudoexfoliative or pigment dispersion glaucoma or a history of angle-closure or secondary glaucoma were excluded, leaving only POAG patients.^7^ In GLGS, genomic DNA was extracted from the peripheral blood, using Gentra Systems Purogene chemistry. The genotyping was done using the Illumina Infinium Global Screening Array® (GSA) MultiEthnic Disease beadchip version, which contains 692,367 markers.^8,9^

### Biobanks that contributed GWAS summary statistics to POAG study

#### Biobank Japan Project

The BioBank Japan Project was supported by the Tailor-Made Medical Treatment program of the Ministry of Education, Culture, Sports, Science, and Technology (MEXT), the Japan Agency for Medical Research and Development (AMED).

S.N. was supported by Takeda Science Foundation. Y.O. was supported by JSPS KAKENHI (19H01021, 20K21834), and AMED (JP21km0405211, JP21ek0109413, JP21ek0410075, JP21gm4010006, and JP21km0405217), JST Moonshot R&D (JPMJMS2021, JPMJMS2024), Takeda Science Foundation, and Bioinformatics Initiative of Osaka University Graduate School of Medicine, Osaka University.

#### BioMe - The Mount Sinai BioMe Biobank

The Mount Sinai BioMe Biobank has been supported by The Andrea and Charles Bronfman Philanthropies and in part by Federal funds from the NHLBI and NHGRI (U01HG00638001; U01HG007417; X01HL134588). We thank all participants in the Mount Sinai Biobank. We also thank all our recruiters who have assisted and continue to assist in data collection and management and are grateful for the computational resources and staff expertise provided by Scientific Computing at the Icahn School of Medicine at Mount Sinai.

#### BioVU

The BioVU projects at Vanderbilt University Medical Center are supported by numerous sources: institutional funding, private agencies, and federal grants. These include the NIH-funded Shared Instrumentation Grant S10OD017985 and S10RR025141; CTSA grants UL1TR002243, UL1TR000445, and UL1RR024975 from the National Center for Advancing Translational Sciences. Its contents are solely the responsibility of the authors and do not necessarily represent official views of the National Center for Advancing Translational Sciences or the National Institutes of Health. Genomic data are also supported by investigator-led projects that include U01HG004798, R01NS032830, RC2GM092618, P50GM115305, U01HG006378, U19HL065962, R01HD074711; and additional funding sources listed at https://victr.vumc.org/biovu-funding/.

#### Colorado Center for Personalized Medicine (CCPM)

The Colorado Center for Personalized Medicine (CCPM) would like to thank Richard Zane, Steve Hess, Sarah White, Emily Hearst, Emily Roberts and the entire Health Data Compass team. CCPM was developed with support from UCHealth, Children’s Hospital Colorado, CU Medicine, CU Department of Medicine and CU School of Medicine.

#### deCODE Genetics

We thank participants in deCODE cardiovascular and obesity studies and collaborators for their cooperation.

#### Estonian Biobank

This research was supported by the European Union through Horizon 2020 research and innovation programme under grant no 810645 and through the European Regional Development Fund project no. MOBEC008, by the Estonian Research Council grant PUT (PRG1291, PRG687 and PRG184) and by the European Union through the European Regional Development Fund project no. MOBERA21 (ERA-CVD project DETECT ARRHYTHMIAS, GA no JTC2018-009), Project No. 2014-2020.4.01.15-0012 and Project No. 2014-2020.4.01.16-0125. We would like to acknowledge Dr. Tõnu Esko; Dr. Lili Milani; Dr. Reedik Mägi, Dr. Mari Nelis and Dr. Andres Metspalu, all from the Institute of Genomics, University of Tartu, Tartu, Estonia.

#### FinnGen

The FinnGen project is funded by two grants from Business Finland (HUS 4685/31/2016 and UH 4386/31/2016) and the following industry partners: AbbVie Inc., AstraZeneca UK Ltd, Biogen MA Inc., Bristol Myers Squibb (and Celgene Corporation & Celgene International II Sàrl), Genentech Inc., Merck Sharp & Dohme Corp, Pfizer Inc., GlaxoSmithKline Intellectual Property Development Ltd., Sanofi US Services Inc., Maze Therapeutics Inc., Janssen Biotech Inc, and Novartis AG. Following biobanks are acknowledged for delivering biobank samples to FinnGen: Auria Biobank (www.auria.fi/biopankki), THL Biobank (www.thl.fi/biobank), Helsinki Biobank (www.helsinginbiopankki.fi), Biobank Borealis of Northern Finland (https://www.ppshp.fi/Tutkimus-ja-opetus/Biopankki/Pages/Biobank-Borealis-briefly-in-English.aspx),Finnish Clinical Biobank Tampere (www.tays.fi/en-US/Research_and_development/Finnish_Clinical_Biobank_Tampere), Biobank of Eastern Finland (www.ita-suomenbiopankki.fi/en), Central Finland Biobank (www.ksshp.fi/fi-FI/Potilaalle/Biopankki), Finnish Red Cross Blood Service Biobank (www.veripalvelu.fi/verenluovutus/biopankkitoiminta) and Terveystalo Biobank (www.terveystalo.com/fi/Yritystietoa/Terveystalo-Biopankki/Biopankki/). All Finnish Biobanks are members of BBMRI.fi infrastructure (www.bbmri.fi). Finnish Biobank Cooperative -FINBB (https://finbb.fi/) is the coordinator of BBMRI-ERIC operations in Finland. The Finnish biobank data can be accessed through the Fingenious® services (https://site.fingenious.fi/en/) managed by FINBB.

#### Generation Scotland

Generation Scotland received core support from the Chief Scientist Office of the Scottish Government Health Directorates [CZD/16/6] and the Scottish Funding Council [HR03006] and is currently supported by the Wellcome Trust [216767/Z/19/Z]. Genotyping of the GS:SFHS samples was carried out by the Genetics Core Laboratory at the Edinburgh Clinical Research Facility, University of Edinburgh, Scotland and was funded by the Medical Research Council UK and the Wellcome Trust (Wellcome Trust Strategic Award “STratifying Resilience and Depression Longitudinally” (STRADL) Reference 104036/Z/14/Z).”

#### The HUNT Study

A special thanks to all the HUNT participants for donating their time, samples and information to help others. The Trøndelag Health Study (HUNT) is a collaboration between HUNT Research Centre (Faculty of Medicine and Health Sciences, NTNU, Norwegian University of Science and Technology), Trøndelag County Council, Central Norway Regional Health Authority, and the Norwegian Institute of Public Health. The genotyping in HUNT was financed by the National Institutes of Health; University of Michigan; the Research Council of Norway; the Liaison Committee for Education, Research and Innovation in Central Norway; and the Joint Research Committee between St Olavs hospital and the Faculty of Medicine and Health Sciences, NTNU. The genetic investigations of the HUNT Study, is a collaboration between researchers from the K.G. Jebsen Center for Genetic Epidemiology, NTNU and the University of Michigan Medical School and the University of Michigan School of Public Health. The K.G. Jebsen Center for Genetic Epidemiology is financed by Stiftelsen Kristian Gerhard Jebsen; Faculty of Medicine and Health Sciences, NTNU, Norway. We want to thank clinicians and other employees at Nord-Trøndelag Hospital Trust for their support and for contributing to data collection in this research project.

We also acknowledge; HUNT-MI Leadership: Kristian Hveem, Cristen Willer, Oddgeir Lingaas Holmen, Mike Boehnke, Goncalo Abecasis, Bjorn Olav Åsvold, Ben Brumpton; Scientific Advisory Committee: Ele Zeggini, Mark Daly, Bjørn Pasternak; HUNT Research Centre: Jørn Søberg Fenstad, Anne Jorunn Vikdal, Marit Næss; HUNT Cloud: Oddgeir Lingaas Holmen, Sandor Zeestraten, Tom Erik Røberg; Data applications and registry linkages: Maiken E. Gabrielsen, Anne Heidi Skogholt; Low-pass whole sequencing genome bioinformatics and statistical analysis: He Zhang, Hyun Min Kang, Jin Chen; Array genotyping: Sten Even Erlandsen, Vidar Beisvåg; GWAS bioinformatics, QC, imputation and statistical analysis: Wei Zhou, Jonas Nielsen, Lars Fritsche, Hyun Min Kang, Oddgeir Holmen, Ben Brumpton, Laurent Thomas; CNV calling: Ellen Schmidt, Ryan Mills; Statistical methods development for analyzing HUNT data: Wei Zhou, Shawn Lee

### Funding

The K. G. Jebsen Centre for Genetic Epidemiology is financed by Stiftelsen Kristian Gerhard Jebsen. The genotyping in HUNT was financed by the National Institutes of Health; University of Michigan; the Research Council of Norway; Stiftelsen Kristian Gerhard Jebsen; the Liaison Committee for Education, Research and Innovation in Central Norway; and the Joint Research Committee between St Olav’s hospital and the Faculty of Medicine and Health Sciences,NTNU.

### LifeLines

The Lifelines Biobank initiative has been made possible by funding from the Dutch Ministry of Health, Welfare and Sport, the Dutch Ministry of Economic Affairs, the University Medical Center Groningen (UMCG the Netherlands), University of Groningen and the Northern Provinces of the Netherlands. The generation and management of GWAS genotype data for the Lifelines Cohort Study is supported by the UMCG Genetics Lifelines Initiative (UGLI). UGLI is partly supported by a Spinoza Grant from NWO, awarded to Cisca Wijmenga. The authors wish to acknowledge the services of the Lifelines Cohort Study, the contributing research centers delivering data to Lifelines, and all the study participants.

UMCG Genetics Lifelines Initiative (UGLI) team: Raul Aguirre-Gamboa (1), Patrick Deelen (1), Lude Franke (1), Jan A Kuivenhoven (2), Esteban A Lopera Maya (1), Ilja M Nolte (3), Serena Sanna (1), Harold Snieder (3), Morris A Swertz (1), Peter M. Visscher (3,4), Judith M Vonk (3), Cisca Wijmenga (1)

(1) Department of Genetics, University of Groningen, University Medical Center Groningen,The Netherlands

(2) Department of Pediatrics, University of Groningen, University Medical Center Groningen, The Netherlands

(3) Department of Epidemiology, University of Groningen, University Medical Center Groningen, The Netherlands

(4) Institute for Molecular Bioscience, The University of Queensland, Brisbane, Queensland, Australia.

### Mass General Brigham (MGB) Biobank

Samples, genomic data, and health information were obtained from the Mass General Brigham Biobank, a biorepository of consented patients samples at Mass General Brigham (parent organization of Massachusetts General Hospital and Brigham and Women’s Hospital). We are grateful to all of the participants and clinical and research teams who made this work possible. Support for genotyping was provided through MGB Personalized Medicine. MGB Biobank Leadership: Elizabeth W. Karlson, MD; Shawn N. Murphy, MD, PhD; Susan A Slaugenhaupt, PhD; Jordan W. Smoller, MD, ScD; Scott T. Weiss, MD, MSc

### Michigan Genomics Initiative

The authors acknowledge the Michigan Genomics Initiative participants, Precision Health at the University of Michigan, the University of Michigan Medical School Central Biorepository, and the University of Michigan Advanced Genomics Core for providing data and specimen storage, management, processing, and distribution services, and the Center for Statistical Genetics in the Department of Biostatistics at the School of Public Health for genotype data curation, imputation, and management in support of the research reported in this publication.

### QIMR Berghofer Biobank (QSKIN)

This work is supported by a project grant (APP1063061) and a Program grant (APP1073898) from the Australian National Health and Medical Research Council (NHMRC). SM is supported by a Research Fellowship from the NHMRC (Aust). NI received scholarship support from the University of Queensland and QIMR Berghofer Medical Research Institute. We thank research staff (David Whiteman, Catherine Olsen, Rachel Neale) and participants from the Australian QSKIN study (https://www.qimrberghofer.edu.au/study/qskin/). SEM is supported in part by APP1172917 from the Australian National Health and Medical Research Council (NHMRC).

### Taiwan Biobank

This research has been conducted using the Taiwan Biobank resource. We thank all the participants and investigators of the Taiwan Biobank. We thank the National Center for Genome Medicine of Taiwan for the technical support in genotyping. We thank the National Core Facility for Biopharmaceuticals (NCFB, MOST 106-2319-B-492-002) and National Center for High-performance Computing (NCHC) of National Applied Research Laboratories (NARLabs) of Taiwan for providing computational and storage resources.

### UK Biobank

Access to data from the UK BioBank was obtained through Application #31063 PI: Ben Neale, Claire Churchhouse Overview: “Methodological extensions to estimate genetic heritability and shared risk factors for phenotypes of the UK Biobank”. Website for Pan-UKBB results can be found: https://pan.ukbb.broadinstitute.org/

## Notes

### Competing Interest Statement

The spouse of Cristen Willer works at Regeneron Pharmaceuticals.
Eric R. Gamazon receives an honorarium from the journal Circulation Research of the American
Heart Association as a member of the Editorial Board.

### Funding Statement

Valeria Lo Faro was supported by the European Union Horizon 2020 research and innovation programme under the Marie Sklodowska-Curie grant agreement No.675033 (EGRET plus). Additional funding was provided by the Rotterdamse Stichting Blindenbelangen (grant ID B20150036).
Eric R. Gamazon is supported by the National Institutes of Health (NIH) Awards R35HG010718,
R01HG011138, R01GM140287, and NIH/NIA AG068026.
Karen Joos was supported by the Joseph Ellis Family and William Black Research Funds, and an
Unrestricted Departmental Grant to the Vanderbilt Eye Institute from Research to Prevent
Blindness, Inc., NY.

## REFERENCES

1. Quigley, H. A. & Broman, A. T. The number of people with glaucoma worldwide in 2010 and 2020. Br. J. Ophthalmol. 90, 262–267 (2006).

2. Gupta, N. et al. Chronic ocular hypertension induces dendrite pathology in the lateral geniculate nucleus of the brain. Exp. Eye Res. 84, 176–184 (2007).

3. Saccà, S. C. Oxidative DNA Damage in the Human Trabecular Meshwork. Archives of Ophthalmology vol. 123 458 (2005).

4. Nemesure, B., Honkanen, R., Hennis, A., Wu, S. Y. & Cristina Leske, M. Incident Open-angle Glaucoma and Intraocular Pressure. Ophthalmology vol. 114 1810–1815 (2007).

5. Gramer, E. & Grehn, F. Pathogenesis and Risk Factors of Glaucoma. (Springer Science & Business Media, 2012).

6. Tham, Y.-C. et al. Global prevalence of glaucoma and projections of glaucoma burden through 2040: a systematic review and meta-analysis. Ophthalmology 121, 2081–2090 (2014).

7. Zetterberg, M. Age-related eye disease and gender. Maturitas vol. 83 19–26 (2016).

8. Bonnemaijer, P. W. M. et al. Differences in clinical presentation of primary open-angle glaucoma between African and European populations. Acta Ophthalmol. 99, e1118–e1126 (2021).

9. Kyari, F., Bastawrous, A., Gilbert, C., Faal, H. & Abdull, M. Epidemiology of glaucoma in Sub-Saharan Africa: Prevalence, incidence and risk factors. Middle East African Journal of Ophthalmology vol. 20 111 (2013).

10. Evangelho, K., Mastronardi, C. A. & de-la-Torre, A. Experimental Models of Glaucoma: A Powerful Translational Tool for the Future Development of New Therapies for Glaucoma in Humans—A Review of the Literature. Medicina vol. 55 280 (2019).

11. Flammer, J. et al. The impact of ocular blood flow in glaucoma. Prog. Retin. Eye Res. 21, 359–393 (2002).

12. Shoeb Ahmad, S., Abdul Ghani, S. & Hemalata Rajagopal, T. Current Concepts in the Biochemical Mechanisms of Glaucomatous Neurodegeneration. J Curr Glaucoma Pract 7, 49–53 (2013).

13. Yanagi, M. et al. Vascular risk factors in glaucoma: a review. Clin. Experiment. Ophthalmol. 39, 252–258 (2011).

14. Emre, M., Orgül, S., Gugleta, K. & Flammer, J. Ocular blood flow alteration in glaucoma is related to systemic vascular dysregulation. Br. J. Ophthalmol. 88, 662–666 (2004).

15. Charlson, M. E. et al. Nocturnal Systemic Hypotension Increases the Risk of Glaucoma Progression. Ophthalmology vol. 121 2004–2012 (2014).

16. Jeganathan, V. S. E. et al. Peripheral artery disease and glaucoma: the singapore malay eye study. Arch. Ophthalmol. 127, 888–893 (2009).

17. Broadway, D. C. & Drance, S. M. Glaucoma and vasospasm. British Journal of Ophthalmology vol. 82 862–870 (1998).

18. Grunwald, J. E., Piltz, J., Hariprasad, S. M. & DuPont, J. Optic nerve and choroidal circulation in glaucoma. Invest. Ophthalmol. Vis. Sci. 39, 2329–2336 (1998).

19. Flammer, J. The vascular concept of glaucoma. Surv. Ophthalmol. 38 Suppl, S3–6 (1994).

20. Drance, S., Anderson, D. R., Schulzer, M. & Collaborative Normal-Tension Glaucoma Study Group. Risk factors for progression of visual field abnormalities in normal-tension glaucoma. Am. J. Ophthalmol. 131, 699–708 (2001).

21. Cursiefen, C. et al. Migraine and tension headache in high-pressure and normal-pressure glaucoma. Am. J. Ophthalmol. 129, 102–104 (2000).

22. Ko, F., et al. Diabetes, Triglyceride Levels, and Other Risk Factors for Glaucoma in the National Health and Nutrition Examination Survey 2005-2008. Invest. Ophthalmol. Vis. Sci. 57, 2152–2157 (2016).

23. Wise, L. A. et al. A prospective study of diabetes, lifestyle factors, and glaucoma among African-American women. Ann. Epidemiol. 21, 430–439 (2011).

24. Gharahkhani, P. et al. Genome-wide meta-analysis identifies 127 open-angle glaucoma loci with consistent effect across ancestries. Nat. Commun. 12, 1258 (2021).

25. Unlu, G. et al. GRIK5 Genetically Regulated Expression Associated with Eye and Vascular Phenomes: Discovery through Iteration among Biobanks, Electronic Health Records, and Zebrafish. Am. J. Hum. Genet. 104, 503–519 (2019).

26. Zhou, W. et al. Global Biobank Meta-analysis Initiative: powering genetic discovery across human diseases. medRxiv 2021.11.19.21266436 (2021).

27. Tanigawa, Y. et al. Rare protein-altering variants in ANGPTL7 lower intraocular pressure and protect against glaucoma. PLoS Genet. 16, e1008682 (2020).

28. Khawaja, A. P. et al. Genome-wide analyses identify 68 new loci associated with intraocular pressure and improve risk prediction for primary open-angle glaucoma. Nat. Genet. 50, 778–782 (2018).

29. Mägi, R. et al. Trans-ethnic meta-regression of genome-wide association studies accounting for ancestry increases power for discovery and improves fine-mapping resolution. Hum. Mol. Genet. 26, 3639–3650 (2017).

30. Allison, K., Patel, D. & Alabi, O. Epidemiology of Glaucoma: The Past, Present, and Predictions for the Future. Cureus 12, e11686 (2020).

31. Kapetanakis, V. V. et al. Global variations and time trends in the prevalence of primary open angle glaucoma (POAG): a systematic review and meta-analysis. British Journal of Ophthalmology vol. 100 86–93 (2016).

32. McMonnies, C. W. Glaucoma history and risk factors. J. Optom. 10, 71–78 (2017).

33. Craig, J. E. et al. Multitrait analysis of glaucoma identifies new risk loci and enables polygenic prediction of disease susceptibility and progression. Nat. Genet. 52, 160–166 (2020).

34. A global reference for human genetic variation. Nature 526, 68–74 (2015).

35. Karczewski, K. J. et al. The mutational constraint spectrum quantified from variation in 141,456 humans. Nature 581, 434–443 (2020).

36. Daya, M. et al. Association study in African-admixed populations across the Americas recapitulates asthma risk loci in non-African populations. Nat. Commun. 10, 880 (2019).

37. Belbin, G. M. et al. Toward a fine-scale population health monitoring system. Cell vol. 184 2068–2083.e11 (2021).

38. Vishnu, A. et al. The role of country of birth, and genetic and self-identified ancestry, in obesity susceptibility among African and Hispanic Americans. Am. J. Clin. Nutr. 110, 16–23 (2019).

39. Fadista, J., Oskolkov, N., Hansson, O. & Groop, L. LoFtool: a gene intolerance score based on loss-of-function variants in 60 706 individuals. Bioinformatics 33, 471–474 (2017).

40. GTEx Consortium. The Genotype-Tissue Expression (GTEx) project. Nat. Genet. 45, 580–585 (2013).

41. Bianconi, E. et al. Sex-Specific Transcriptome Differences in Human Adipose Mesenchymal Stem Cells. Genes 11, (2020).

42. Oliva, M. et al. The impact of sex on gene expression across human tissues. Science 369, (2020).

43. Du, L. et al. Downregulation of the ubiquitin ligase KBTBD8 prevented epithelial ovarian cancer progression. Mol. Med. 26, 96 (2020).

44. Yamada, Y. et al. Identification of 26 novel loci that confer susceptibility to early-onset coronary artery disease in a Japanese population. Biomed Rep 9, 383–404 (2018).

45. Kan, Z. et al. Diverse somatic mutation patterns and pathway alterations in human cancers. Nature 466, 869–873 (2010).

46. Simonson, B. et al. DDiT4L promotes autophagy and inhibits pathological cardiac hypertrophy in response to stress. Sci. Signal. 10, (2017).

47. Williams, S., Bateman, A. & O’Kelly, I. Altered expression of two-pore domain potassium (K2P) channels in cancer. PLoS One 8, e74589 (2013).

48. Pardo, L. A. & Stühmer, W. The roles of K channels in cancer. Nature Reviews Cancer vol. 14 39–48 (2014).

49. Alvarez-Baron, C. P., Jonsson, P., Thomas, C., Dryer, S. E. & Williams, C. The Two-Pore Domain Potassium Channel KCNK5: Induction by Estrogen Receptor α and Role in Proliferation of Breast Cancer Cells. Molecular Endocrinology vol. 25 1326–1336 (2011).

50. van der Harst, P. & Verweij, N. Identification of 64 Novel Genetic Loci Provides an Expanded View on the Genetic Architecture of Coronary Artery Disease. Circ. Res. 122, 433–443 (2018).

51. Tsukasaki, K. et al. Mutations in the mitotic check point gene, MAD1L1, in human cancers. Oncogene 20, 3301–3305 (2001).

52. Barbieri, M. et al. Association of genetic variation in adaptor protein APPL1/APPL2 loci with non-alcoholic fatty liver disease. PLoS One 8, e71391 (2013).

53. Ma, X.-W., Ding, S., Ma, X.-D., Gu, N. & Guo, X.-H. Genetic variability in adapter proteins with APPL1/2 is associated with the risk of coronary artery disease in type 2 diabetes mellitus in Chinese Han population. Chin. Med. J. 124, 3618–3621 (2011).

54. Li, Y. et al. Long non-coding RNA FENDRR inhibits cell proliferation and is associated with good prognosis in breast cancer. Onco. Targets. Ther. 11, 1403–1412 (2018).

55. Liu, J. & Du, W. LncRNA FENDRR attenuates colon cancer progression by repression of SOX4 protein. Onco. Targets. Ther. 12, 4287–4295 (2019).

56. Li, N. et al. Causal variants screened by whole exome sequencing in a patient with maternal uniparental isodisomy of chromosome 10 and a complicated phenotype. Experimental and Therapeutic Medicine vol. 11 2247–2253 (2016).

57. van den Berg, M. E., et al. Discovery of novel heart rate-associated loci using the Exome Chip. Hum. Mol. Genet. 26, 2346–2363 (2017).

58. Yehia, L. et al. Germline Heterozygous Variants in SEC23B Are Associated with Cowden Syndrome and Enriched in Apparently Sporadic Thyroid Cancer. Am. J. Hum. Genet. 97, 661– 676 (2015).

59. Yang, C. et al. Mutations in the coat complex II component SEC23B promote colorectal cancer metastasis. Cell Death Dis. 11, 157 (2020).

60. Wagner, A. H. et al. Exon-level expression profiling of ocular tissues. Exp. Eye Res. 111, 105– 111 (2013).

61. Bryan, J. M. et al. Identifying core biological processes distinguishing human eye tissues with precise systems-level gene expression analyses and weighted correlation networks. Hum. Mol. Genet. 27, 3325–3339 (2018).

62. Thul, P. J. et al. A subcellular map of the human proteome. Science 356, (2017).

63. Uhlén, M., et al. Proteomics. Tissue-based map of the human proteome. Science 347, 1260419 (2015).

64. Wang, Y., et al. Global biobank analyses provide lessons for computing polygenic risk scores across diverse cohorts. medRxiv 2021.11.18.21266545 (2021).

65. Martin, A. R. et al. Clinical use of current polygenic risk scores may exacerbate health disparities. Nat. Genet. 51, 584–591 (2019).

66. Martin, A. R. et al. Human Demographic History Impacts Genetic Risk Prediction across Diverse Populations. Am. J. Hum. Genet. 100, 635–649 (2017).

67. Duncan, L. et al. Analysis of polygenic risk score usage and performance in diverse human populations. Nat. Commun. 10, 1–9 (2019).

68. Wang, Y. et al. Theoretical and empirical quantification of the accuracy of polygenic scores in ancestry divergent populations. Nat. Commun. 11, 3865 (2020).

69. Gamazon, E. R. et al. A gene-based association method for mapping traits using reference transcriptome data. Nat. Genet. 47, 1091–1098 (2015).

70. Hu, Y. et al. A statistical framework for cross-tissue transcriptome-wide association analysis. Nat. Genet. 51, 568–576 (2019).

71. Zhou, D. et al. A unified framework for joint-tissue transcriptome-wide association and Mendelian randomization analysis. Nat. Genet. 52, 1239–1246 (2020).

72. Zhang, H. et al. Lipidomics reveals carnitine palmitoyltransferase 1C protects cancer cells from lipotoxicity and senescence. Journal of Pharmaceutical Analysis vol. 11 340–350 (2021).

73. Millner, A. & Atilla-Gokcumen, G. E. Lipid Players of Cellular Senescence. Metabolites 10, (2020).

74. Wang, L. et al. Peakwide mapping on chromosome 3q13 identifies the kalirin gene as a novel candidate gene for coronary artery disease. Am. J. Hum. Genet. 80, 650–663 (2007).

75. Ji, J.-Z. et al. CNTF promotes survival of retinal ganglion cells after induction of ocular hypertension in rats: the possible involvement of STAT3 pathway. Eur. J. Neurosci. 19, 265–272 (2004).

76. Pease, M. E. et al. Effect of CNTF on retinal ganglion cell survival in experimental glaucoma. Invest. Ophthalmol. Vis. Sci. 50, 2194–2200 (2009).

77. Shpak, A. A., Guekht, A. B., Druzhkova, T. A., Kozlova, K. I. & Gulyaeva, N. V. Ciliary neurotrophic factor in patients with primary open-angle glaucoma and age-related cataract. Mol. Vis. 23, 799–809 (2017).

78. Xu, S. et al. The role of collagen in cancer: from bench to bedside. J. Transl. Med. 17, 309 (2019).

79. Youle, R. J. & van der Bliek, A. M. Mitochondrial Fission, Fusion, and Stress. Science vol. 337 1062–1065 (2012).

80. Finak, G. et al. Stromal gene expression predicts clinical outcome in breast cancer. Nat. Med. 14, 518–527 (2008).

81. Karnoub, A. E. et al. Mesenchymal stem cells within tumour stroma promote breast cancer metastasis. Nature 449, 557–563 (2007).

82. Chiappetta, G. et al. High mobility group HMGI(Y) protein expression in human colorectal hyperplastic and neoplastic diseases. International Journal of Cancer vol. 91 147–151 (2001).

83. Zhou, X. & Chada, K. HMGI family proteins: architectural transcription factors in mammalian development and cancer. Keio J. Med. 47, 73–77 (1998).

84. Krupenko, S. A. & Krupenko, N. I. ALDH1L1 and ALDH1L2 Folate Regulatory Enzymes in Cancer. Adv. Exp. Med. Biol. 1032, 127–143 (2018).

85. Gonzalez, J. M., Jr. Existence of the canonical Wnt signaling pathway in the human trabecular meshwork. Investigative ophthalmology & visual science vol. 53 6972 (2012).

86. Wang, W.-H. et al. Increased expression of the WNT antagonist sFRP-1 in glaucoma elevates intraocular pressure. J. Clin. Invest. 118, 1056–1064 (2008).

87. Wang, X. et al. Mutual regulation of the Hippo/Wnt/LPA/TGF-β signaling pathways and their roles in glaucoma (Review). International Journal of Molecular Medicine (2017) doi:10.3892/ijmm.2017.3352.

88. Tan, J. et al. CELSR2 deficiency suppresses lipid accumulation in hepatocyte by impairing the UPR and elevating ROS level. FASEB J. 35, e21908 (2021).

89. Hjeij, R. et al. ARMC4 mutations cause primary ciliary dyskinesia with randomization of left/right body asymmetry. Am. J. Hum. Genet. 93, 357–367 (2013).

90. Onoufriadis, A. et al. Combined exome and whole-genome sequencing identifies mutations in ARMC4 as a cause of primary ciliary dyskinesia with defects in the outer dynein arm. J. Med. Genet. 51, 61–67 (2014).

91. Samani, N. J. et al. The novel genetic variant predisposing to coronary artery disease in the region of the PSRC1 and CELSR2 genes on chromosome 1 associates with serum cholesterol. J. Mol. Med. 86, 1233–1241 (2008).

92. Jiang, L. et al. Differential cellular localization of CELSR2 and ING4 and correlations with hormone receptor status in breast cancer. Histol. Histopathol. 33, 835–842 (2018).

93. Yamada, Y. et al. Identification of differentially methylated CpG islands in prostate cancer. Int. J. Cancer 112, 840–845 (2004).

94. Xu, M., Zhu, S., Xu, R. & Lin, N. Identification of CELSR2 as a novel prognostic biomarker for hepatocellular carcinoma. BMC Cancer 20, 313 (2020).

95. Namba, S., et al. A practical guideline of genomics-driven drug discovery in the era of global biobank meta-analysis. medRxiv 2021.12.03.21267280 (2021).

96. Cole, B. S. et al. The Role of Genetic Ancestry as a Risk Factor for Primary Open-angle Glaucoma in African Americans. Invest. Ophthalmol. Vis. Sci. 62, 28 (2021).

97. Zhang, Y., Yan, L., Liu, J., Cui, S. & Qiu, J. cGMP-dependent protein kinase II determines β-catenin accumulation that is essential for uterine decidualization in mice. Am. J. Physiol. Cell Physiol. 317, C1115–C1127 (2019).

98. Fotesko, K., Thomsen, B. S. V., Kolko, M. & Vohra, R. Girl Power in Glaucoma: The Role of Estrogen in Primary Open Angle Glaucoma. Cell. Mol. Neurobiol. (2020) doi:10.1007/s10571-020-00965-5.

99. Salowe, R. et al. Primary Open-Angle Glaucoma in Individuals of African Descent: A Review of Risk Factors. J. Clin. Exp. Ophthalmol. 6, (2015).

100. Siesky, B. et al. Differences in ocular blood flow in glaucoma between patients of African and European descent. J. Glaucoma 24, 117–121 (2015).

101. The Advanced Glaucoma Intervention Study (AGIS): 3. Baseline characteristics of black and white patients. Ophthalmology 105, 1137–1145 (1998).

102. Khachatryan, N. et al. Primary Open-Angle African American Glaucoma Genetics (POAAGG) Study: gender and risk of POAG in African Americans. PLoS One 14, e0218804 (2019).

103. Ye, X., She, X. & Shen, L. Association of sex with the global burden of glaucoma: an analysis from the global burden of disease study 2017. Acta Ophthalmol. (2020) doi:10.1111/aos.14330.

104. Vasconcellos, J. P. C. et al. Association of SALL1 rs1362756 and SIX1/SIX6 rs33912345 variants with POAG in a Brazilian population. Invest. Ophthalmol. Vis. Sci. 61, 1250–1250 (2020).

105. Carnes, M. U. et al. Discovery and functional annotation of SIX6 variants in primary open-angle glaucoma. PLoS Genet. 10, e1004372 (2014).

106. Teotia, P. et al. Modeling Glaucoma: Retinal Ganglion Cells Generated from Induced Pluripotent Stem Cells of Patients with SIX6 Risk Allele Show Developmental Abnormalities. Stem Cells 35, 2239–2252 (2017).

107. Iglesias, A. I. et al. Exome sequencing and functional analyses suggest that SIX6 is a gene involved in an altered proliferation–differentiation balance early in life and optic nerve degeneration at old age. Human Molecular Genetics vol. 23 1320–1332 (2014).

108. Skowronska-Krawczyk, D. et al. P16INK4a Upregulation Mediated by SIX6 Defines Retinal Ganglion Cell Pathogenesis in Glaucoma. Mol. Cell 59, 931–940 (2015).

109. Staples, C. J. et al. Ccdc13 is a novel human centriolar satellite protein required for ciliogenesis and genome stability. J. Cell Sci. 127, 2910–2919 (2014).

110. Yang, N. et al. INTU is essential for oncogenic Hh signaling through regulating primary cilia formation in basal cell carcinoma. Oncogene 36, 4997–5005 (2017).

111. Nobbio, L. et al. Impaired expression of ciliary neurotrophic factor in Charcot-Marie-Tooth type 1A neuropathy. J. Neuropathol. Exp. Neurol. 68, 441–455 (2009).

112. Treutlein, B. et al. Reconstructing lineage hierarchies of the distal lung epithelium using single-cell RNA-seq. Nature 509, 371–375 (2014).

113. Steggink, L. C. et al. Genome-wide association study of cardiovascular disease in testicular cancer patients treated with platinum-based chemotherapy. Pharmacogenomics J. 21, 152–164 (2021).

114. Tan, P. Intrinsic Subtypes of Gastric Cancer, Based on Gene Expression Pattern, Predict Survival and Respond Differently to Chemotherapy. SciVee (2011) doi:10.4016/32808.01.

115. Liang, Y. et al. Discovery of Aberrant Alteration of Genome in Colorectal Cancer by Exome Sequencing. Am. J. Med. Sci. 358, 340–349 (2019).

116. Boutin, T. S. et al. Insights into the genetic basis of retinal detachment. Hum. Mol. Genet. 29, 689–702 (2020).

117. Kichaev, G. et al. Leveraging Polygenic Functional Enrichment to Improve GWAS Power. Am. J. Hum. Genet. 104, 65–75 (2019).

118. Ma, N. & Zhou, J. Functions of Endothelial Cilia in the Regulation of Vascular Barriers. Front Cell Dev Biol 8, 626 (2020).

119. Pala, R., Jamal, M., Alshammari, Q. & Nauli, S. The Roles of Primary Cilia in Cardiovascular Diseases. Cells vol. 7 233 (2018).

120. Sun, C., Zhou, J. & Meng, X. Primary cilia in retinal pigment epithelium development and diseases. J. Cell. Mol. Med. 25, 9084–9088 (2021).

121. May-Simera, H. L. et al. Primary Cilium-Mediated Retinal Pigment Epithelium Maturation Is Disrupted in Ciliopathy Patient Cells. Cell Rep. 22, 189–205 (2018).

122. Dilan, T. L. et al. Bardet-Biedl syndrome-8 (BBS8) protein is crucial for the development of outer segments in photoreceptor neurons. Hum. Mol. Genet. 27, 283–294 (2018).

123. Zhang, W., Li, L., Su, Q., Gao, G. & Khanna, H. Gene Therapy Using a miniCEP290 Fragment Delays Photoreceptor Degeneration in a Mouse Model of Leber Congenital Amaurosis. Human Gene Therapy vol. 29 42–50 (2018).

124. Zhou, P. & Zhou, J. The Primary Cilium as a Therapeutic Target in Ocular Diseases. Front. Pharmacol. 11, 977 (2020).

125. Hackam, A. S. The Wnt signaling pathway in retinal degenerations. IUBMB Life 57, 381–388 (2005).

126. Lo Faro, V., Ten Brink, J. B., Snieder, H., Jansonius, N. M. & Bergen, A. A. Genome-wide CNV investigation suggests a role for cadherin, Wnt, and p53 pathways in primary open-angle glaucoma. BMC Genomics 22, 590 (2021).

127. Takahashi, H. et al. The myocilin (MYOC) gene expression in the human trabecular meshwork. Curr. Eye Res. 20, 81–84 (2000).

128. Itakura, T., Peters, D. M. & Fini, M. E. Glaucomatous MYOC mutations activate the IL-1/NF-κB inflammatory stress response and the glaucoma marker SELE in trabecular meshwork cells. Mol. Vis. 21, 1071–1084 (2015).

129. O’Brien, E. T., Ren, X. & Wang, Y. Localization of myocilin to the golgi apparatus in Schlemm’s canal cells. Invest. Ophthalmol. Vis. Sci. 41, 3842–3849 (2000).

130. Scholtens, S. et al. Cohort Profile: LifeLines, a three-generation cohort study and biobank. Int. J. Epidemiol. 44, 1172–1180 (2015).

131. Genetics of Glaucoma in People of African Descent (GGLAD) Consortium, et al. Association of Genetic Variants With Primary Open-Angle Glaucoma Among Individuals With African Ancestry. JAMA 322, 1682–1691 (2019).

132. Purcell, S. et al. PLINK: A Tool Set for Whole-Genome Association and Population-Based Linkage Analyses. The American Journal of Human Genetics vol. 81 559–575 (2007).

133. Wang, K., Li, M. & Hakonarson, H. ANNOVAR: functional annotation of genetic variants from high-throughput sequencing data. Nucleic Acids Research vol. 38 e164–e164 (2010).

134. Oscanoa, J. et al. SNPnexus: a web server for functional annotation of human genome sequence variation (2020 update). Nucleic Acids Research vol. 48 W185–W192 (2020).

135. Sim, N.-L. et al. SIFT web server: predicting effects of amino acid substitutions on proteins. Nucleic Acids Res. 40, W452–7 (2012).

136. Adzhubei, I. A. et al. A method and server for predicting damaging missense mutations. Nat. Methods 7, 248–249 (2010).

137. Pers, T. H. et al. Biological interpretation of genome-wide association studies using predicted gene functions. Nat. Commun. 6, 5890 (2015).

138. Barbeira, A. N. et al. Exploring the phenotypic consequences of tissue specific gene expression variation inferred from GWAS summary statistics. Nat. Commun. 9, 1825 (2018).

139. Friedland, A. B. Relationship between arterial pulsations and intraocular pressure. Exp. Eye Res. 37, 421–428 (1983).

140. Guidoboni, G. et al. Neurodegenerative Disorders of the Eye and of the Brain: A Perspective on Their Fluid-Dynamical Connections and the Potential of Mechanism-Driven Modeling. Frontiers in Neuroscience vol. 14 (2020).

141. Chan, J. W., Chan, N. C. Y. & Sadun, A. A. Glaucoma as Neurodegeneration in the Brain. Eye and Brain vol. 13 21–28 (2021).

142. Consortium, T. G. & The GTEx Consortium. The GTEx Consortium atlas of genetic regulatory effects across human tissues. Science vol. 369 1318–1330 (2020).

143. Website. Michigan Imputation Server. https://imputationserver.sph.umich.edu/.

144. Delaneau, O., Marchini, J., 1000 Genomes Project Consortium & 1000 Genomes Project Consortium. Integrating sequence and array data to create an improved 1000 Genomes Project haplotype reference panel. Nat. Commun. 5, 3934 (2014).

145. Mancuso, N. et al. Probabilistic fine-mapping of transcriptome-wide association studies. Nat. Genet. 51, 675–682 (2019).

146. Lo Faro, V., et al. Mitochondrial genome study identifies association between primary open-angle glaucoma and variants in MT-CYB, MT-ND4 genes and haplogroups. Front. Genet. 12:781189 (2021).

147. Bulik-Sullivan, B. K. et al. LD Score regression distinguishes confounding from polygenicity in genome-wide association studies. Nature Genetics vol. 47 291–295 (2015).

148. MacArthur, J. et al. The new NHGRI-EBI Catalog of published genome-wide association studies (GWAS Catalog). Nucleic Acids Research vol. 45 D896–D901 (2017).

149. Bailey, J. N. C. et al. Genome-wide association analysis identifies TXNRD2, ATXN2 and FOXC1 as susceptibility loci for primary open-angle glaucoma. Nat. Genet. 48, 189–194 (2016).

150. Osman, W., Low, S.-K., Takahashi, A., Kubo, M. & Nakamura, Y. A genome-wide association study in the Japanese population confirms 9p21 and 14q23 as susceptibility loci for primary open angle glaucoma. Hum. Mol. Genet. 21, 2836–2842 (2012).

151. Springelkamp, H. et al. New insights into the genetics of primary open-angle glaucoma based on meta-analyses of intraocular pressure and optic disc characteristics. Hum. Mol. Genet. 26, 438– 453 (2017).

152. Gharahkhani, P. et al. Common variants near ABCA1, AFAP1 and GMDS confer risk of primary open-angle glaucoma. Nat. Genet. 46, 1120–1125 (2014).

153. Nakano, M. et al. Common variants in CDKN2B-AS1 associated with optic-nerve vulnerability of glaucoma identified by genome-wide association studies in Japanese. PLoS One 7, e33389 (2012).

154. Li, Z. et al. A common variant near TGFBR3 is associated with primary open angle glaucoma. Hum. Mol. Genet. 24, 3880–3892 (2015).

155. Fan, B. J., Wang, D. Y., Pasquale, L. R., Haines, J. L. & Wiggs, J. L. Genetic Variants Associated with Optic Nerve Vertical Cup-to-Disc Ratio Are Risk Factors for Primary Open Angle Glaucoma in a US Caucasian Population. Investigative Opthalmology & Visual Science vol. 52 1788 (2011).

156. Ramdas, W. D. et al. A Genome-Wide Association Study of Optic Disc Parameters. PLoS Genetics vol. 6 e1000978 (2010).

157. Cheng, C.-Y. et al. Association of common SIX6 polymorphisms with peripapillary retinal nerve fiber layer thickness: the Singapore Chinese Eye Study. Invest. Ophthalmol. Vis. Sci. 56, 478–483 (2014).

158. Wiggs, J. L. et al. Common variants at 9p21 and 8q22 are associated with increased susceptibility to optic nerve degeneration in glaucoma. PLoS Genet. 8, e1002654 (2012).

159. Cao, D. et al. CDKN2B polymorphism is associated with primary open-angle glaucoma (POAG) in the Afro-Caribbean population of Barbados, West Indies. PLoS One 7, e39278 (2012).

160. Shiga, Y. et al. Genome-wide association study identifies seven novel susceptibility loci for primary open-angle glaucoma. Hum. Mol. Genet. 27, 1486–1496 (2018).

161. Mori, K., et al. Stronger Association of CDKN2B-AS1 Variants in Female Normal-Tension Glaucoma Patients in a Japanese Population. Investigative Opthalmology & Visual Science vol. 57 6416 (2016).

162. Ng, S. K. et al. Genetic Association at the 9p21 Glaucoma Locus Contributes to Sex Bias in Normal-Tension Glaucoma. Investigative Opthalmology & Visual Science vol. 57 3416 (2016).

163. Chen, Y. et al. Genetic Variants Associated With Different Risks for High Tension Glaucoma and Normal Tension Glaucoma in a Chinese Population. Investigative Opthalmology & Visual Science vol. 56 2595 (2015).

164. Taylor, K. D. et al. Genetic Architecture of Primary Open-Angle Glaucoma in Individuals of African Descent: The African Descent and Glaucoma Evaluation Study III. Ophthalmology 126, 38–48 (2019).

165. Rong, S. S. et al. Association of the SIX6 locus with primary open angle glaucoma in southern Chinese and Japanese. Exp. Eye Res. 180, 129–136 (2019).

166. Zhao, W. et al. The cis and trans effects of the risk variants of coronary artery disease in the Chr9p21 region. BMC Med. Genomics 8, 21 (2015).

167. Myers, T. A., Chanock, S. J. & Machiela, M. J. LDlinkR: An R Package for Rapidly Calculating Linkage Disequilibrium Statistics in Diverse Populations. Frontiers in Genetics vol. 11 (2020).

168. Koyama, S. et al. Population-specific and trans-ancestry genome-wide analyses identify distinct and shared genetic risk loci for coronary artery disease. Nat. Genet. 52, 1169–1177 (2020).

169. Nikpay, M. et al. A comprehensive 1,000 Genomes-based genome-wide association meta-analysis of coronary artery disease. Nat. Genet. 47, 1121–1130 (2015).

170. Hartiala, J. A. et al. Genome-wide analysis identifies novel susceptibility loci for myocardial infarction. Eur. Heart J. 42, 919–933 (2021).

171. Wu, Y. et al. Genome-wide association study of medication-use and associated disease in the UK Biobank. Nat. Commun. 10, 1891 (2019).

172. Nelson, C. P. et al. Association analyses based on false discovery rate implicate new loci for coronary artery disease. Nat. Genet. 49, 1385–1391 (2017).

173. Michailidou, K. et al. Association analysis identifies 65 new breast cancer risk loci. Nature 551, 92–94 (2017).

174. Matsunaga, H., et al. Transethnic Meta-Analysis of Genome-Wide Association Studies Identifies Three New Loci and Characterizes Population-Specific Differences for Coronary Artery Disease. Circ Genom Precis Med 13, e002670 (2020).

175. Landi, M. T. et al. Genome-wide association meta-analyses combining multiple risk phenotypes provide insights into the genetic architecture of cutaneous melanoma susceptibility. Nat. Genet. 52, 494–504 (2020).

176. Zhou, W. et al. Efficiently controlling for case-control imbalance and sample relatedness in large-scale genetic association studies. Nat. Genet. 50, 1335–1341 (2018).

177. Roden, D. M. et al. Development of a large-scale de-identified DNA biobank to enable personalized medicine. Clin. Pharmacol. Ther. 84, 362–369 (2008).

## REFERENCES

1. Aung, T. et al. Genetic association study of exfoliation syndrome identifies a protective rare variant at LOXL1 and five new susceptibility loci. Nat. Genet. 49, 993–1004 (2017).

2. Buniello, A. et al. The NHGRI-EBI GWAS Catalog of published genome-wide association studies, targeted arrays and summary statistics 2019. Nucleic Acids Res. 47, D1005–D1012 (2019).

3. Vithana, E. N. et al. Genome-wide association analyses identify three new susceptibility loci for primary angle closure glaucoma. Nat. Genet. 44, 1142–1146 (2012).

4. Khor, C. C. et al. Genome-wide association study identifies five new susceptibility loci for primary angle closure glaucoma. Nat. Genet. 48, 556–562 (2016).

5. Choquet, H. et al. A multiethnic genome-wide association study of primary open-angle glaucoma identifies novel risk loci. Nat. Commun. 9, 2278 (2018).

6. Heeg, G. P., Blanksma, L. J., Hardus, P. L. L. & Jansonius, N. M. The Groningen Longitudinal Glaucoma Study. I. Baseline sensitivity and specificity of the frequency doubling perimeter and the GDx nerve fibre analyser. Acta Ophthalmologica Scandinavica vol. 83 46–52 (2005).

7. Bonnemaijer, P. W. M. et al. Differences in clinical presentation of primary open-angle glaucoma between African and European populations. Acta Ophthalmol. 99, e1118–e1126 (2021).

8. Lo Faro, V., Ten Brink, J. B., Snieder, H., Jansonius, N. M. & Bergen, A. A. Genome-wide CNV investigation suggests a role for cadherin, Wnt, and p53 pathways in primary open-angle glaucoma. BMC Genomics 22, 590 (2021).

9. Lo Faro, V., et al. Mitochondrial genome study identifies association between primary open-angle glaucoma and variants in MT-CYB, MT-ND4 genes and haplogroups. Front. Genet. 0, (1AD).

10. Tsoi, H. et al. A novel missense mutation in CCDC88C activates the JNK pathway and causes a dominant form of spinocerebellar ataxia. J. Med. Genet. 51, 590–595 (2014).

11. Wang, H.-Y. et al. Protein Kinase A-Mediated Septin7 Phosphorylation Disrupts Septin Filaments and Ciliogenesis. Cells 10, (2021).

12. Scheffer, D. I., Shen, J., Corey, D. P. & Chen, Z.-Y. Gene Expression by Mouse Inner Ear Hair Cells during Development. Journal of Neuroscience vol. 35 6366–6380 (2015).

13. Wu, M.-M. et al. Lovastatin attenuates hypertension induced by renal tubule-specific knockout of ATP-binding cassette transporter A1, by inhibiting epithelial sodium channels. Br. J. Pharmacol. 176, 3695–3711 (2019).

14. Zappaterra, M., Gioiosa, S., Chillemi, G., Zambonelli, P. & Davoli, R. Muscle transcriptome analysis identifies genes involved in ciliogenesis and the molecular cascade associated with intramuscular fat content in Large White heavy pigs. PLoS One 15, e0233372 (2020).

15. Wheway, G. et al. An siRNA-based functional genomics screen for the identification of regulators of ciliogenesis and ciliopathy genes. Nat. Cell Biol. 17, 1074–1087 (2015).

16. Klena, N. T., Gibbs, B. C. & Lo, C. W. Cilia and Ciliopathies in Congenital Heart Disease. Cold Spring Harbor Perspectives in Biology vol. 9 a028266 (2017).

17. Li, Y. et al. Global genetic analysis in mice unveils central role for cilia in congenital heart disease. Nature 521, 520–524 (2015).

18. Chevalier, B. et al. miR-34/449 control apical actin network formation during multiciliogenesis through small GTPase pathways. Nat. Commun. 6, 8386 (2015).

19. Shida, T., Cueva, J. G., Xu, Z., Goodman, M. B. & Nachury, M. V. The major alpha-tubulin K40 acetyltransferase alphaTAT1 promotes rapid ciliogenesis and efficient mechanosensation. Proc. Natl. Acad. Sci. U. S. A. 107, 21517–21522 (2010).

20. Balestra, F. R., Strnad, P., Flückiger, I. & Gönczy, P. Discovering regulators of centriole biogenesis through siRNA-based functional genomics in human cells. Dev. Cell 25, 555–571 (2013).

21. He, X. et al. BAG6 is a novel microtubule-binding protein that regulates ciliogenesis by modulating the cell cycle and interacting with γ-tubulin. Exp. Cell Res. 387, 111776 (2020).

22. Rothé, B., Gagnieux, C., Leal-Esteban, L. C. & Constam, D. B. Role of the RNA-binding protein Bicaudal-C1 and interacting factors in cystic kidney diseases. Cellular Signalling vol. 68 109499 (2020).

23. Kwon, O. S. et al. Exon junction complex dependent mRNA localization is linked to centrosome organization during ciliogenesis. Nat. Commun. 12, 1351 (2021).

24. Jin, X. et al. L-type calcium channel modulates cystic kidney phenotype. Biochim. Biophys. Acta 1842, 1518–1526 (2014).

25. Rangel, L. et al. Caveolin-1α regulates primary cilium length by controlling RhoA GTPase activity. Sci. Rep. 9, 1116 (2019).

26. Fu, S. et al. Primary Cilia as a Biomarker in Mesenchymal Stem Cells Senescence: Influencing Osteoblastic Differentiation Potency Associated with Hedgehog Signaling Regulation. Stem Cells Int. 2021, 8850114 (2021).

27. Doornbos, C. & Roepman, R. Moonlighting of mitotic regulators in cilium disassembly. Cell. Mol. Life Sci. 78, 4955–4972 (2021).

28. Sieving, P. A. et al. Ciliary neurotrophic factor (CNTF) for human retinal degeneration: phase I trial of CNTF delivered by encapsulated cell intraocular implants. Proc. Natl. Acad. Sci. U. S. A. 103, 3896–3901 (2006).

29. Fischer, A. J. et al. Differential Gene Expression in Human Conducting Airway Surface Epithelia and Submucosal Glands. American Journal of Respiratory Cell and Molecular Biology vol. 40 189–199 (2009).

30. Failler, M. et al. Whole-genome screen identifies diverse pathways that negatively regulate ciliogenesis. Mol. Biol. Cell 32, 169–185 (2021).

31. Cao, J. et al. miR-129-3p controls cilia assembly by regulating CP110 and actin dynamics. Nat. Cell Biol. 14, 697–706 (2012).

32. Loktev, A. V. & Jackson, P. K. Neuropeptide Y family receptors traffic via the Bardet-Biedl syndrome pathway to signal in neuronal primary cilia. Cell Rep. 5, 1316–1329 (2013).

33. Marley, A., Choy, R. W.-Y. & von Zastrow, M. GPR88 reveals a discrete function of primary cilia as selective insulators of GPCR cross-talk. PLoS One 8, e70857 (2013).

34. Angrisani, A., Di Fiore, A., De Smaele, E. & Moretti, M. The emerging role of the KCTD proteins in cancer. Cell Commun. Signal. 19, 56 (2021).

35. Shamseldin, H. E. et al. The morbid genome of ciliopathies: an update. Genet. Med. 22, 1051–1060 (2020).

36. Das, A., Qian, J. & Tsang, W. Y. USP9X counteracts differential ubiquitination of NPHP5 by MARCH7 and BBS11 to regulate ciliogenesis. PLoS Genet. 13, e1006791 (2017).

37. McDonough, J. E. et al. Gene correlation network analysis to identify regulatory factors in idiopathic pulmonary fibrosis. Thorax 74, 132–140 (2019).

38. Bachmann-Gagescu, R. et al. The Ciliopathy Protein CC2D2A Associates with NINL and Functions in RAB8-MICAL3-Regulated Vesicle Trafficking. PLoS Genet. 11, e1005575 (2015).

39. Zhou, J., Yang, F., Leu, N. A. & Wang, P. J. MNS1 is essential for spermiogenesis and motile ciliary functions in mice. PLoS Genet. 8, e1002516 (2012).

40. Avolio, R. et al. Protein Syndesmos is a novel RNA-binding protein that regulates primary cilia formation. Nucleic Acids Res. 46, 12067–12086 (2018).

41. Miyamoto, T. et al. Insufficiency of ciliary cholesterol in hereditary Zellweger syndrome. EMBO J. 39, e103499 (2020).

42. Chen, Q. et al. Prdx1 promotes the loss of primary cilia in esophageal squamous cell carcinoma. BMC Cancer 20, 372 (2020).

43. Vogt, J. et al. Mutation analysis of CHRNA1, CHRNB1, CHRND, and RAPSN genes in multiple pterygium syndrome/fetal akinesia patients. Am. J. Hum. Genet. 82, 222–227 (2008).

44. Martinez-Sanz, J. et al. Binding of human centrin 2 to the centrosomal protein hSfi1. FEBS J. 273, 4504–4515 (2006).

45. Minin, G. D. et al. Tmed2regulates Smoothened trafficking and Hedgehog signalling. doi:10.1101/2020.04.20.049957.

46. Jobling, A. I. et al. The Role of the Microglial Cx3cr1 Pathway in the Postnatal Maturation of Retinal Photoreceptors. J. Neurosci. 38, 4708–4723 (2018).

47. Westlake, C. J. et al. Primary cilia membrane assembly is initiated by Rab11 and transport protein particle II (TRAPPII) complex-dependent trafficking of Rabin8 to the centrosome. Proc. Natl. Acad. Sci. U. S. A. 108, 2759–2764 (2011).

48. Karunakaran, K. B., Chaparala, S., Lo, C. W. & Ganapathiraju, M. K. Cilia interactome with predicted protein–protein interactions reveals connections to Alzheimer’s disease, aging and other neuropsychiatric processes. Scientific Reports vol. 10 (2020).

49. Kyun, M.-L., et al. Wnt3a Stimulation Promotes Primary Ciliogenesis through β-Catenin Phosphorylation-Induced Reorganization of Centriolar Satellites. Cell Rep. 30, 1447–1462.e5 (2020).

50. Thesis title “KRAB zinc finger protein-mediated control of transposon-embedded regulatory sequences from human germline to early embryos” (https://infoscience.epfl.ch/record/286266)

